# CBioProfiler: a web and standalone pipeline for cancer biomarker and subtype characterization

**DOI:** 10.1101/2022.01.17.22269448

**Authors:** Xiaoping Liu, Zisong Wang, Hongjie Shi, Sheng Li, Xinghuan Wang

## Abstract

Cancer is a leading cause of death worldwide, and the identification of biomarkers and subtypes that can predict the long-term survival of cancer patients is essential for their risk stratification, treatment, and prognosis. However, there are currently no standardized tools for exploring cancer biomarkers or subtypes. In this study, we introduce CBioProfiler, a web server and standalone application that includes two pipelines for analyzing cancer biomarkers and subtypes. The cancer biomarker pipeline consists of five modules for identifying and annotating cancer survival-related biomarkers using multiple machine learning survival algorithms. The subtype pipeline includes three modules for data preprocessing, subtype identification using multiple unsupervised machine learning methods, and subtype evaluation and validation. CBioProfiler also includes a novel R package, CuratedCancerPrognosisData, which has reviewed, curated, and integrated gene expression data and clinical data from 268 gene expression studies of 43 common blood and solid tumors, including data from 47,686 clinical samples. The web server is available at https://www.cbioprofiler.com/ and https://cbioprofiler.znhospital.cn/CBioProfiler/, and the standalone app and source code can be found at https://github.com/liuxiaoping2020/CBioProfiler.

## Introduction

In recent years, with the advancement of high-throughput sequencing technologies including DNA sequencing, RNA sequencing, microarray, single-cell sequencing, etc., and their wide application in medical research and clinical practice, a large number of gene expression profiling studies of cancer patients has been published[1, 2]. The gene expression data of these gene expression profiling studies and the clinical data of the corresponding patients are mostly stored in public databases such as GEO (Gene Expression Omnibus) (https://www.ncbi.nlm.nih.gov/geo/), TCGA (The Cancer Genome Atlas) (https://portal.gdc.cancer.gov/), ArrayExpress (https://www.ebi.ac.uk/arrayexpress/), TARGET (Therapeutically Applicable Research To Generate Effective Treatments) (https://ocg.cancer.gov/programs/target), ICGC (International Cancer Genome Consortium) (https://dcc.icgc.org/), CGGA (Chinese Glioma Genome Atlas) (http://www.cgga.org.cn/), etc., and some of them are uploaded as research supplements on the official website of the journal or related research institutions. However, given that the data storage, preprocessing, and operation interface of each database are not completely the same, and the clinical data and gene expression data of each study have great differences in data collection, preprocessing, format, and documentation, people who want to make full and effective use of these high-throughput data to serve their research and guide clinical practice are facing great obstacles.

With the aging of the population and changes in people’s lifestyles, malignant tumors have become one of the main killers threatening human health and life[3, 4]. Therefore, the development and validation of novel tumor markers and subtypes that can be used for tumor diagnosis, risk stratification, and prognosis are of great significance for the early detection and personalized treatment of tumors. With the advancement of artificial intelligence, more and more machine learning strategies have been applied to the screening and validation of biomarkers and subtypes for cancer patients[5–7]. However, due to the lack of a unified, standardized and rigorous model and variable selection process, the reliability of relevant biomarkers and subtypes is questionable.

Thus, in the present study, we developed and introduced CBioProfiler (**C**ancer **Bio**marker and subtype **Profiler**), a web server and standalone application that reviewed, curated and integrated the gene expression data and corresponding clinical data of 47,686 samples from 268 gene expression studies of 43 common blood and solid tumors, for screening, validation, and annotation of cancer biomarkers and subtypes from molecular level to clinical settings (Figure 1 and https://github.com/liuxiaoping2020/CBioProfiler_tutorial/blob/main/CBioProfiler_tutorial.pdf).

**Figure 1.**
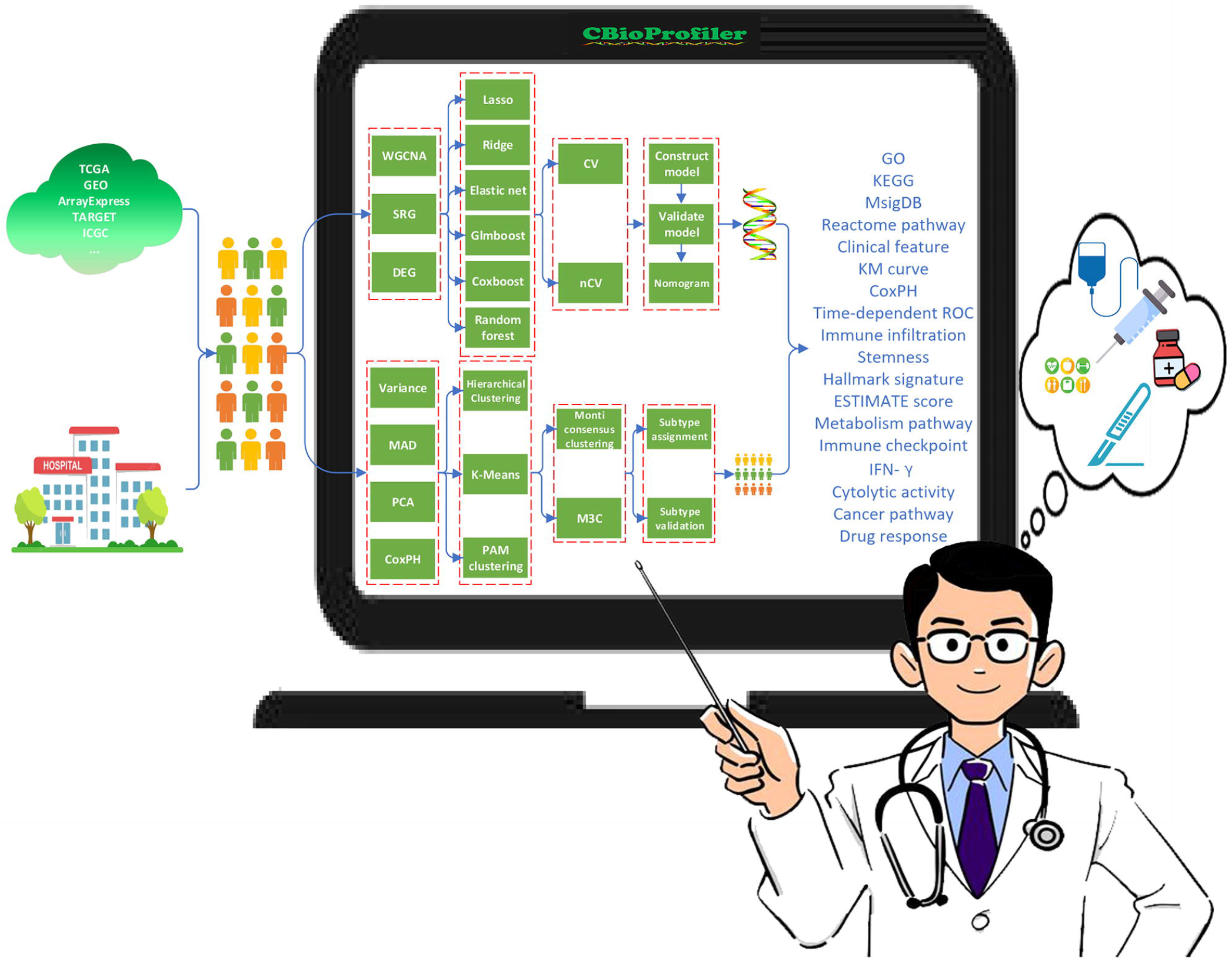
Overview of CBioProfiler. CBioProfiler was developed to facilitate researchers and clinicians to screen, characterize, annotate and translate cancer biomarkers and subtypes from molecular level to clinical settings more comfortably with graphical user interfaces (GUI), which will help implement targeted clinical diagnosis and treatment measures for different patients to achieve precision medicine. The whole pipeline of CBioProfiler includes two main pipelines: cancer biomarker pipeline and cancer subtype pipeline. The cancer biomarker pipeline includes 5 modules: (1) dimensionality reduction using three methods of weighted gene co-expression network analysis (WGCNA), univariate Cox proportional hazards regression model (CoxPH), differentially expressed gene (DEG) analysis, (2) benchmark experiment with 6 machine learning learners (Lasso, Ridge, Elastic net, Glmboost, Coxboost, Randomforest) using cross validation (CV) and nested cross validation (nCV) based on R package mlr, (3) prediction model construction using Cox proportional hazards regression model and nomogram, (4) clinical annotation using a variety of clinical approaches (correlation with clinical features, Kaplan-Meier curve, CoxPH model, time-dependent ROC, most correlated genes, correlation with specific gene, gene expression in different groups, correlation with immune infiltration, correlation with stemness score, correlation with ESTIMATE score, correlation with immune checkpoint, correlation with IFN-gamma score, correlation with cytolytic activity, correlation with cancer pathway, correlation with metabolism pathway, correlation with hallmark signature, correlation with drug response, and (5) biological annotation using over-representation analysis (ORA) and gene set enrichment analysis (GSEA). The subtype pipeline includes 3 modules: (1) data preprocessing (feature selection based on variance, median absolute deviation (MAD), CoxPH model, and principal component analysis (PCA), (2) subtype identification (integration of multiple unsupervised machine learning methods (K-means clustering (K-means), hierarchical clustering, partitioning around medoids (PAM) clustering, etc.) using two popular consensus clustering methods (ConsensusClusterPlus and M3C), (3) subtype evaluation and validation. **Abbreviations**: TCGA, The Cancer Genome Atlas; GEO, gene expression omnibus; TARGET, Therapeutically Applicable Research To Generate Effective Treatments; GEP, expression profile; ICGC, International Cancer Genome Consortium; WGCNA, weighted gene co-expression network analysis; SRG, survival-related gene; DEG, differentially expressed gene; CV, cross validation; nCV, nested cross validation;

## Results

### Curated public gene expression studies

We have reviewed, curated, normalized and integrated the gene expression data and corresponding clinical data of 43 common blood and solid tumors from GEO, TCGA, ICGC, TARGET, ArrayExpress and other public databases. These public data from 47,686 clinical samples of 268 gene expression studies (Figure 2, Supplementary table 1 and https://liuxiaoping2020.github.io/CBioProfilerDatasource/) was integrated to build an R package “CuratedCancerPrognosisData”, the associated source code was deposited at https://zenodo.org/record/5728447#.Ya9vhsj1dk4.

**Figure 2.**
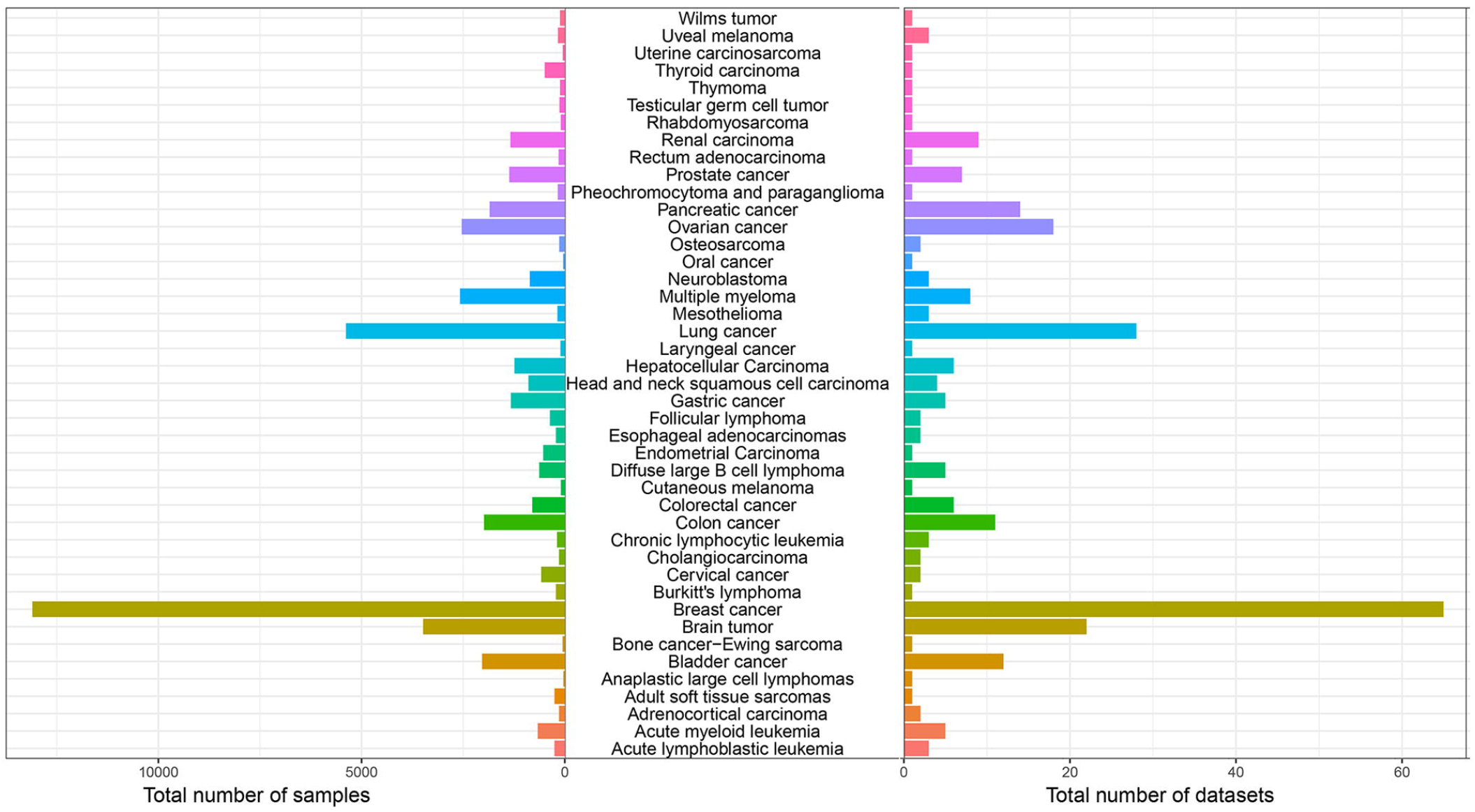
Total number of samples and datasets of included in CuratedCancerPrognosisData.

### Compared with other similar online tools or standalone apps

As shown in table 1, compared with other similar tools, CBioProfiler is significantly better than them in terms of data, features, enrichment analysis and software: (1) CBioProfiler includes the largest disease types and samples, and provides a personalized data submission interface for researchers to analyze their own data. (2) CBioProfiler covers the most functional modules. (3) CBioProfiler provides an online version and a standalone local app version.

### Case study: dimensionality reduction

CBioProfiler enables 3 methods (weighted gene co-expression analysis (WGCNA), survival related gene (SRG), differentially expressed gene (DEG)) to conduct dimensionality reduction. WGCNA[8] is a biometric method that can cluster genes with similar expression patterns or functions into the same module, while unassigned genes are categorized into grey modules. Bladder cancer represents one of the most common types of malignant cancers of human genitourinary system[9]. Kim WJ et al. published a far-reaching bladder cancer gene expression study GSE13507, which evaluated the predictive effect of bladder cancer prognosis-related genes for patients[10]. In this example, we used the WGCNA to perform dimensionality reduction on the GSE13507, and then screened genes that were closely related to the patient’s overall survival (OS) for subsequent studies. After Euclidean distance-based clustering, 3 samples were detected as outliers, and the remaining 162 samples were used to construct co-expression network (Supplementary figure 1). Then, according to the soft-thresholding power 8 (Figure 3A), we constructed a co-expression network, which clustered all genes into 10 modules (Figure 3B). Next, we analyzed the relationship between the gene modules and the clinical features of bladder cancer patients. As a result, ‘green’ module was most positively relevant to the OS, while “blue” module was most negatively correlated with the OS of bladder cancer patients (Figure 3C-D). Finally, we could output the genes of any module or non-gray module for subsequent research.

**Figure 3.**
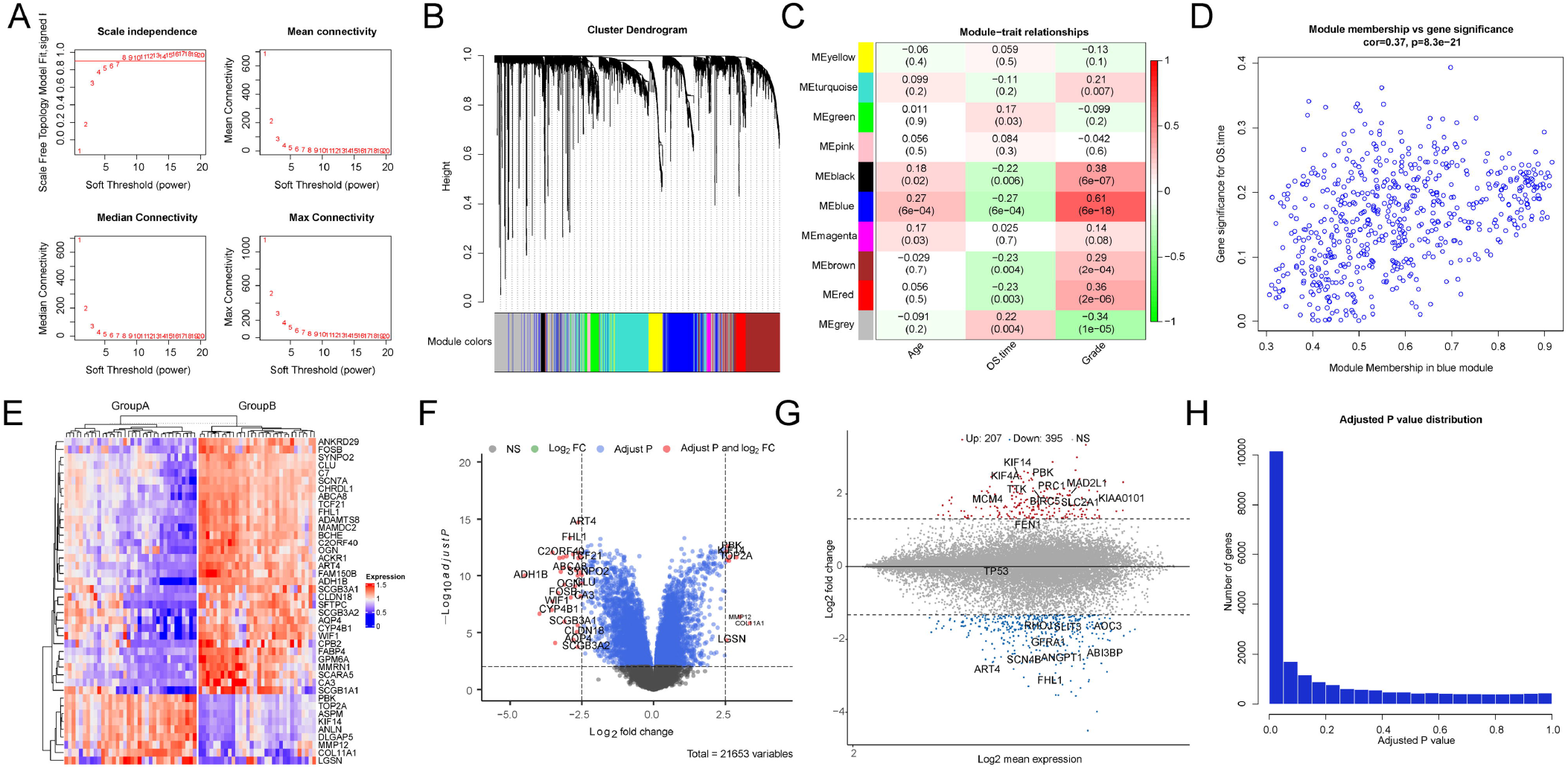
Main result output of WGCNA and DEG analysis. (A) Selection of soft threshold power; (B). Modules detected by network analysis. (C) Module-trait relationships. (D) Gene Significance (GS) for grade vs. Module Membership (MM) in the turquoise module. (E) Heatmap showing differentially expressed genes in group A and group B. (F) Volcano plot showing differentially expressed genes in group A and group B. (G) MA plot showing differentially expressed genes in group A and group B. (H) P adjusting plot of differentially expressed gene analysis.

Cox proportional hazards regression model (CoxPH) is often used clinically to evaluate the impact of clinical phenotypes on patient survival. Thus, we use the breast cancer gene expression study METABRIC[11] to illustrate the use of univariate CoxPH for dimensionality reduction analysis. The METABRIC project introduced a novel risk stratification system for patients with breast cancer based on multi-omics high-throughput data. We performed univariate CoxPH to analyze the impact of a single gene on the OS of breast cancer patients. We included the top 60 genes that are closely related to the OS of patients (Supplementary table 2).

Exploring differentially expressed genes (DEGs) between biological groups is of great significance to clarify the biological significance of the groups. Therefore, it is also very recommended to screen DEGs for dimensionality reduction analysis. Okayama et al. conducted transcription profiling of 226 stage I–II lung adenocarcinomas, which clustered these lung adenocarcinomas into two groups (group A: patients that are mainly males, ever-smokers, and advanced stages; group B: patients that are mainly never-smokers and early stages)[12]. As shown in figure 3E-H, there were 42 significant DEGs detected at adjusted P value < 0.01 and absolute log fold change > 2.5 between group A lung adenocarcinomas and group B lung adenocarcinomas. The DEGs were visualized using heatmap, volcano plot, MA plot, and adjusted P plot.

### Case study: building and validating prediction model and constructing associated nomogram

To build a prediction model, METABRIC was treated as the discovery cohort, which was then randomly stratified into training set and test set according to a ratio of 0.85. The 60 genes selected by the CoxPH were applied to train 3 machine learning models (lasso, elastic net, and glmboost) to create a prediction model based on 10-fold cross validation. The performances of these models were then evaluated and compared using 100 bootstraps. As shown in supplementary figure 2, the performance of elastic net outperformed both lasso and glmboost. Thus, elastic net was selected to build the prediction model. Results of time-dependent receiver operating characteristic curve (ROC) suggested that the areas under the curves (AUCs) of the prediction model in the training set at 1-, 3-, 5-, and 7-years were 0.597, 0.713, 0.700, and 0.693, respectively (Figure 4A), while the corresponding AUCs in the test set were 0.708, 0.773, 0.775 and 0.703, respectively (Figure 4B). KM plot suggested that patients in the low-risk group had significant better OS compared to those in the high-risk group in training set and test set (Figure 4D-E). Moreover, the risk score remained an independent prognostic factor after adjusting for other clinical features of breast cancer patients (Supplementary figure 3).

**Figure 4.**
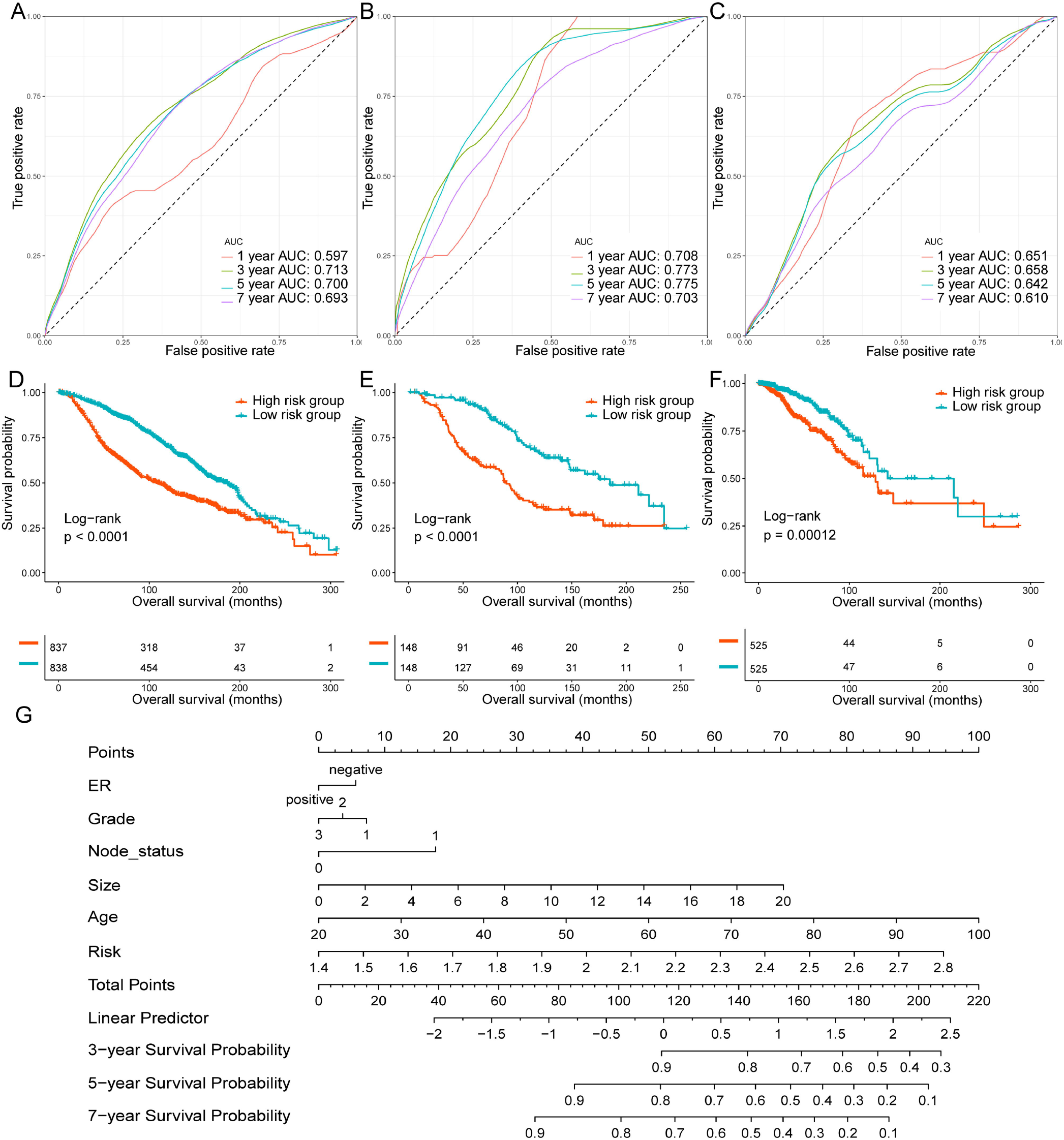
Construction and evaluation and translation of the prediction model. (A) Areas under the curve (AUCs) of time dependent ROC analysis at 3-, 5-. 10-, 15-year in the training set. (B) AUCs of time dependent ROC analysis at 3-, 5-. 10-, 15-year in the test set. (C) AUCs of time dependent ROC analysis at 3-, 5-. 10-, 15-year in the validation set. (D) Survival differences between the low-risk group and high-risk group in the training set. (E) Survival differences between the low-risk group and high-risk group in the test set. (F) Survival differences between the low-risk group and high-risk group in the validation set. (G) Nomogram prediction the 3-, 5-, 7-year survival probabilities based on CoxPH model that integrates the risk, ER status, tumor size, node status, age, and grade of breast cancer patients.

TCGA-BRCA[13] is an independent multi-omics study of breast cancer, thus, we use it as an external validation cohort. Time-dependent ROC analysis suggested that the AUCs of the prediction model in the validation cohort were 0.651, 0.658, 0.642, and 0.610 at 1-, 3-, 5-, and 7-years respectively (Figure 4C) and the risk score could also stratify patients into different risk groups (Figure 4F, Supplementary figure 4).

In order to facilitate the prediction of a patient’s long-term survival rate by physicians and researchers, CBioProfiler can generate a nomogram to predict the patient’s long-term survival rate. In this example, we included the patient’s risk score, ER status, tumor size, lymph node metastasis status, age and grade to draw a nomogram predicting the OS probabilities of patients at 3-, 5-, and 10-years (Figure 4G). The users can estimate the survival probability of each patient based on the ‘Total point’ which is the sum of the ‘Points’ corresponding to each clinical feature. Then, we internally and externally validated and calibrated the performance of the nomogram (Supplementary figure 5, 6).

### Case study: clinical annotation

In order to further clarify the clinical significance of the molecular markers screened by CBioProfiler, we chose ABAT (4-aminobutyrate aminotransferase) for clinical annotation analysis. As shown in supplementary table 3, the expression level of ABAT was closely related to the patient’s ER, tumor size, and grade. Survival analysis showed that the OS of breast cancer patients in the ABAT high expression group was significantly better than that in the ABAT low expression group (Figure 5A). After adjusting for other clinical factors, the expression level of ABAT still has independent prognostic significance for breast cancer patients (Figure 5B). Correlation analysis between the expression of ABAT and the enrichment score of ssGSEA analysis of the gene sets provided by Bindea et al.[14] suggested that the expression of ABAT was significantly correlated with enrichment score of B cells, CD8 T cells, cytotoxic cells, dendritic cells, eosinophils, immature dendritic cells, macrophages, neutrophils, NK CD56bright cells, NK CD56dim cells, plasmacytoid dendritic cells, T cells, T helper cells, T central memory cells, T effector memory cells, T follicular helper cells, T gamma delta cells, Th1 cells, Th17 cells, Treg cells, angiogenesis, and antigen presentation machinery (Figure 5C). Meanwhile, the expression of ABAT was negatively correlated several well-known immune checkpoint molecules (PDCD1, CD274, PDCD1LG2, CTLA4, PVR, LAG3, TIGIT, HAVCR2, VTCN1, CD86, CD28, CD80, CD27, CD40, IL2RB, TNFRSF9, TNFRSF4, ICOS, CD276, BTLA, KIR3DL1, CYBB, and SIGLEC7, Supplementary figure 7) and stromal score, immune score and estimate score (Supplementary figure 8) calculated using R package ESTIMATE. The expression of ABAT in the ER-positive group was significantly lower than that in the ER-negative group (Figure 5D). ABAT expression was also correlated with interferon gamma score (Figure 5E), stemness score (Figure 5F) and cytotoxic activity (Supplementary figure 9) of breast cancer. Finally, the expression of ABAT were significant correlated with many well-known cancer related signaling pathways (Supplementary figure 10), hallmarks signatures (Supplementary figure 11), and metabolism pathways (Supplementary figure 12).

**Figure 5.**
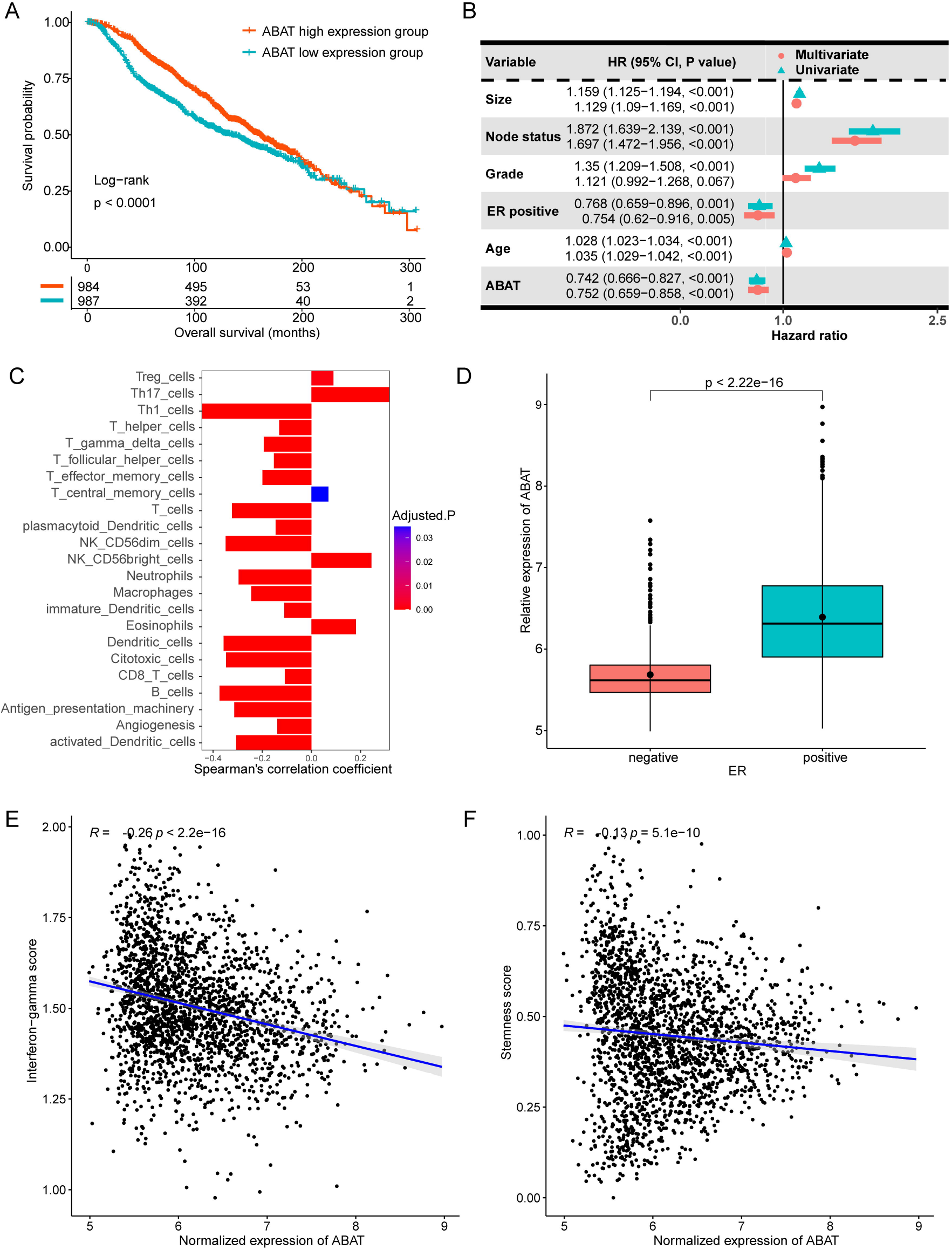
Clinical annotation ABAT in the METABRIC cohort. (A) Survival differences between ABAT low and high expression groups. (B) CoxPH model identifying the prediction ability of ABAT. (C) Most correlated genes of ABAT based on Spearman’s correlation analysis. (D) Relative expression of ABAT in the ER-negative and ER-positive groups. (E) Correlation between the relative expression ABAT and the enrichment score of stemness. (F) Correlation between the relative expression ABAT and the enrichment scores of the immune cells infiltrated in the microenvironment of breast cancer cells.

### Case study: biological annotation

CBioProfiler provides a variety of enrichment analysis and corresponding visualization methods so that researchers can clarify the biological function and significance of the biomarkers they screened. In this example, we performed gene ontology (GO) enrichment analysis on 136 DEGs between group A and group B at |logFC| > 2 in GSE31210[12] by CBioProfiler. Over representation analysis (ORA) and gene set enrichment analysis (GSEA) were implemented, respectively. Figure 6A-C showed the top 10 GO terms (sister chromatid segregation, mitotic sister chromatid segregation, mitotic nuclear division, nuclear division, organelle fission, nuclear chromosome segregation, mitotic cell cycle phase transition, chromosome segregation, spindle organization, and mitotic spindle organization) that the 136 genes were enriched in. While results of GSEA suggested that these genes were mainly enriched in cell cycle, mitotic cell cycle, cell cycle process, and nuclear division (Figure 6D-H).

**Figure 6.**
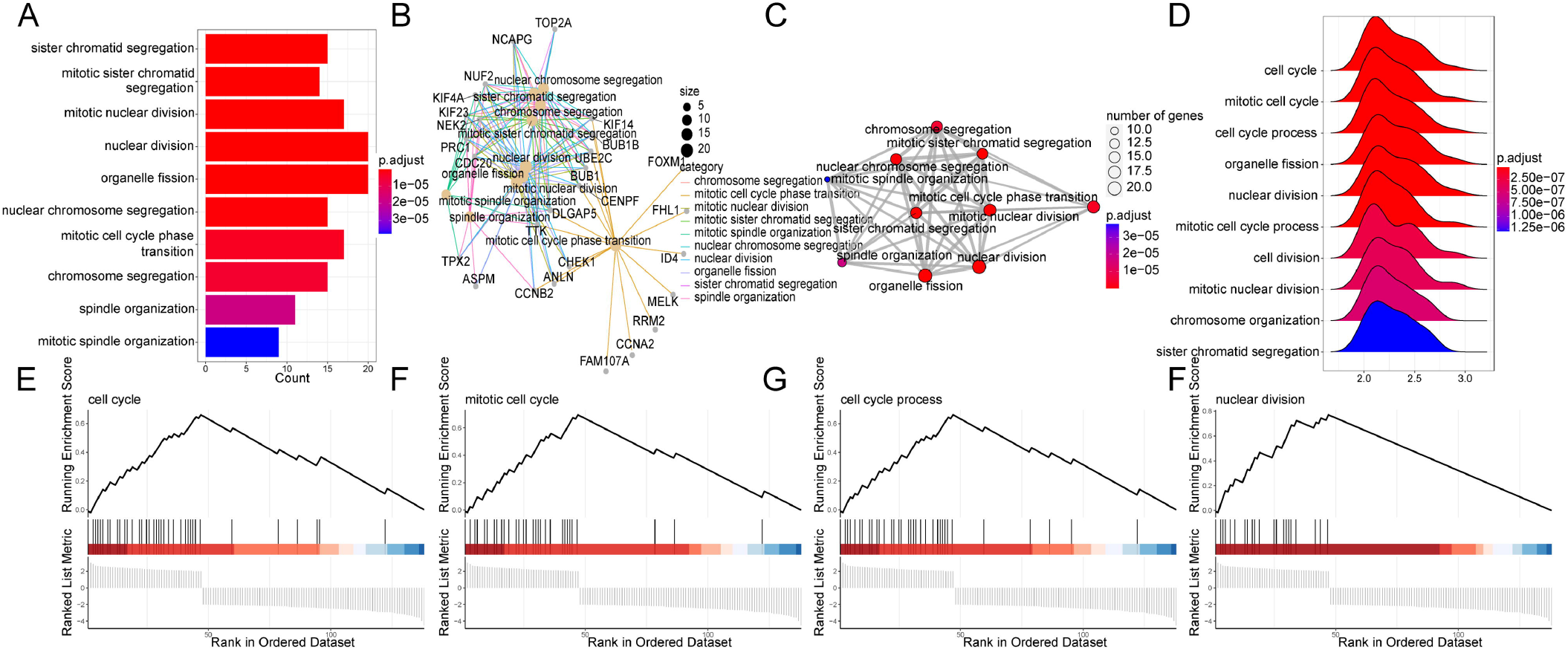
Functional enrichment analysis of the biomarkers identified by CBioProfiler. (A-D) Top 10 gene ontology biological process terms that enriched in the biomarkers. (E-H) Top gene ontology biological process terms identified by gene set enrichment analysis.

### Case study: cancer subtype identification, validation, and annotation

As mentioned above, GSE31210, published by Okayama et al., is a transcription profiling of 226 stage I–II lung adenocarcinomas[12]. Herein, we performed a Monte Carlo simulation-based consensus clustering analysis [15] of lung adenocarcinoma gene expression profiles that had been variable screened using the CoxPH to identify potential subtypes of lung adenocarcinomas in GSE31210 cohort (training set) (Supplement 1, Figure 7A, supplementary figure 13) and validated the subtypes on the TCGA-LUAD cohort (validation set)[16] (Supplement 2).

**Figure 7.**
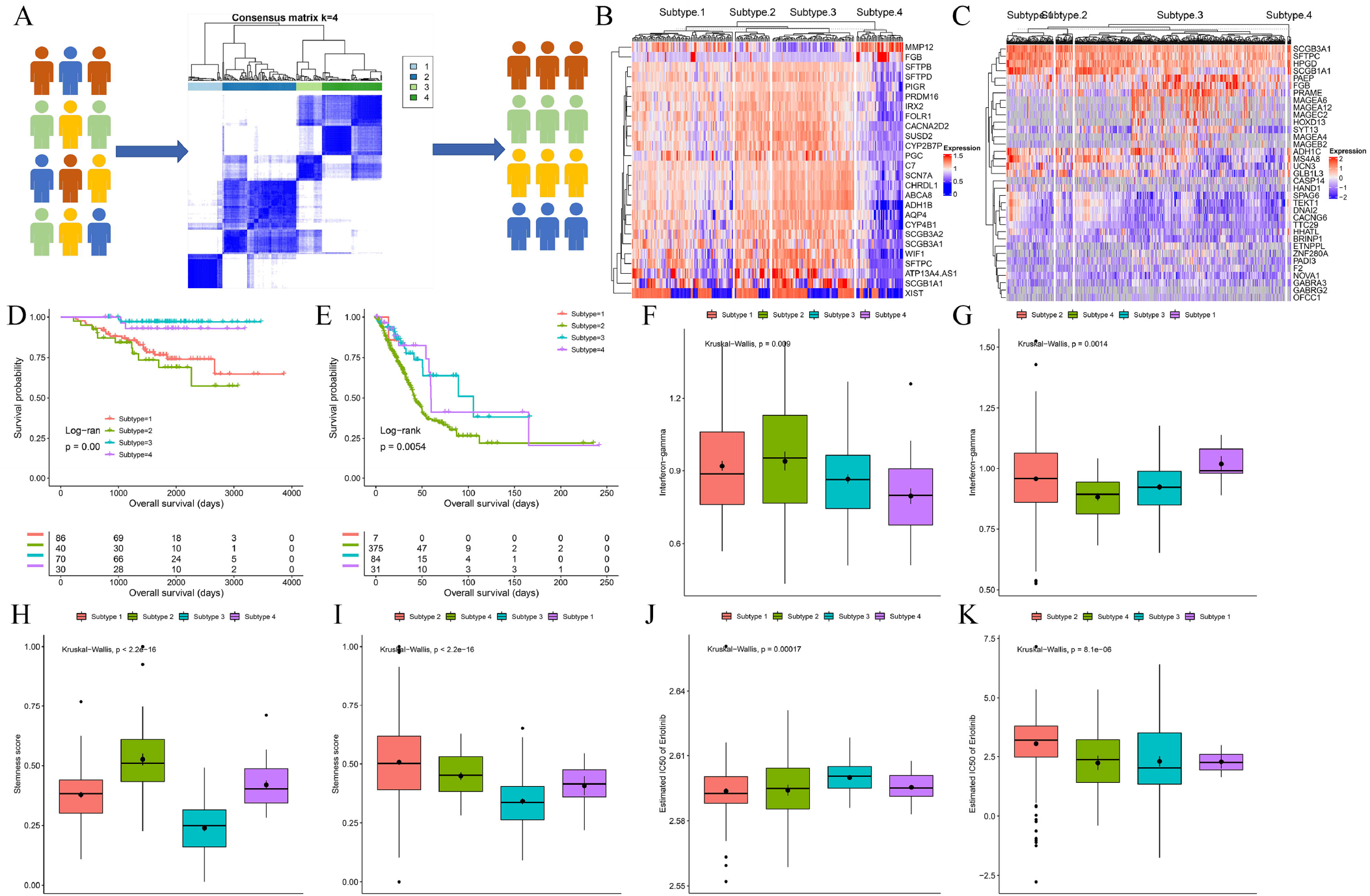
Identification, validation and characterization of cancer biomarkers. (A) Preprocessed gene expression profile of GSE31210 (training set) was subjected to Monte Carlo simulation-based consensus clustering to identify cancer subtypes and validated on TCGA-LUAD cohort (validation set). (B) Differentially expressed genes among the different subtypes in the training set. (C) Differentially expressed genes among the different subtypes in the validation set. (C) Survival differences of patients among the different subtypes in the training set. (E) Survival differences of patients among the different subtypes in the validation set. (F) Comparison of interferon-gamma score among the different subtypes in the training set. (G) Comparison of interferon-gamma score among the different subtypes in the validation set. (H) Comparison of stemness score among the different subtypes in the training set. (I) Comparison of stemness score among the different subtypes in the validation set. (J) Comparison of response of erlotinib among the different subtypes in the training set. (K) Comparison of response of erlotinib among the different subtypes in the validation set.

In order to further clarify the potential clinical and biological significance of different subtypes, we compared the differences between different subtypes in the two cohorts from multiple aspects. Figures 7B and 7C showed the DEGs among different subtypes in the two cohorts, respectively. Survival analysis showed that there were significant differences among different subtypes (Figure 7D-E), and the corresponding time-dependent ROC analysis confirmed that the subtype had good predictive performance (Supplementary figure 14).

In addition, patients with different subtypes of lung adenocarcinoma had significant differences in clinical features (Supplementary table 4-5), interferon gamma score (Figure 7F-G), stemness score (Figure 7H-I), erlotinib response (Figure 7J-K), cancer-related signaling pathways (Supplementary figure 15-16), ESTIMATE score (Supplementary figure 17-18), Hallmark signature score (Supplementary figure 19-20), immune microenvironment (Supplementary figure 21-22), immune checkpoints (Supplementary figure 23-24), and metabolic related signaling pathways (Supplementary figure 25-26).

### Case study: meta-analysis

To assist users in synthesizing and drawing conclusions from a larger body of evidence, CBioProfiler enables the performance of meta-analysis on the predictive ability of a biomarker using multiple gene expression and prognosis studies. This facilitates the generation of more robust and reliable results compared to individual studies alone. As mentioned above, the prognostication ability of ABAT in breast cancer has been analyzed in the breast cancer gene expression study METABRIC. To draw a more robust and generalized conclusion, we included a total of 31 breast cancer gene expression studies (GSE3143, GSE10886_GPL1390, GSE10886_GPL887, GSE18229_GPL1390, GSE18229_GPL887, GSE22226_GPL1708, GSE22226_GPL4133, GSE2607_GPL1390, GSE2607_GPL887, GSE6130_GPL1390, GSE6130_GPL887, Caldas_2007, GSE12071, GSE10510, GSE159956, GSE22133_GPL5345, E_TABM_158, GSE1456_GPL96, GSE16446, GSE20711, GSE37751, GSE42568, GSE48390, GSE58812, GSE7390, ICGC_BRCA_FR, ICGC_BRCA_KR, TCGA_BRCA, METABRIC, NKI, and UCSF) containing 6992 patients with breast cancer. As shown in supplementary figure 27, the result of meta-analysis confirmed that ABAT was significantly associated with the overall survival of patients with breast cancer (HR=0.83, 95% CI: 0.75-0.9, P < 0.01).

## Discussions

CBioProfiler, to our knowledge, is the first web and standalone app that integrates multiple popular machine learning algorithms and cross-validation strategies for tumor biomarker and subtype identification, validation, and clinical and biological annotation. Moreover, we also developed CuratedCancerPrognosisData, an R package that reviewed, integrated 47,686 clinical samples from 268 gene expression studies of 43 common blood and solid tumors. Compared with other similar online tools or standalone apps[17–28] (table 1) based on public data from TCGA, GEO, etc., CBioProfiler has the following advantages: (1) CBioProfiler includes the largest disease types and samples, completely open to download the curated data for academic use, and provides a personalized data submission interface for researchers to analyze their own data. (2) CBioProfiler offers the most comprehensive set of analysis modules. These modules can be used either as a complete pipeline for screening, evaluating, validating, and annotating tumor molecular markers or individually to achieve specific research goals. For example, users can perform DEG, WGCNA, and SRG analyses separately, or they can perform clinical annotations, such as survival analysis and immune infiltration analysis, for some molecular markers they are interested in. (3) CBioProfiler is available in both an online version and a standalone local app version. The online version allows users to quickly conduct research on related molecular markers, while the standalone app version can be downloaded for more computationally intensive analysis.

We developed and introduced CBioProfiler, a web server and standalone application that reviewed, curated, and integrated the gene expression data and corresponding clinical information of 47,686 clinical samples from 268 gene expression studies of 43 common blood and solid tumors. This was done in order to identify, validate, and annotate cancer biomarkers and subtypes from the molecular level to clinical settings.

## Methods

### Data collection and curation

We searched and downloaded gene expression profile studies of tumor patients from GEO, TCGA, ICGC, ArrayExpress, TARGET, CGGA, and other public databases or websites.

The following criteria were used to include datasets in our research: (1) The research subjects were cancer patients; (2) The dataset contained gene expression profiling data; (3) The dataset reported at least one type of follow-up information, such as overall survival (OS), progression-free survival (PFS), relapse-free survival (RFS), disease-free survival (DFS), or distant metastasis-free survival (DMFS); (4) The sample size of the dataset was greater than 20. For data from GEO and ArrayExpress, R package GEOquery (version: 2.56.0)[29] and ArrayExpress(version: 1.48.0)[30] were used to download it, respectively. If the raw CEL data was available, robust multichip average (RMA) method[31] was used to normalize the raw data using R package affy (version: 1.66.0)[32] or oligo (version: 1.52.1)[33], otherwise, the normalized data was used. For data from TCGA and MMRF-CoMMpass[34, 35], we downloaded the RNA-seq count data and transformed it to transcripts per kilobase million (TPM) value from GDC using R package TCGAbiolinks (version: 1.14.0)[36]. For data from TARGET, ICGC, and CGGA, normalized data was downloaded and used indirectly. Annotation files provided by the submitters were used to annotate the gene expression profiling data. When multiple probes match to the same gene, we choose the most variant probe, and when multiple genes correspond to the same probe, we dropped them since unspecific annotation. Clinical data was uniformly reformatted and curated using in-house R scripts for each dataset, and independent double-checking was conducted by investigators to ensure the accuracy of the curation. The workflow of the curation is summarized in supplementary figure 28.

### Cancer biomarker pipeline

#### Dimensionality reduction

CBioProfiler uses three of the most commonly used bioinformatics methods to reduce the dimensionality of data: (1) WGCNA[36]; (2)SRGs[37]; (3) DEGs[38]. WGCNA includes 3 steps: firstly, Euclidean distance-based sample network is used to filter outlying samples, and then a weighted gene co-expression network is constructed to identify gene modules whose expression profiles are similar based on adjacency matrix and appropriate soft threshold. Finally, correlations between gene modules and clinical features are calculated. Empirical Bayesian methods is used to perform DEG analysis using R package limma (version: 3.44.3)[38]. SRG is implemented based on univariate CoxPH model using R package survival (version: 3.2-3).

### Survival learners

For the benchmark experiment, CBioProfiler includes 5 embedded machine learning algorithms, including penalized Cox regression (PCR) with LASSO[39, 40] and Ridge[41, 42], Elastic net[43], Glmboost[44], Coxboost[45], and RandomForestSRC[46] for survival analysis. LASSO (least absolute shrinkage and selection operator), proposed by Robert Tibshirani in 1996, obtains a more refined model by constructing a L1 norm penalty function, which forces the sum of absolute values of coefficients to be less than a certain fixed value and set some regression coefficients to zero. Therefore, it is a regression analysis method that performs feature selection and regularization at the same time, and aims to enhance the prediction accuracy and interpretability of statistical models[39]. Ridge regression is similar to linear regression, both of which are to solve the over-fitting problem of standard linear regression. The difference is that ridge regression adds the L2 norm penalty[42]. Elastic network integrates the L1 norm and the L2 norm, which makes it have both the variable selection and regularization advantages of lasso and ridge regression[43]. Glmboost fits generalized linear model and conduct variable selection based on component-wise boosting[44]. Unlike glmboost, CoxBoost fits a CoxPH by component-wise likelihood based boosting[45]. For feature selection, the above five survival learners reserve features with non-zero coefficients. Random forest is an ensemble model, and the original random forest (RF) is only available for regression and classification. randomforestSRC extends RF to survival analysis and conducts variable selection based on maximal subtree information[46]. Parameter sets for the survival learners are summarized in supplementary table 6.

### Benchmark experiment

Benchmark experiment is supported by the R package mlr (version: 2.18.0)[47]. To train and validate the survival learners, we apply cross validation (CV) and nested cross validation (nCV) to perform a benchmark experiment. During CV, the whole data set is randomly split training set and test set, then k-fold CV is applied to the training set: (1) divide the training set into equal K folds; (2) use the first fold as inner test set, and the rest as inner training set. (3) train the model and calculate the C-index of the model on the inner test set; (4) use a different fold as inner test set each time, and repeat steps (2) and (3) K times. (5) apply the best model to test set and external independent validation cohort. CV is designed for model selection, when the best model is selected, bootstrap resampling can be used to evaluate and compare the performances of different survival learners. The workflow of CV is summarized in supplementary figure 29.

With respect to nCV, the whole data set is divided into N outer folds, and then each outer fold is divided into training set and test set. Then, the main steps of nCV can be summarized as: (1) divide the training set into equal K folds; (2) use the first fold as inner test set, and the rest as inner training set. (3) train the model and calculate the C-index of the model on the inner test set; (4) use a different fold as inner test set each time, and repeat steps (2) and (3) K times. (5) apply the best model to outer fold test set. (6) select the best outer model features and parameters and train on the whole data set to get final model. (7) if the users have divided the whole data set into two parts, one is for training nCV, the other is for validation, then they can validate the final model on the validation part and external validation cohort, otherwise, they can validate the final model on external cohort. nCV utilizes multi-layer CV to implement model selection. The workflow is summarized in supplementary figure 30.

### Prediction model

After the benchmark experiments are finished, CBioProfiler could calculate the fitted relative risk of patients in the training set, test set and external validation set. LASSO, Ridge, Elastic net, Glmboost, and Coxboost calculate relative risk using the predict function, while randomforestSRC uses the sum of cumulative hazard function (CHF). Thus, based on the relative risk score, we build and validate prediction model for certain survival endpoints using time-dependent receiver operating characteristic curve (ROC) (implemented with R package survivalROC (version: 1.0.3)[48]), Kaplan-Meier curve (implemented and visualized using R packages survival (version: 3.2-3) and survminer (version: 0.4.8) and CoxPH. Moreover, CBioProfiler allows to construct nomogram that included the relative risk score and other clinical features, which helps researchers and physicians predict the survival probability of cancer patients. The nomogram can be internally and externally validated and calibrated based on bootstrap resampling and calibration analysis.

### Clinical annotation

To help users investigate the clinical relevance of the biomarkers they identified, CBioProfiler allows users to analyze the correlation between a given biomarker and clinical features (“Correlation with clinical features” module), characterize the prognostication significance of given biomarkers (“Kaplan-Meier curve” module, “CoxPH model” module, and “Time-dependent ROC” module), identify genes correlated with given biomarkers (“Most correlated genes” module, “Correlation with specific gene” module), compare the expression levels of given biomarkers among different groups (“Gene expression in different groups” module), and investigate the relationship between given biomarkers and immune cell infiltration [49], cancer stemness score[50], and ESTIMATE score[51], immune checkpoint[52, 53], IFN-gamma score[54], cytolytic activity[55], cancer related signaling pathway[56], metabolism pathway[57], hallmark signature[58], drug response[59, 60]. For Spearman’ correlation and Pearson’s correlation are used for correlation analysis.

### Biological annotation

Yu and colleagues developed clusterProfiler (version: 3.18.1)[61], a very outstanding R language package for gene functional annotation. CBioProfiler integrates some useful functions of clusterProfiler to annotate tumor markers, allowing users to perform functional annotation of their biomarkers regarding gene ontology (GO), Kyoto Encyclopedia of Genes and Genomes (KEGG), Molecular Signatures Database (MSigDB), and Reactome pathway based on over representation analysis (ORA)[62, 63] and gene set enrichment analysis (GSEA)[64].

### Cancer subtype pipeline

The subtype pipeline includes 3 modules: (1) data preprocessing (feature selection based on variance, median absolute deviation (MAD), CoxPH model[65], and principal component analysis (PCA)[66], (2) subtype identification (integration of multiple unsupervised machine learning methods (K-means clustering[67], hierarchical clustering[68], partitioning around medoids (PAM) clustering[69], etc.) using two popular consensus clustering methods (ConsensusClusterPlus (version: 1.52.0)[70] and M3C (version: 1.10.0)[15]), (3) subtype evaluation and validation. In order to further clarify the biological and clinical significance of different subtypes, CBioProfiler also provides a variety of biological annotation modules (similar to biomarker modules). For group comparison, T, Anova, Kruskal-Wallis, and Wilcoxon test are available for use.

### Meta-analysis pipeline

CBioProfiler offers a Meta-analysis module that helps researchers evaluate the effect of biomarkers on disease prognosis. This module utilizes the methods of Schwarzer et al.[71], which involves calculating the correlation between a particular gene and the survival time of patients in a specific cohort using a univariate Cox proportional hazard model. The module then performs meta-analysis based on the hazard ratio and its 95% confidence interval of the patients.

## Declarations

### Ethics approval and consent to participate

All data utilized in this study has been previously published and is publicly available. The research performed in this study conformed to the principles of the Helsinki Declaration.

### Consent for publication

Not applicable

### Authors’ contributions

XL, XW, SL designed the study, XL developed and deployed the program, wrote and reviewed the manuscript, ZW participated in the development of the program, HS participated in deploying the program.

## Availability of data and materials

CBioProfiler is publicly available as a web server at https://www.cbioprofiler.com/ and https://cbioprofiler.znhospital.cn/CBioProfiler/, The source code for CuratedCancerPrognosisData is deposited at https://zenodo.org/record/5728447#.Ya9vhsj1dk4.

## Code availability

The associated source code of CBioProfiler can be downloaded from https://github.com/liuxiaoping2020/CBioProfiler or https://gitee.com/liuxiaoping2020/CBioProfiler.

## Data Availability

All data produced in the present work are contained in the manuscript

https://www.cbioprofiler.com/

https://cbioprofiler.znhospital.cn/CBioProfiler/

## Acknowledgement

We thank Dr. Shixiang Wang Sun Yat-sen University Cancer Center, for his important help in the deployment process of CBioProfiler.

