## Supplementary figures and tables for "CBioProfiler: a web and standalone pipeline for cancer biomarker and subtype characterization"

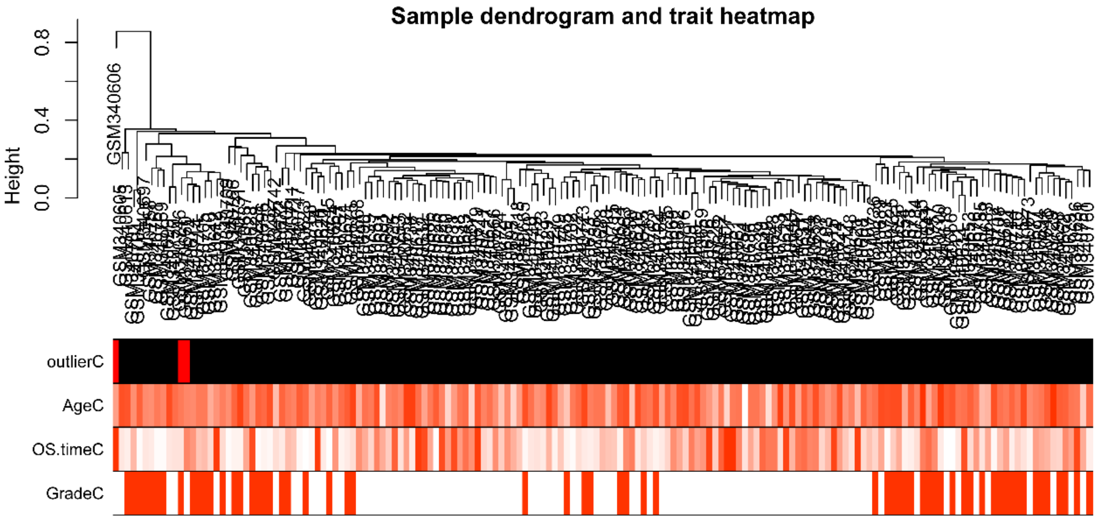


**Supplementary figure 1.** Detection of outliers based on sample dendrogram.

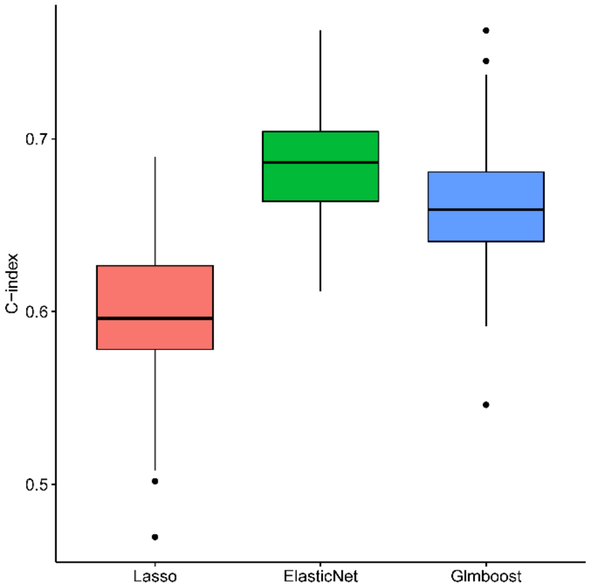


**Supplementary figure 2.** Comparison of C-indexes of three survival learners based on cross validation and bootstraps.

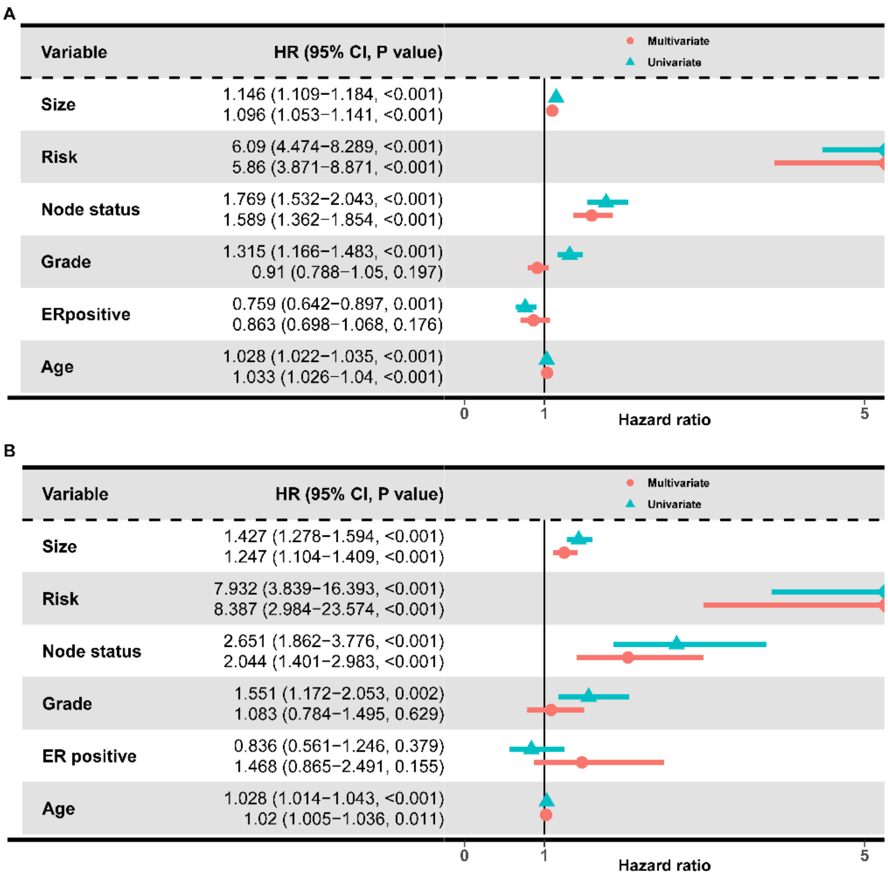


**Supplementary figure 3.** Cox proportion hazards regression model identifying independent prognostication role of the risk score in the training set (A) and test set (B).

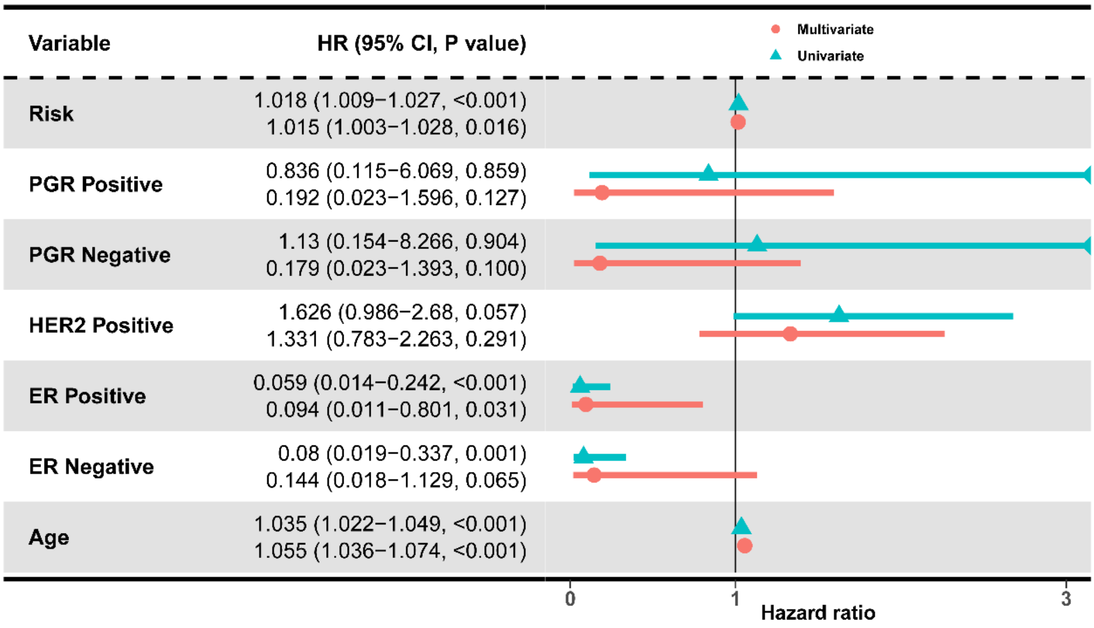


**Supplementary figure 4.** Cox proportion hazards regression model identifying independent prognostication role of the risk score in the validation set.

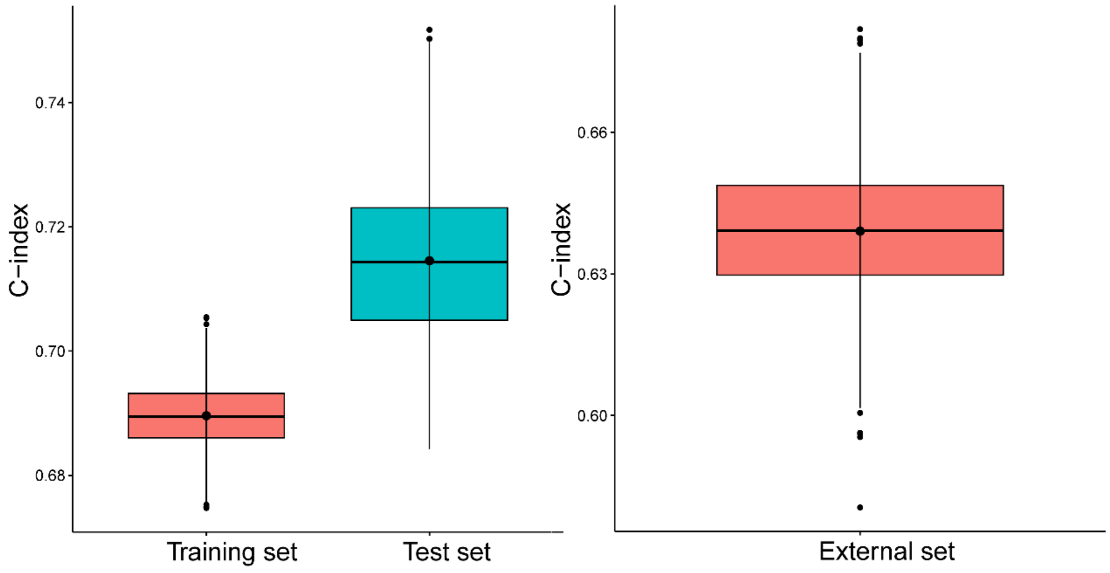


**Supplementary figure 5.** Internally (A) and externally (B) validation of the nomogram.

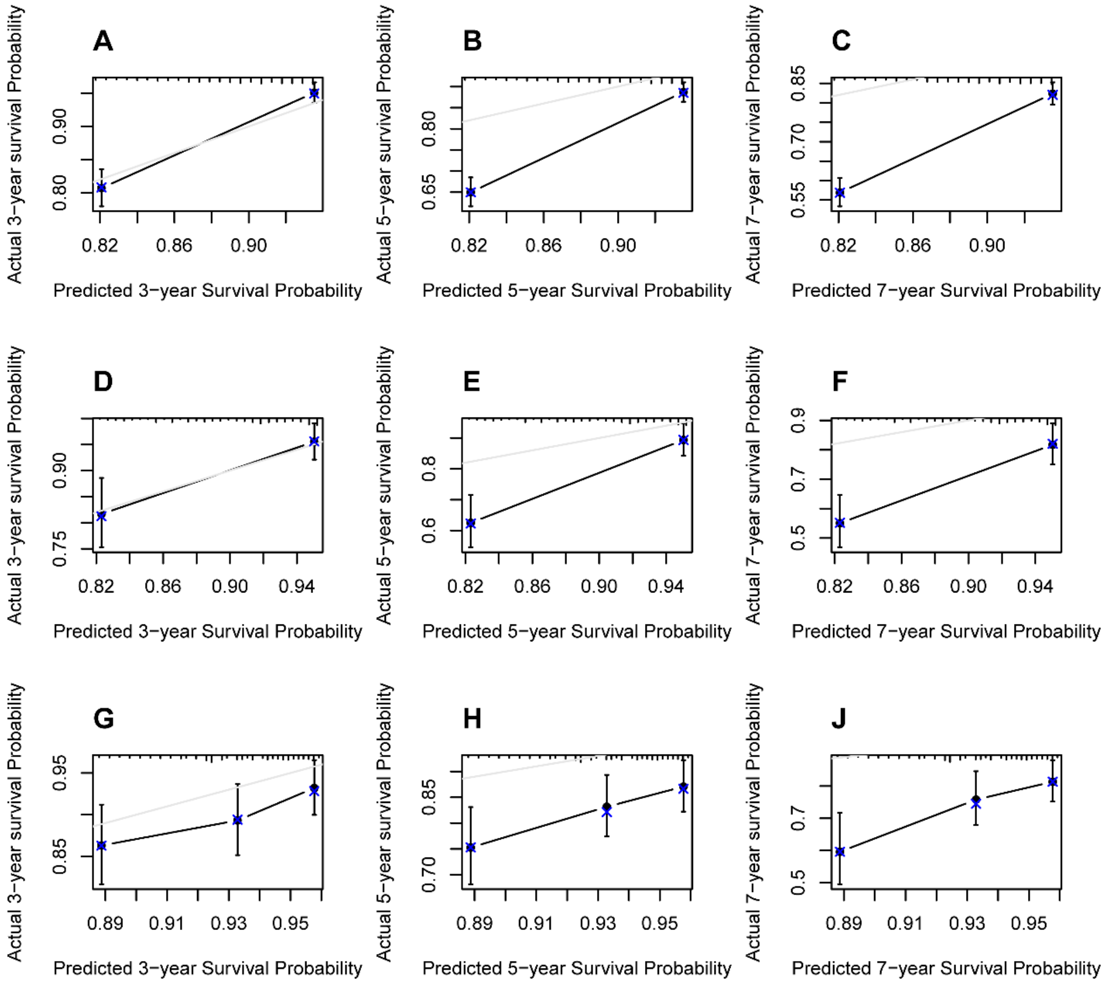


**Supplementary figure 6.** Internally and externally calibration of the nomogram in the training set (A-C), test set (D-F) and validation set (G-J).

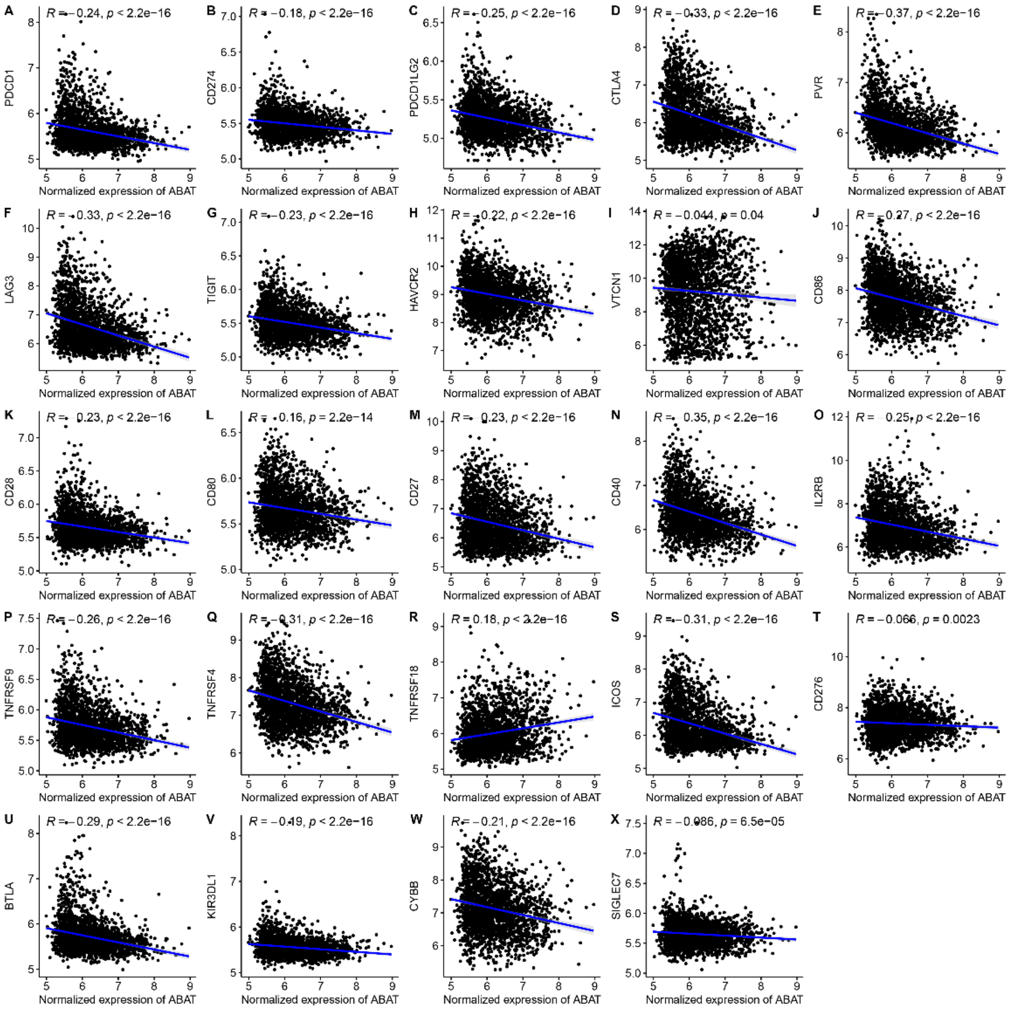


**Supplementary figure 7.** The correlation between the expression of ABAT and well-known immune checkpoint molecules. The correlation analysis was performed using Spearman's rank correlation.

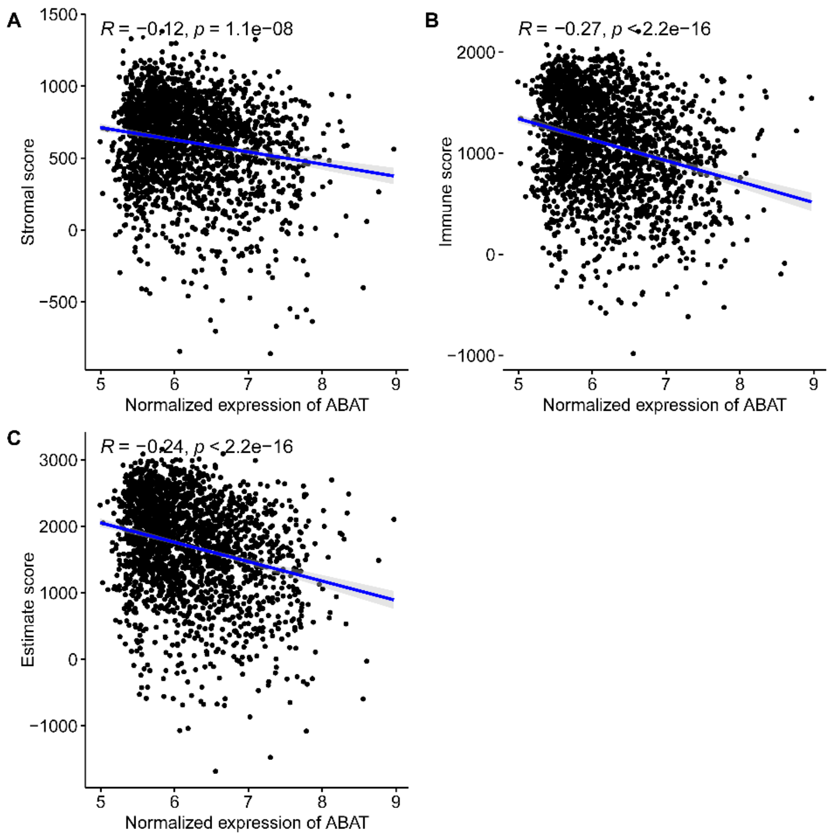


**Supplementary figure 8.** The correlation between the expression of ABAT and (A) stromal score, (B) immune score and (C) estimate score calculated using R packages ESTIMATE. The correlation analysis was performed using Spearman's rank correlation.

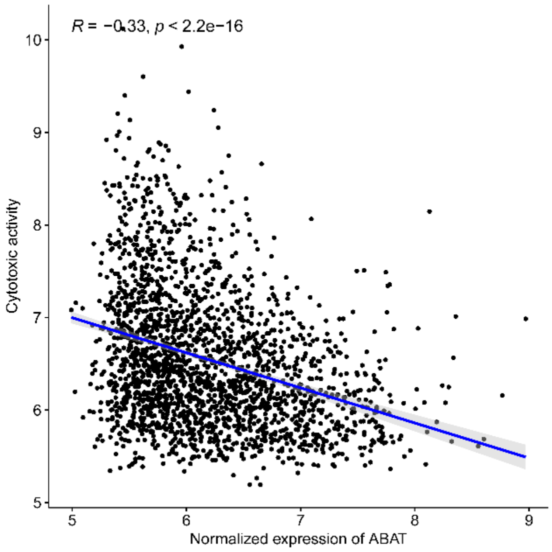


**Supplementary figure 9.** The correlation between the expression of ABAT and cytotoxic activity. The correlation analysis was performed using Spearman's rank correlation.

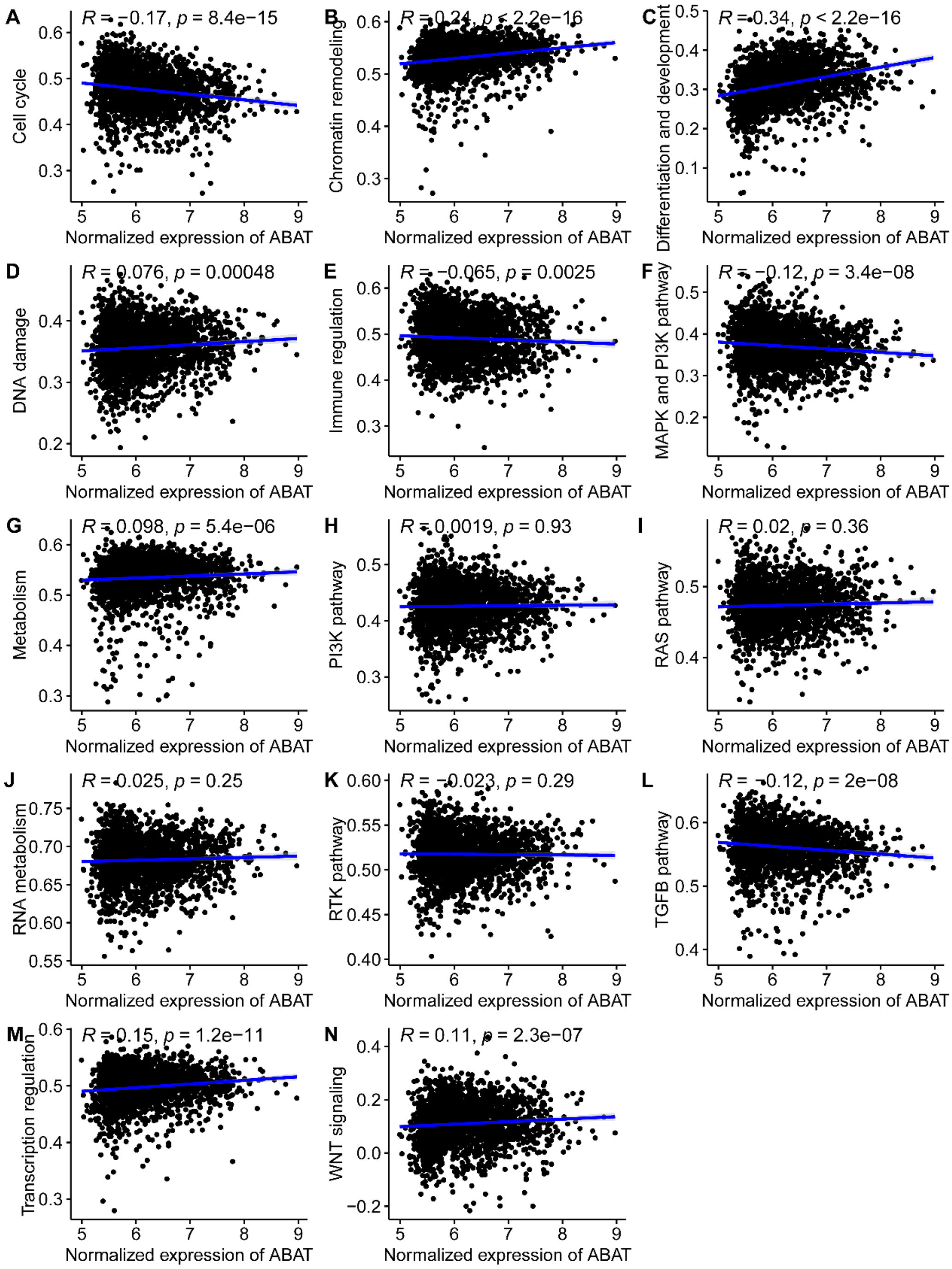


**Supplementary figure 10.** The correlation between the expression of ABAT and well-known cancer related pathway. The correlation analysis was performed using Spearman's rank correlation.

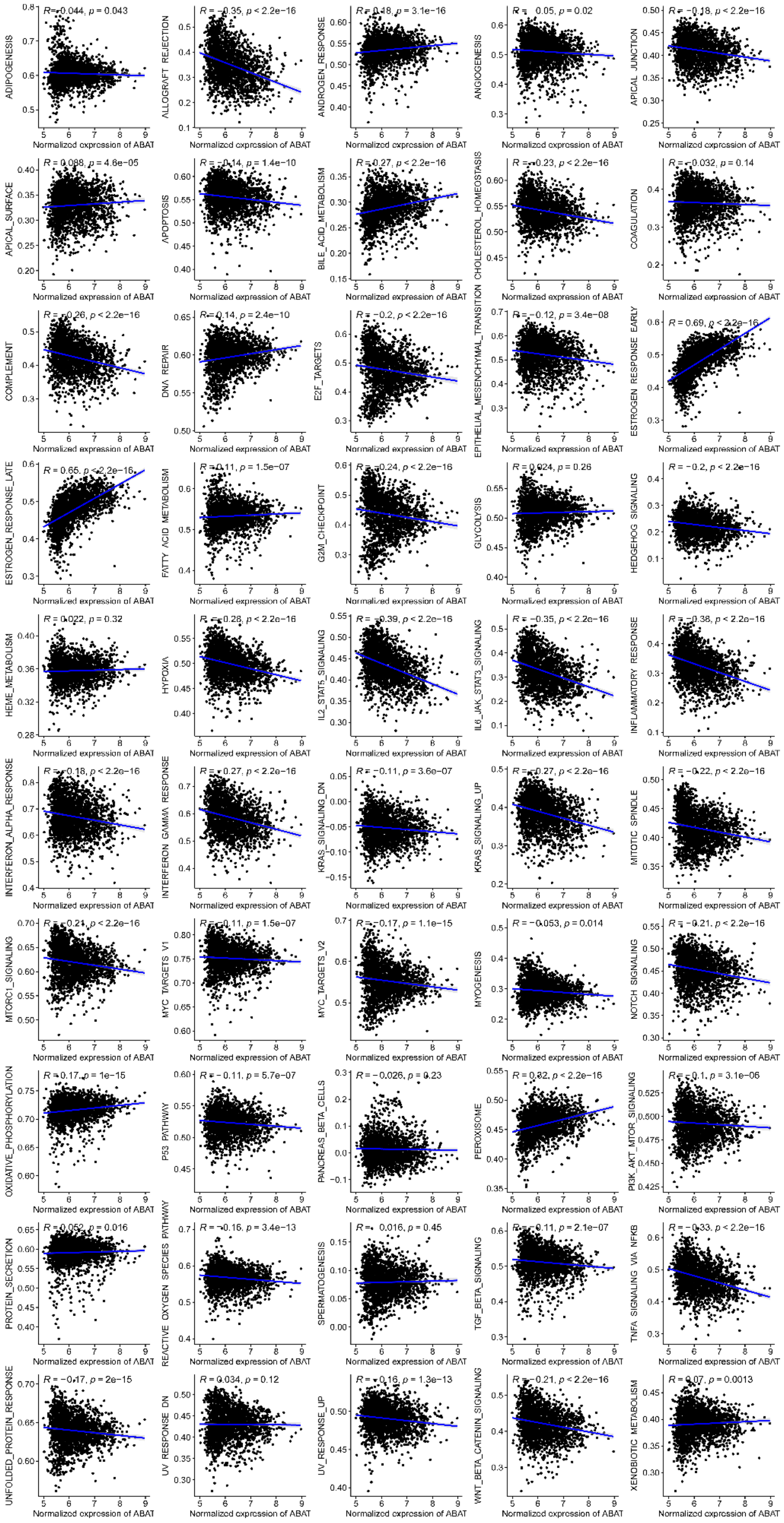


**Supplementary figure 11.** The correlation between the expression of ABAT and Hallmarks signature. The correlation analysis was performed using Spearman's rank correlation.

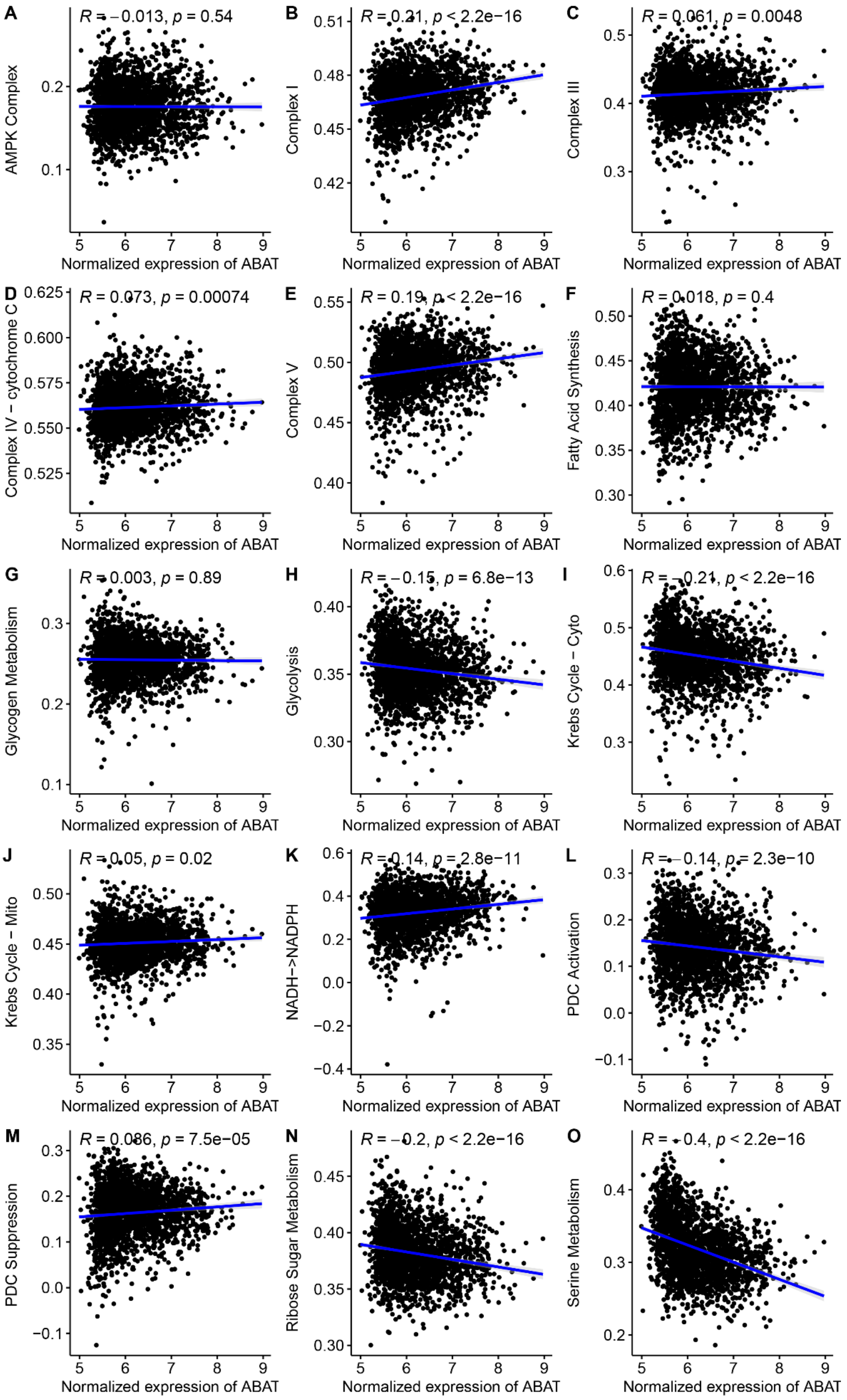


**Supplementary figure 12.** The correlation between the expression of ABAT and metabolism pathway. The correlation analysis was performed using Spearman's rank correlation.

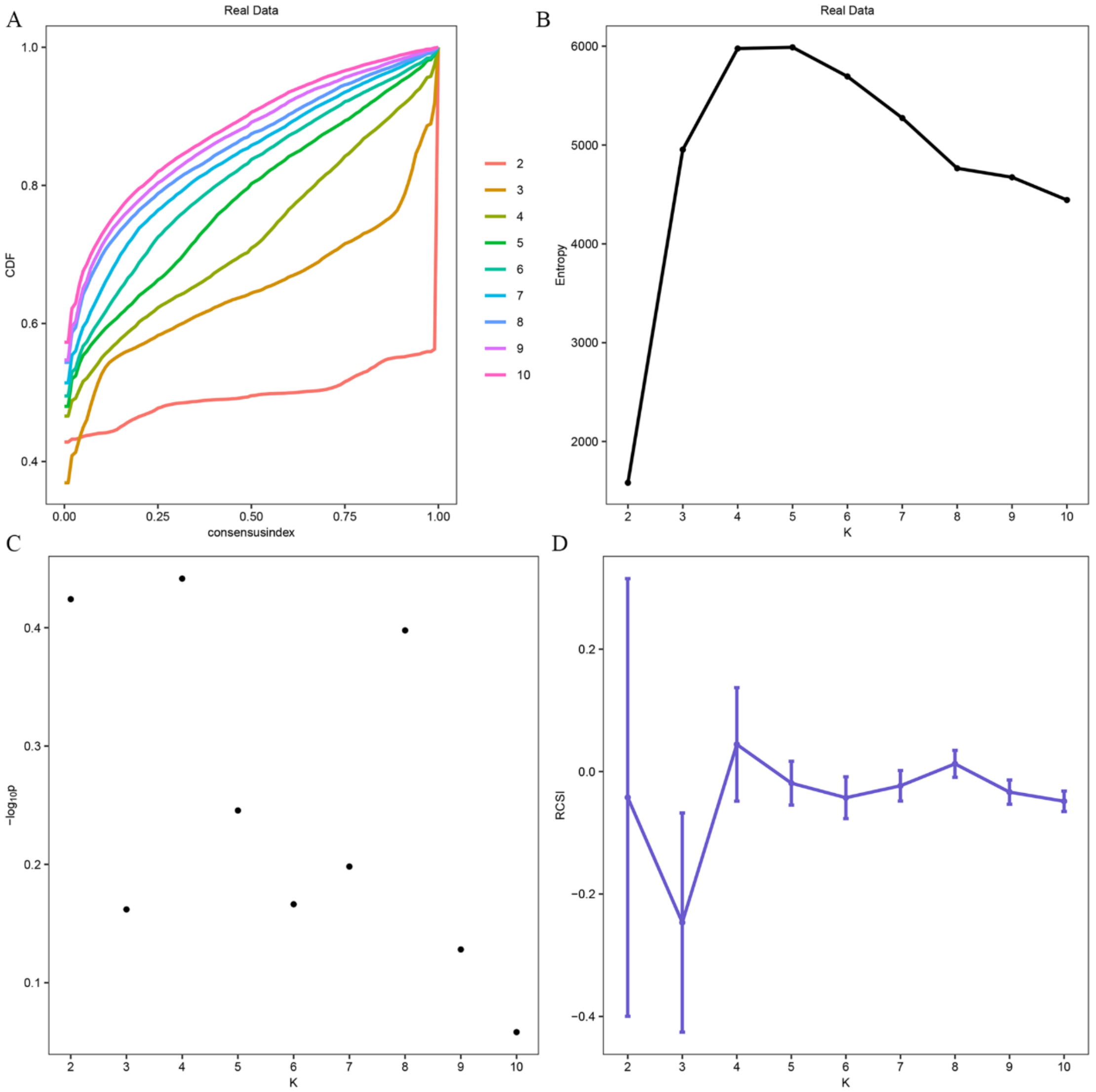


**Supplementary figure 13. Useful indexes for the determination of best K of clusters.** (A) CDF; (B) Entropy; (C) p values from the beta distribution; (D) Relative Cluster Stability Index (RCSI).

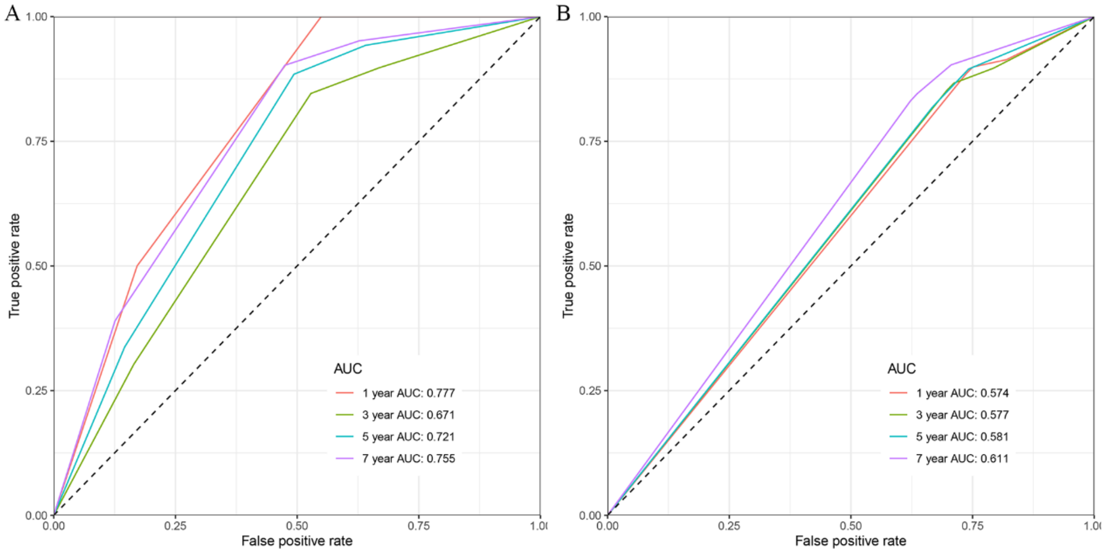


**Supplementary figure 14.** Prediction performance of the cancer subtypes in the training set (A)

and validation set (B).

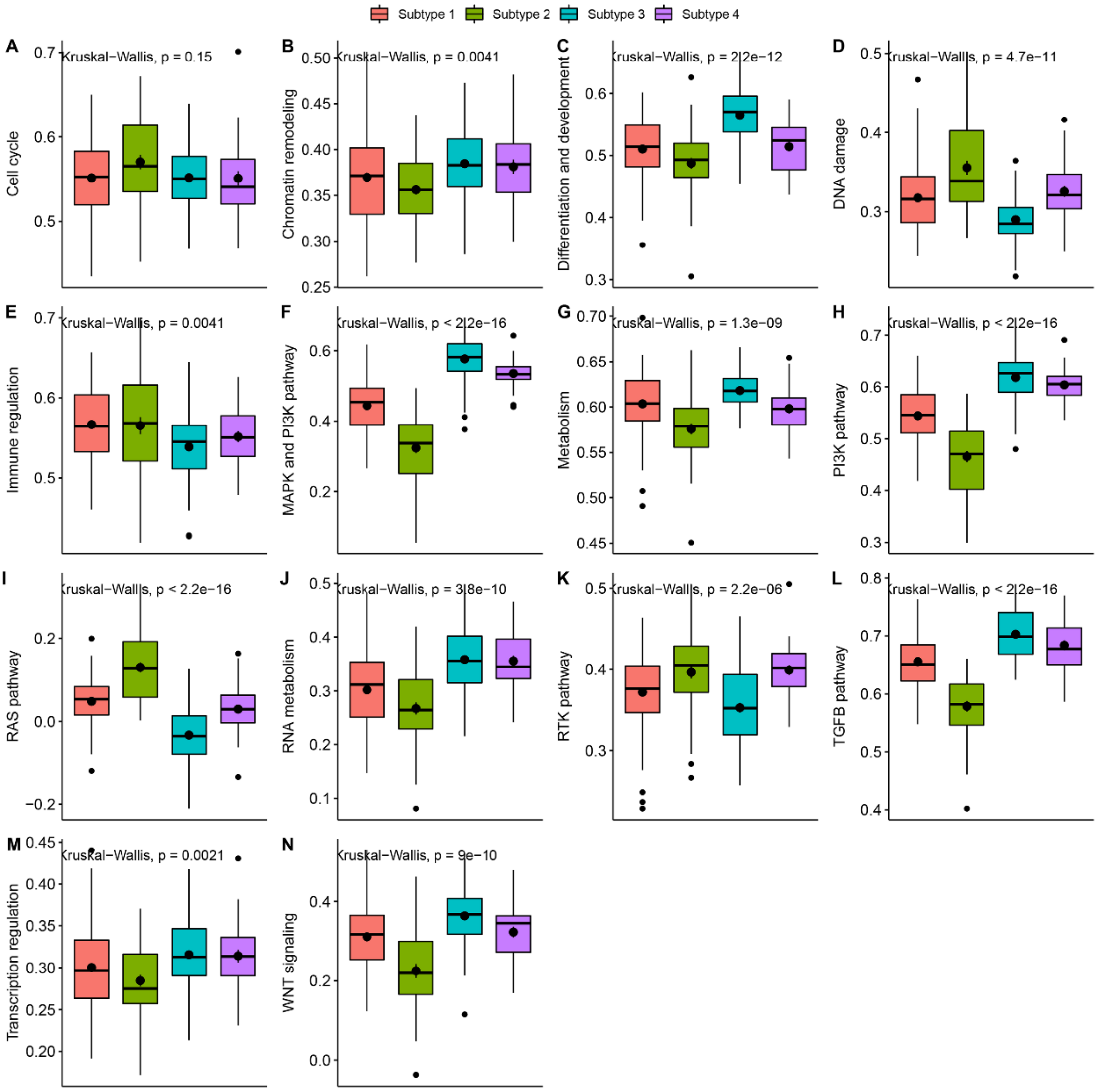


**Supplementary figure 15.** Comparison of common cancer related pathway score among the cancer subtypes in the training set.

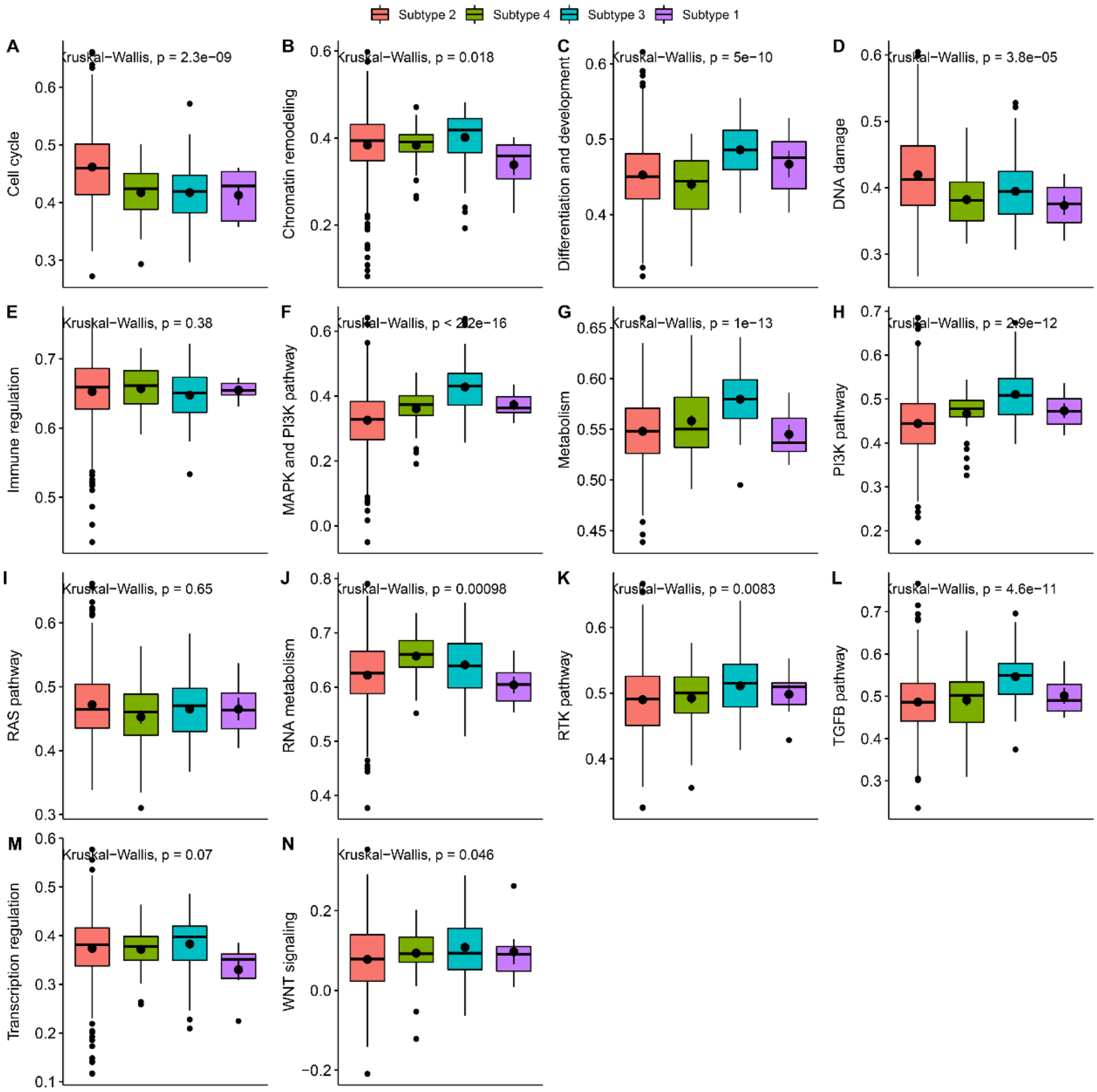


**Supplementary figure 16.** Comparison of common cancer related pathway score among the cancer subtypes in the validation set.

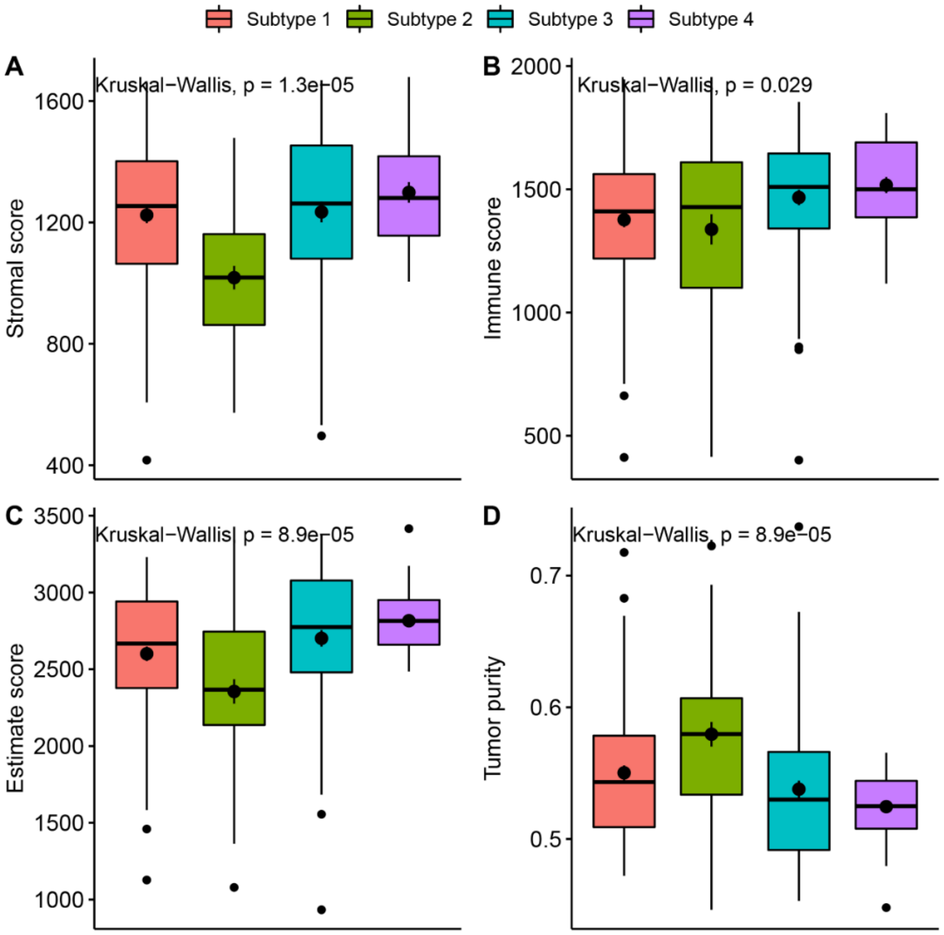


**Supplementary figure 17.** Comparison of the estimation of stromal and immune cells (ESTIMATE score) among the cancer subtypes in the training set.

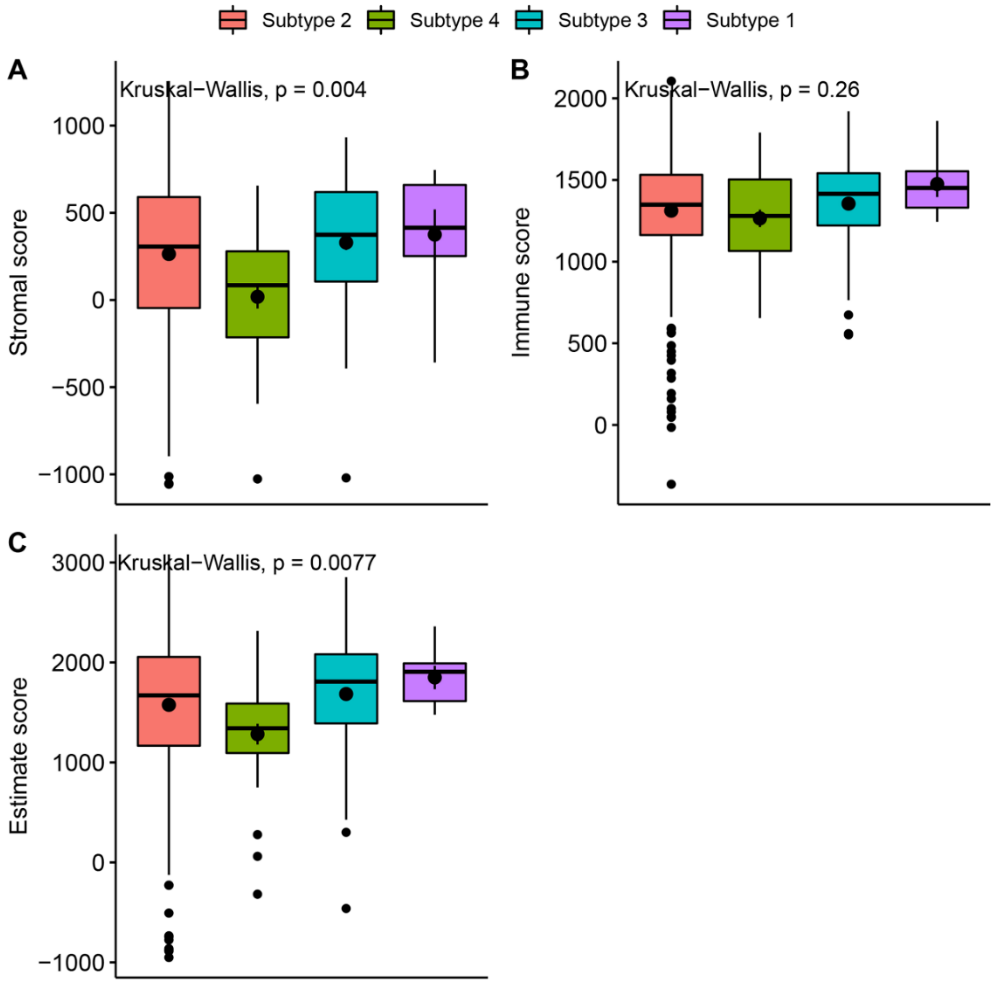


**Supplementary figure 18.** Comparison of the estimation of stromal and immune cells (ESTIMATE score) among the cancer subtypes in the validation set.

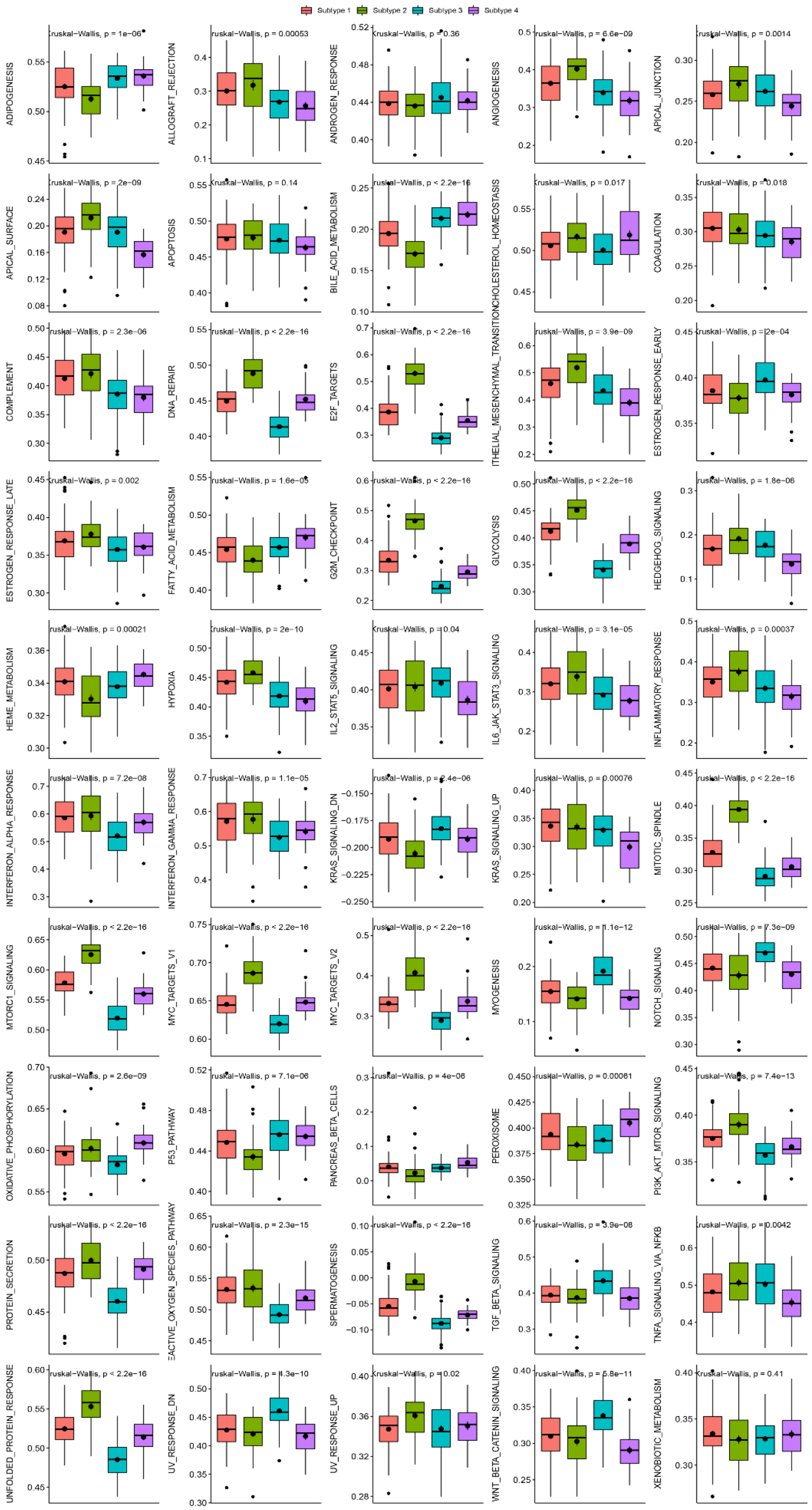


**Supplementary figure 19.** Comparison of hallmark signature score among the cancer subtypes in the training set.

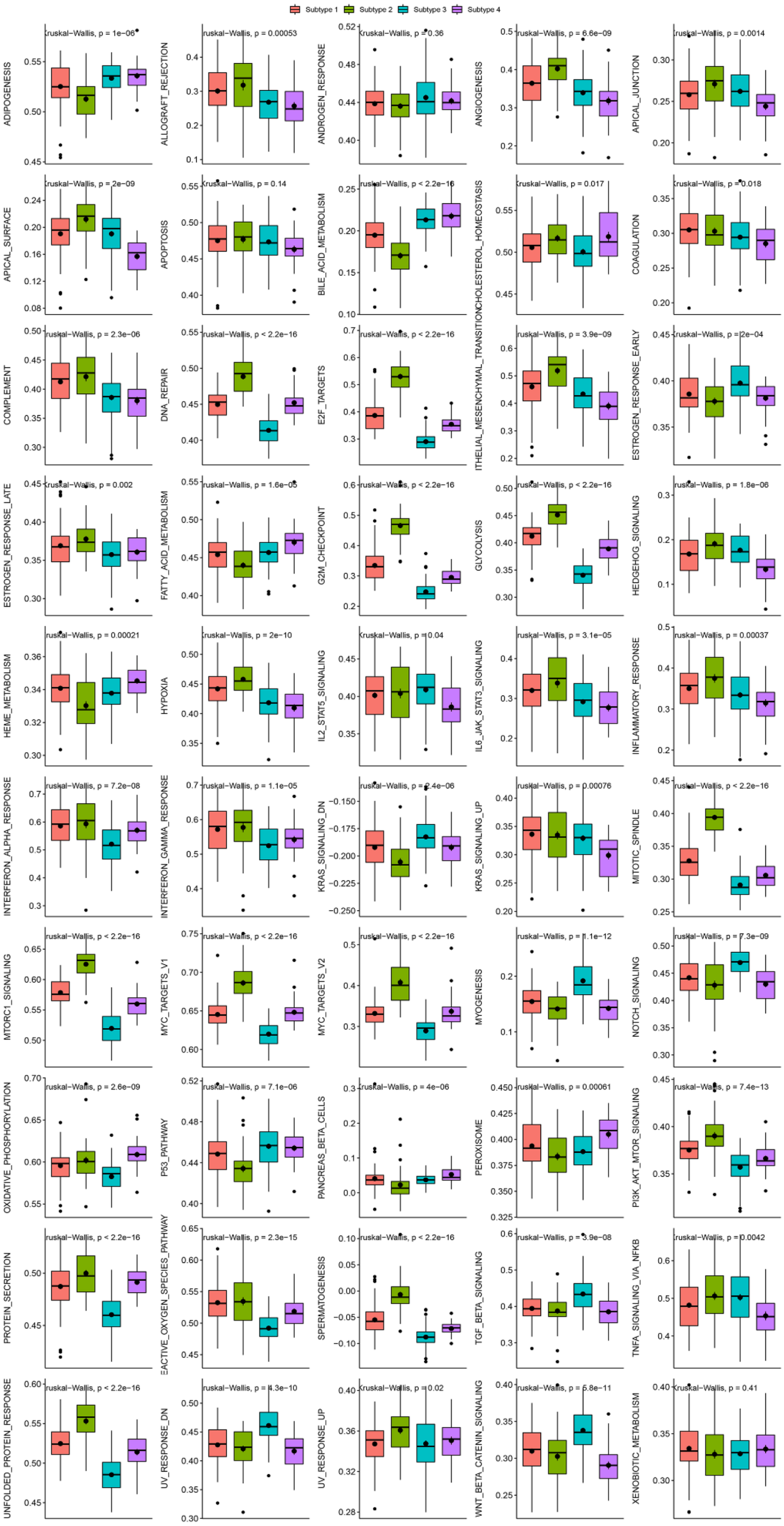


**Supplementary figure 20.** Comparison of hallmark signature score among the cancer subtypes in the validation set.

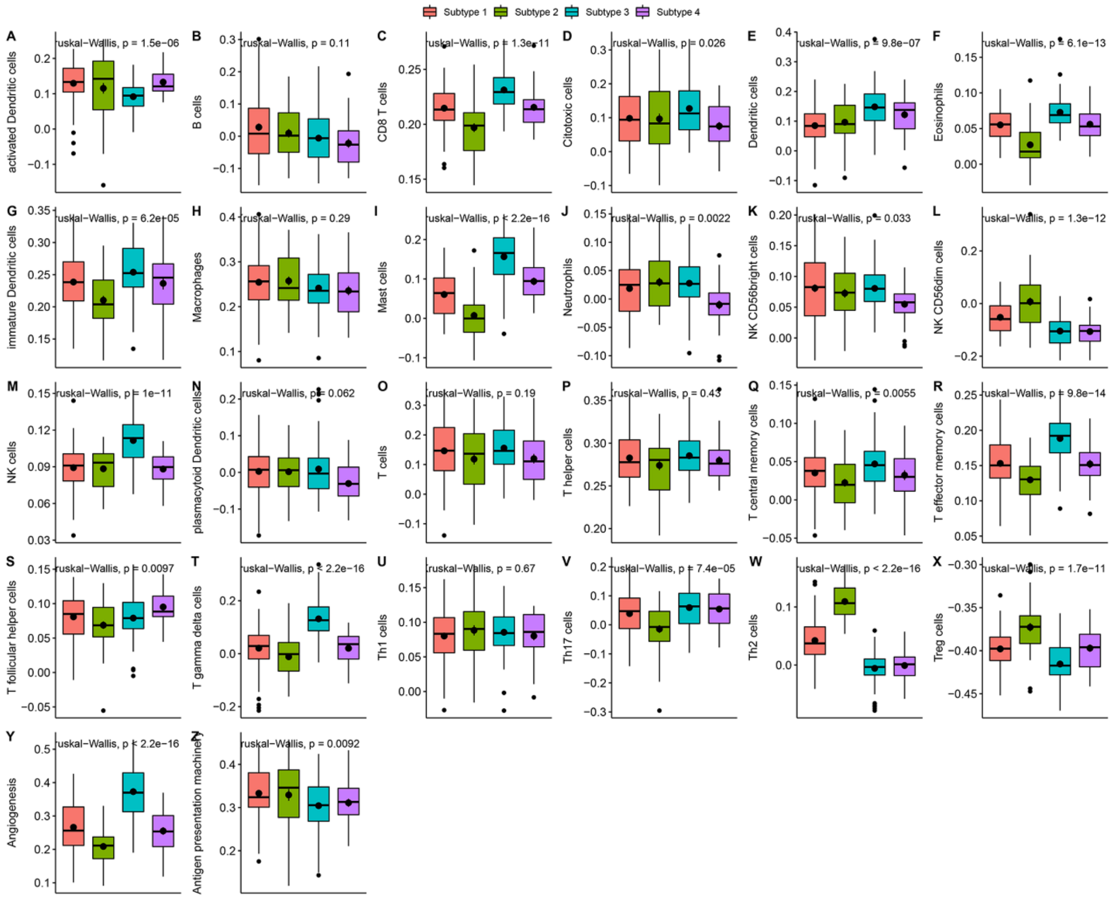

**Supplementary figure 21.** Comparison of immune cell infiltrations among the cancer subtypes in the training set.

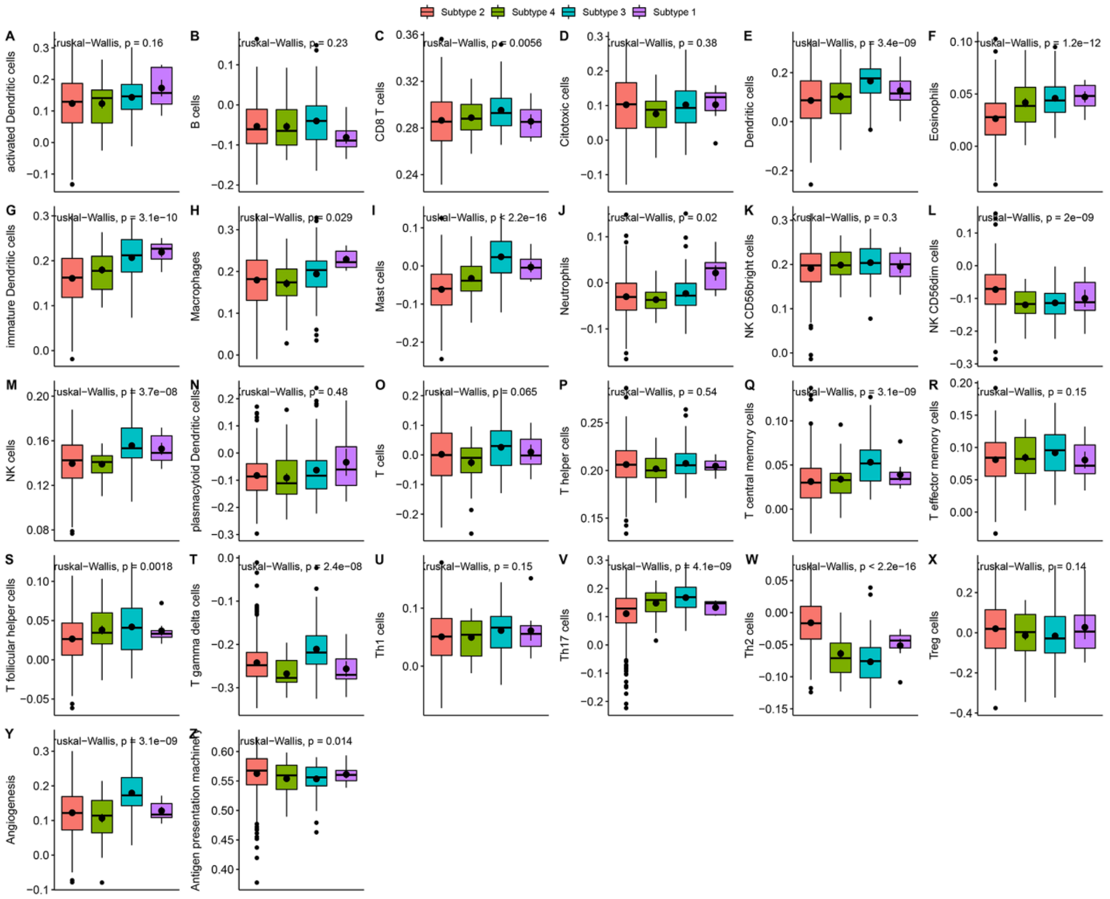


**Supplementary figure 22**. Comparison of immune cell infiltrations among the cancer subtypes in the validation set.

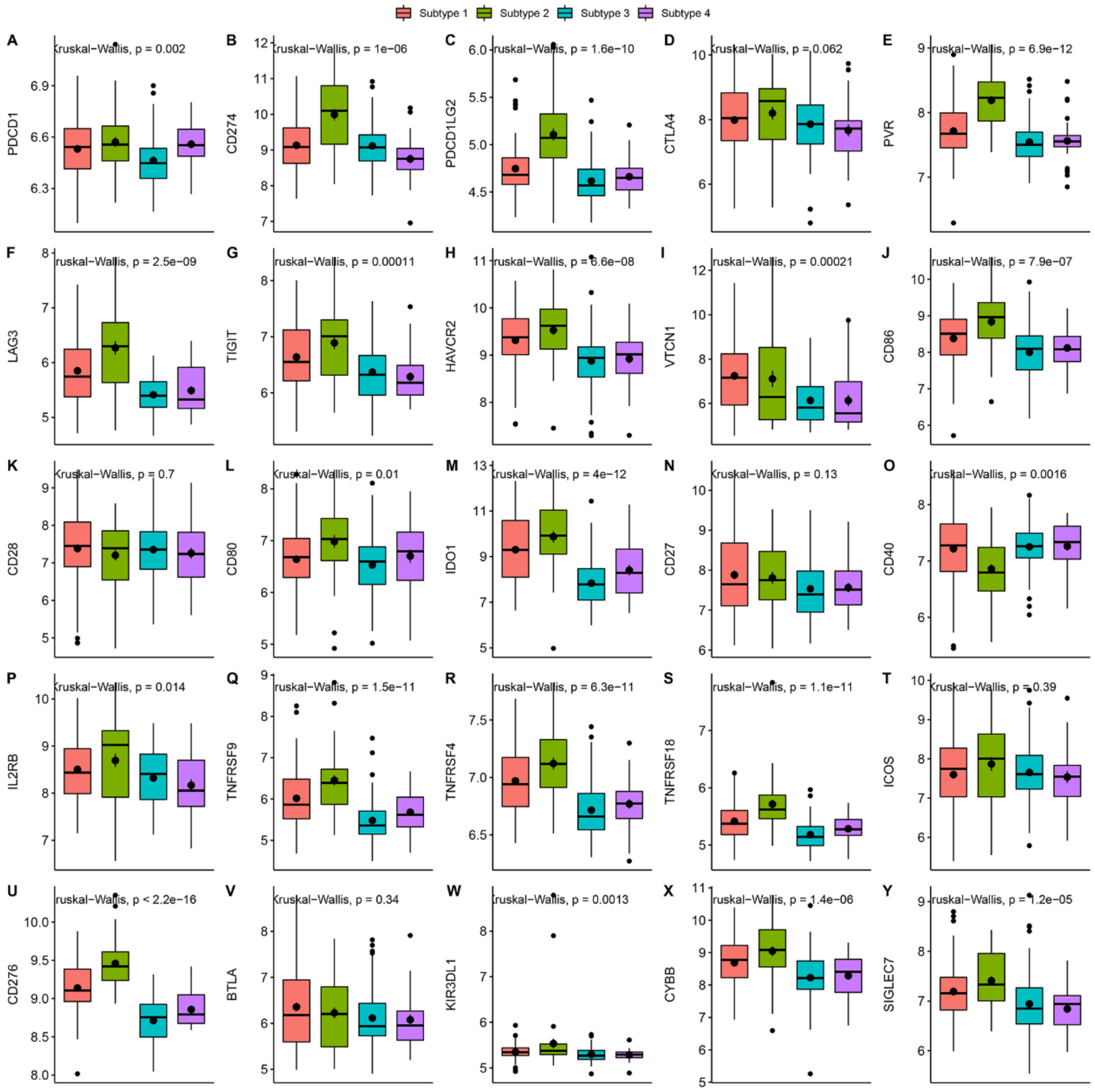


**Supplementary figure 23.** Comparison of the expression of immune checkpoint molecules among the cancer subtypes in the training set.

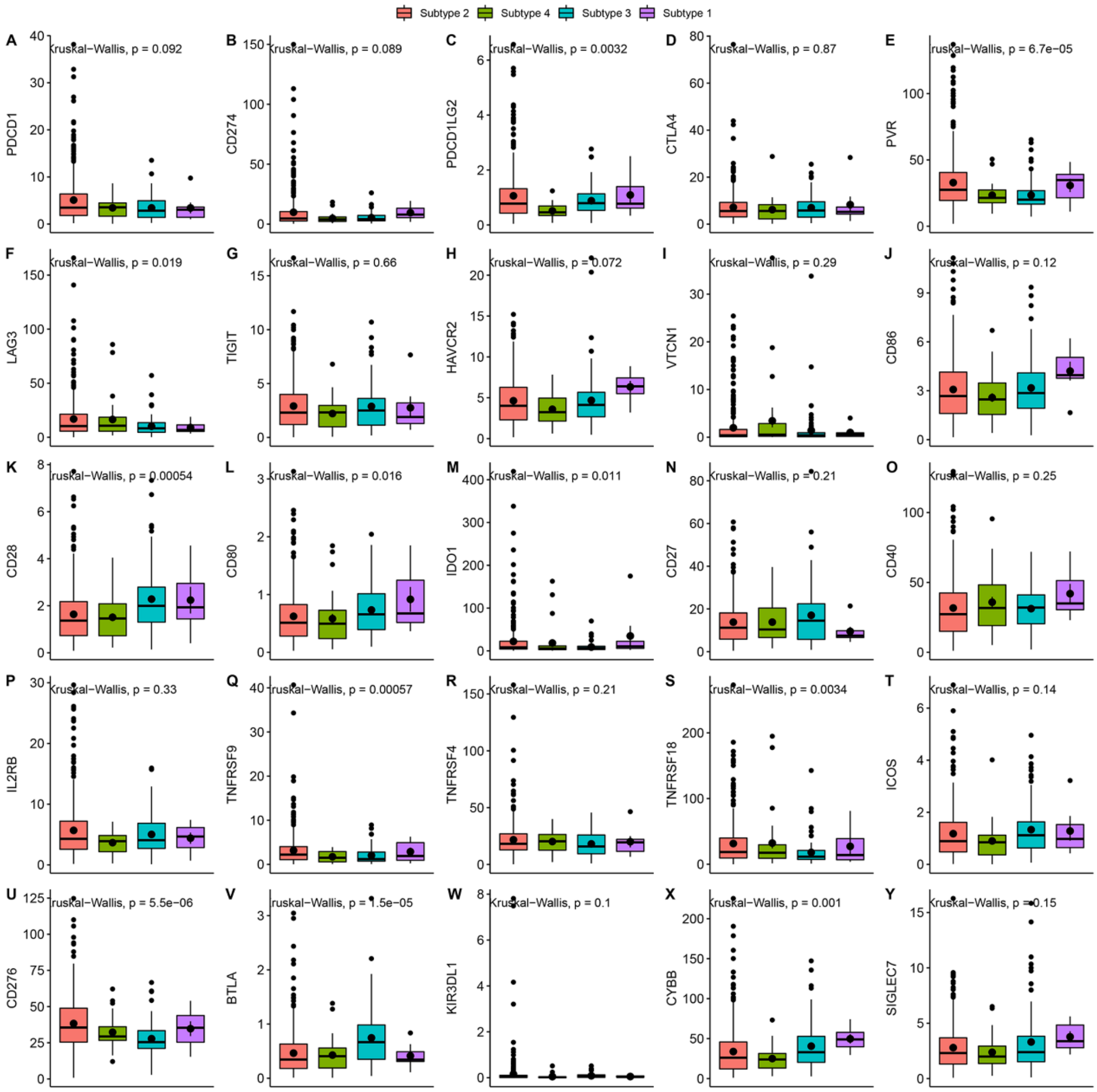


**Supplementary figure 24.** Comparison of the expression of immune checkpoint molecules among the cancer subtypes in the validation set.

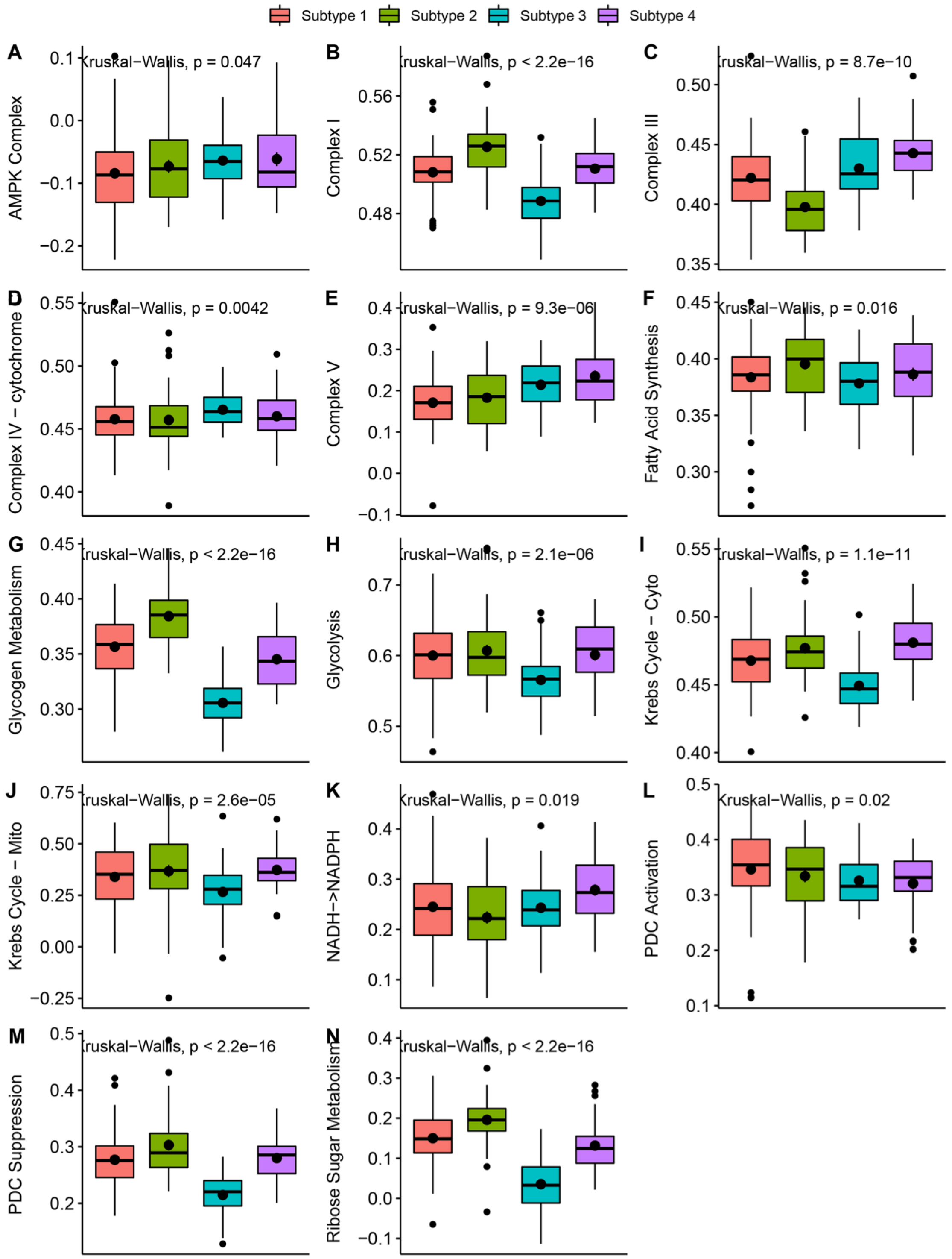


**Supplementary figure 25.** Comparison of the metabolic related signaling pathways among the cancer subtypes in the training set.

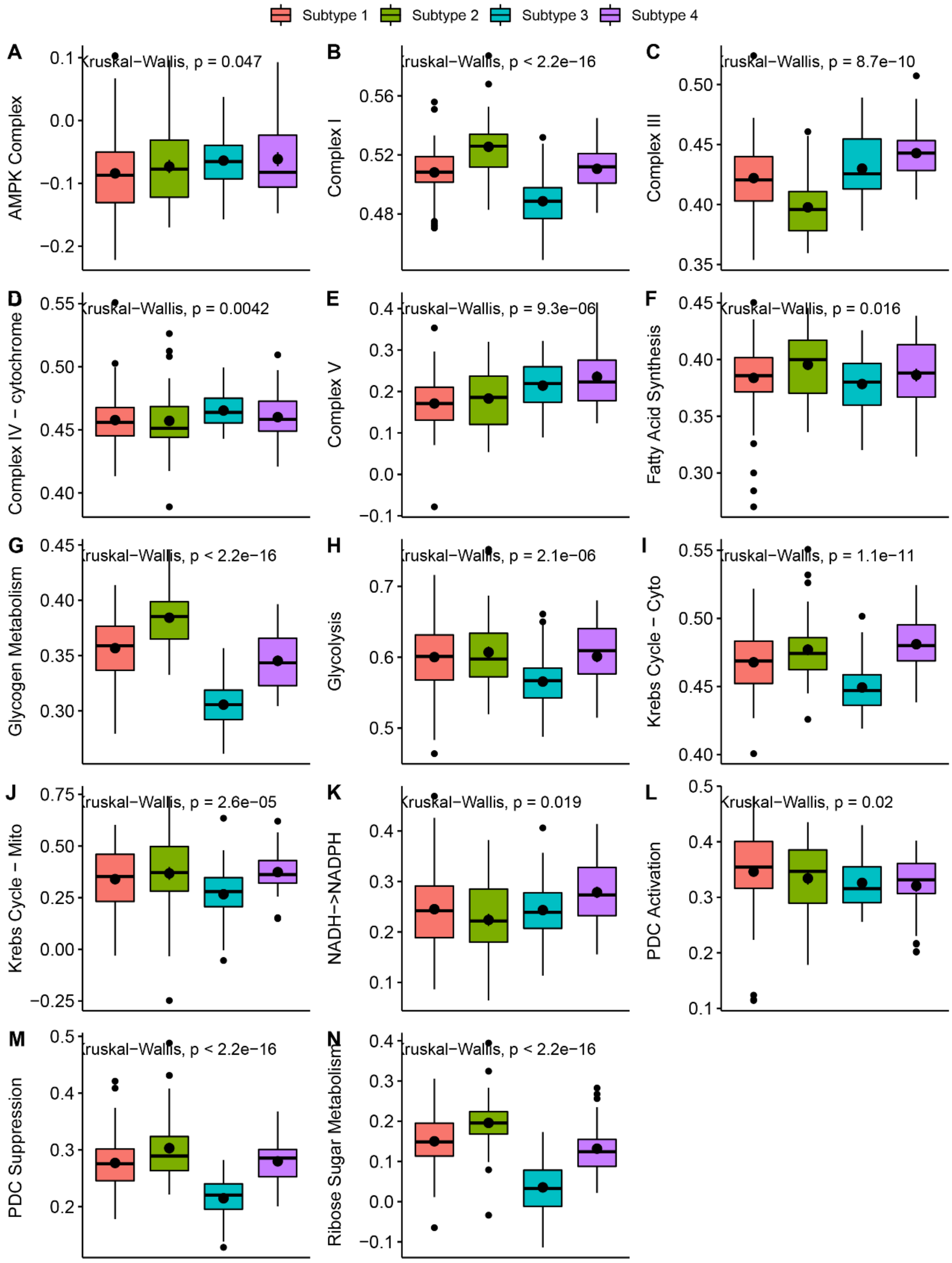


**Supplementary figure 26.** Comparison of the metabolic related signaling pathways among the cancer subtypes in the validation set.

| Study | logHR | SE(logHR) | Hazard Ratio | HR | 95%−CI | Weight |
|---|---|---|---|---|---|---|
| GSE3143 | −0.0021 | 0.0011 | | 1.00 | [1.00; 1.00] | 6.6% |
| GSE10886_GPL1390 | −0.0788 | 0.1491 | | 0.92 | [0.69; 1.24] | 3.7% |
| GSE10886_GPL887 | −61.7826 | Inf | | 0.00 | [0.00; Inf] | 0.0% |
| GSE18229_GPL1390 | −0.4483 | 0.1360 | | 0.64 | [0.49; 0.83] | 4.0% |
| GSE18229_GPL887 | −0.4871 | 0.2166 | | 0.61 | [0.40; 0.94] | 2.5% |
| GSE22226_GPL1708 | 0.0741 | 0.0795 | | 1.08 | [0.92; 1.26] | 5.4% |
| GSE22226_GPL4133 | −0.0614 | 0.1734 | | 0.94 | [0.67; 1.32] | 3.2% |
| GSE2607_GPL1390 | −0.3829 | 0.2203 | | 0.68 | [0.44; 1.05] | 2.5% |
| GSE2607_GPL887 | −1.0217 | 0.4219 | | 0.36 | [0.16; 0.82] | 0.9% |
| GSE6130_GPL1390 | −0.1944 | 0.1691 | | 0.82 | [0.59; 1.15] | 3.3% |
| GSE6130_GPL887 | −0.8702 | 0.4074 | | 0.42 | [0.19; 0.93] | 1.0% |
| Caldas_2007 | −0.7618 | 0.3538 | | 0.47 | [0.23; 0.93] | 1.2% |
| GSE12071 | −0.0334 | 0.1998 | | 0.97 | [0.65; 1.43] | 2.8% |
| GSE10510 | −0.5463 | 0.3386 | | 0.58 | [0.30; 1.12] | 1.3% |
| GSE159956 | −2.5370 | 0.6718 | | 0.08 | [0.02; 0.30] | 0.4% |
| GSE22133_GPL5345 | −0.2056 | 0.0712 | | 0.81 | [0.71; 0.94] | 5.6% |
| E_TABM_158 | 0.1877 | 0.1280 | | 1.21 | [0.94; 1.55] | 4.2% |
| GSE1456_GPL96 | −0.0956 | 0.1041 | | 0.91 | [0.74; 1.11] | 4.8% |
| GSE16446 | −0.4048 | 0.2804 | | 0.67 | [0.39; 1.16] | 1.8% |
| GSE20711 | −0.1145 | 0.1071 | | 0.89 | [0.72; 1.10] | 4.7% |
| GSE37751 | −0.0912 | 0.2817 | | 0.91 | [0.53; 1.59] | 1.8% |
| GSE42568 | −0.3968 | 0.0943 | | 0.67 | [0.56; 0.81] | 5.0% |
| GSE48390 | −0.2096 | 0.1998 | | 0.81 | [0.55; 1.20] | 2.8% |
| GSE58812 | −0.0865 | 0.1875 | | 0.92 | [0.64; 1.32] | 3.0% |
| GSE7390 | −0.1465 | 0.0937 | | 0.86 | [0.72; 1.04] | 5.1% |
| ICGC_BRCA_FR | 0.1511 | 0.6215 | | 1.16 | [0.34; 3.93] | 0.5% |
| ICGC_BRCA_KR | −0.0006 | 0.0004 | | 1.00 | [1.00; 1.00] | 6.6% |
| TCGA_BRCA | −0.0108 | 0.0091 | | 0.99 | [0.97; 1.01] | 6.6% |
| METABRIC | −0.2982 | 0.0550 | | 0.74 | [0.67; 0.83] | 6.0% |
| NKI | −1.9124 | 0.4413 | | 0.15 | [0.06; 0.35] | 0.9% |
| UCSF | −0.0345 | 0.2595 | | 0.97 | [0.58; 1.61] | 2.0% |
| **Random effects model** | | | | **0.83** | **[0.76; 0.90]** | **100.0%** |

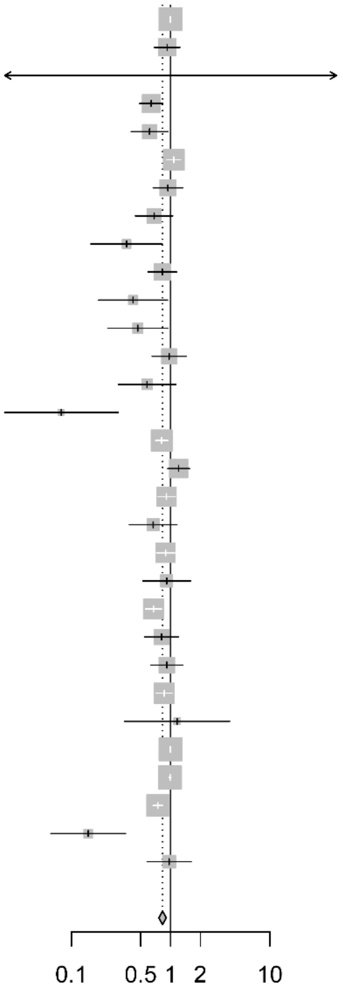


Heterogeneity: $I^2$ = 79%, $\tau^2$ = 0.0291, $p$ < 0.01

**Supplementary figure 27.** Meta-analysis of the prognosis role of ABAT in breast cancer.

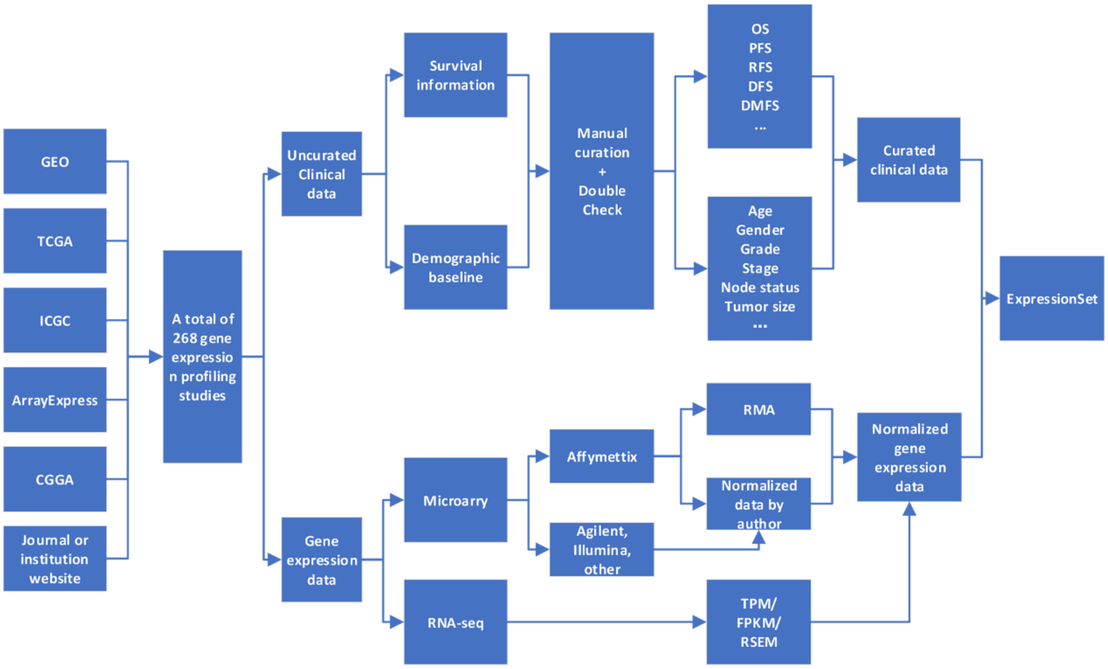

**Supplementary figure 28.** Flow chart of curation of public gene expression studies integrated in CuratedCancerPrognosisData.

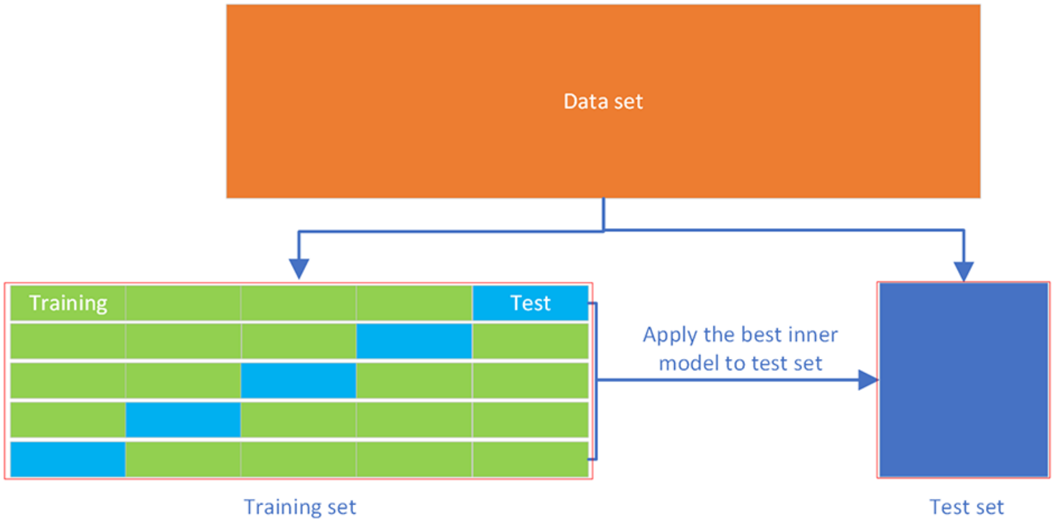


**Supplementary figure 29.** Workflow of cross validation.

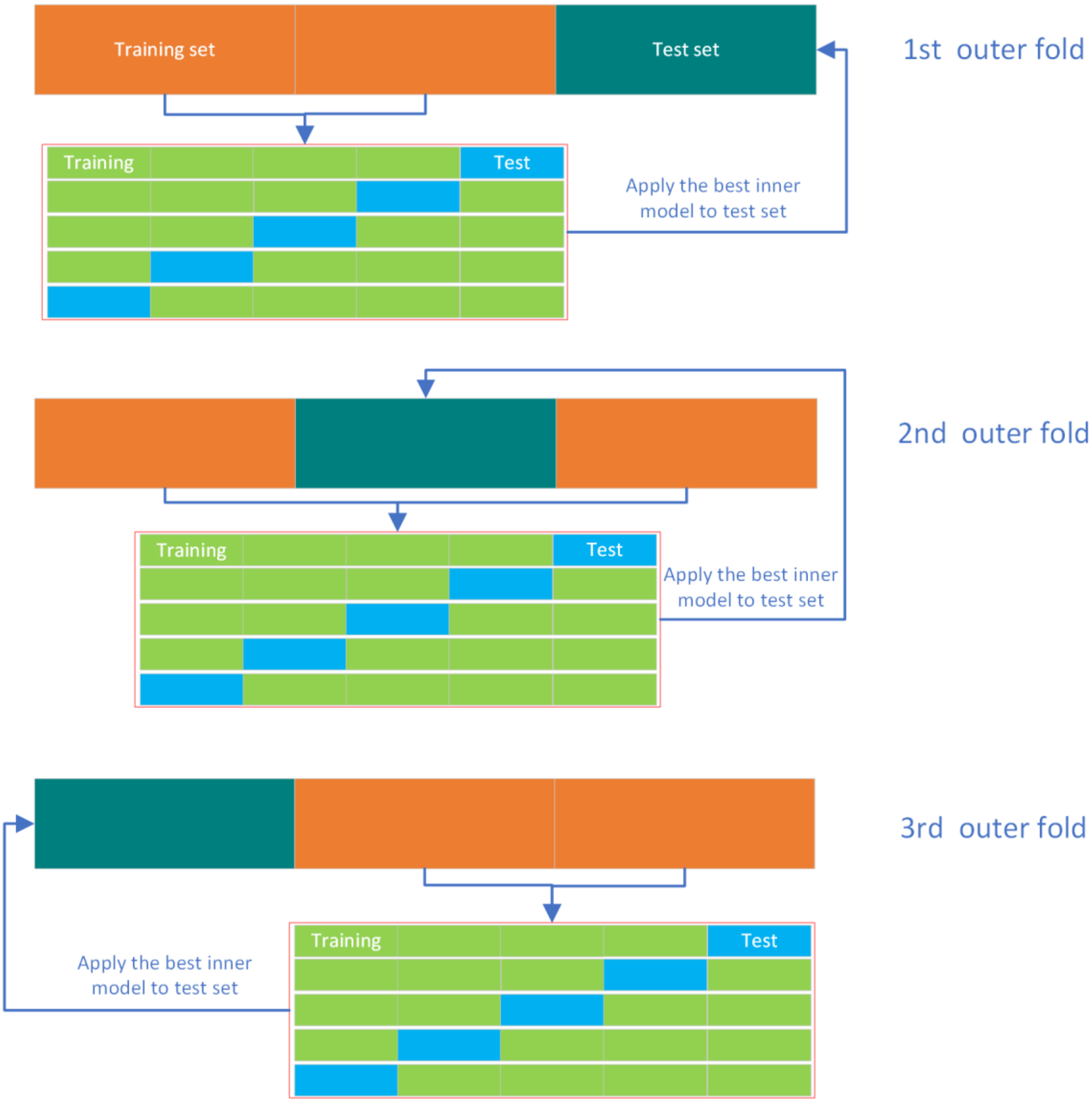


**Supplementary figure 30.** Workflow of nested cross validation.

**Supplementary table 1.** Public gene expression studies that included in the present study

| ID | Cancer | Dataset | Total genes | Total samples | Platform | Last update | Link | PMID |
|---|---|---|---|---|---|---|---|---|
| 1 | Acute lymphoblastic leukemia | TARGET_ALL_P3 | 58387 | 117 | Illumina HiSeq | 2019 | https://xenabrowser.net/datapages/ | 25207766 |
| 2 | Acute lymphoblastic leukemia | E_MTAB_1216 | 12547 | 101 | Affymetrix GeneChip Human Genome HG-U133A | 2014 | https://www.ebi.ac.uk/arrayexpress/experiments/E-MTAB-1216 | NA |
| 3 | Acute lymphoblastic leukemia | E_MTAB_1205 | 21653 | 38 | Affymetrix GeneChip Human Genome U133 Plus 2.0 | 2014 | https://www.ebi.ac.uk/arrayexpress/experiments/E-MTAB-1205 | 23436797 |
| 4 | Acute myeloid leukemia | TARGET_AML | 58387 | 145 | Illumina HiSeq | 2019 | https://xenabrowser.net/datapages/ | 26941285 |
| 5 | Acute myeloid leukemia | GSE12417_GPL570 | 21653 | 79 | Affymetrix Human Genome U133 Plus 2.0 Array | 2021 | https://www.ncbi.nlm.nih.gov/geo/query/acc.cgi?acc=GSE12417 | 18716133 |
| 6 | Acute myeloid leukemia | GSE12417_GPL96 | 12547 | 163 | Affymetrix Human Genome U133A Array | 2021 | https://www.ncbi.nlm.nih.gov/geo/query/acc.cgi?acc=GSE12417 | 18716133 |
| 7 | Acute myeloid leukemia | GSE12417_GPL97 | 10603 | 163 | Affymetrix Human Genome U133B Array | 2021 | https://www.ncbi.nlm.nih.gov/geo/query/acc.cgi?acc=GSE12417 | 18716133 |
| 8 | Acute myeloid leukemia | TCGA_LAML | 34849 | 117 | HTSeq | 2021 | https://portal.gdc.cancer.gov/projectsTCGA-LAML | 23634996 |
| 9 | Adrenocortical carcinoma | TCGA_ACC | 34849 | 79 | HTSeq | 2021 | https://portal.gdc.cancer.gov/projectsTCGA-ACC | 27165744 |

| | | | | | | | | |
|---|---|---|---|---|---|---|---|---|
| 10 | Adult soft tissue sarcomas | TCGA_SARC | 34849 | 255 | HTSeq | 2021 | https://portal.gdc.cancer.gov/projectsTCGA-SARC | 29100075 |
| 11 | Anaplastic large cell lymphomas | E_TABM_117 | 12547 | 37 | Affymetrix GeneChip Human Genome HG-U133A | 2014 | https://www.ebi.ac.uk/arrayexpress/experiments/E-TABM-117 | 17077326 |
| 12 | Bladder cancer | GSE13507 | 24357 | 256 | Illumina human-6 v2.0 expression beadchip | 2020 | https://www.ncbi.nlm.nih.gov/geo/query/acc.cgi?acc=GSE13507 | 20059769 |
| 13 | Bladder cancer | GSE5287 | 12178 | 30 | Affymetrix Human Genome U133A Array | 2018 | https://www.ncbi.nlm.nih.gov/geo/query/acc.cgi?acc=GSE5287 | 17671123 |
| 14 | Bladder cancer | GSE1827 | 6225 | 80 | JAKE | 2015 | https://www.ncbi.nlm.nih.gov/geo/query/acc.cgi?acc=GSE1827 | 15930339 |
| 15 | Bladder cancer | GSE19915_GPL3883 | 10314 | 84 | Swegene Human 27K RAP UniGene188 array | 2012 | https://www.ncbi.nlm.nih.gov/geo/query/acc.cgi?acc=GSE19915 | 20406976 |
| 16 | Bladder cancer | GSE19915_GPL5186 | 11671 | 98 | SWEGENE H_v3.0.1 35K | 2012 | https://www.ncbi.nlm.nih.gov/geo/query/acc.cgi?acc=GSE19915 | 20406976 |
| 17 | Bladder cancer | GSE32894 | 16940 | 308 | Illumina HumanHT-12 V3.0 expression beadchip | 2020 | https://www.ncbi.nlm.nih.gov/geo/query/acc.cgi?acc=GSE32894 | 22553347 |
| 18 | Bladder cancer | E_MTAB_4321 | 38267 | 476 | FPKM normalized RNAseq data | NA | https://www.ebi.ac.uk/arrayexpress/experiments/E-MTAB-4321 | 27321955 |
| 19 | Bladder cancer | GSE19423 | 24357 | 48 | Illumina human-6 v2.0 expression beadchip | 2013 | https://www.ncbi.nlm.nih.gov/geo/query/acc.cgi?acc=GSE19423 | 20233890 |
| 20 | Bladder cancer | GSE48276 | 20818 | 116 | Illumina HumanHT-12 WG-DASL V4.0 R2 expression beadchip | 2017 | https://www.ncbi.nlm.nih.gov/geo/query/acc.cgi?acc=GSE48276 | 24525232 |

| | | | | | | | | |
|---|---|---|---|---|---|---|---|---|
| 21 | Bladder cancer | GSE19750 | 21653 | 48 | Affymetrix Human Genome U133 Plus 2.0 Array | 2019 | https://www.ncbi.nlm.nih.gov/geo/query/acc.cgi?acc=GSE19750 | 24238056 |
| 22 | Bladder cancer | GSE31684 | 21653 | 93 | Affymetrix Human Genome U133 Plus 2.0 Array | 2019 | https://www.ncbi.nlm.nih.gov/geo/query/acc.cgi?acc=GSE31684 | 22228636 |
| 23 | Bladder cancer | TCGA_BLCA | 34849 | 400 | HTSeq | 2021 | https://portal.gdc.cancer.gov/projectsTCGA-BLCA | 24476821 |
| 24 | Bone cancer-Ewing sarcoma | ICGC_BOCA_FR | 24818 | 57 | HTSeq | 2019 | https://dcc.icgc.org/releases/current/Projects/BOCA-FR | 25223734 |
| 25 | Brain tumor-GBM | E_MTAB_951 | 21653 | 23 | Affymetrix GeneChip Human Genome U133 Plus 2.0 | 2014 | https://www.ebi.ac.uk/arrayexpress/experiments/E-MTAB-951 | 24804210 |
| 26 | Brain tumor-GBM | GSE13041_GPL570 | 21653 | 27 | Affymetrix Human Genome U133 Plus 2.0 Array | 2019 | https://www.ncbi.nlm.nih.gov/geo/query/acc.cgi?acc=GSE13041 | 18940004 |
| 27 | Brain tumor-GBM | GSE13041_GPL8300 | 8619 | 49 | Affymetrix Human Genome U95 Version 2 Array | 2019 | https://www.ncbi.nlm.nih.gov/geo/query/acc.cgi?acc=GSE13041 | 18940004 |
| 28 | Brain tumor-GBM | GSE13041_GPL96 | 12547 | 191 | Affymetrix Human Genome U133A Array | 2019 | https://www.ncbi.nlm.nih.gov/geo/query/acc.cgi?acc=GSE13041 | 18940004 |
| 29 | Brain tumor-GBM | GSE42669 | 20202 | 58 | Affymetrix Human Gene 1.0 ST Array | 2018 | https://www.ncbi.nlm.nih.gov/geo/query/acc.cgi?acc=GSE42669 | 23333277 |
| 30 | Brain tumor-GBM | GSE7696 | 21653 | 84 | Affymetrix Human Genome U133 Plus 2.0 Array | 2019 | https://www.ncbi.nlm.nih.gov/geo/query/acc.cgi?acc=GSE7696 | 18565887 |

| | | | | | | | | |
|---|---|---|---|---|---|---|---|---|
| 31 | Brain tumor-GBM | TCGA_GBM | 34849 | 143 | HTSeq | 2021 | https://portal.gdc.cancer.gov/projectsTCGA-GBM | 18772890 |
| 32 | Brain tumor-Glioma | E_MTAB_2768 | 21653 | 71 | Affymetrix GeneChip Human Genome U133 Plus 2.1 | 2018 | https://www.ebi.ac.uk/arrayexpress/experiments/E-MTAB-2768 | 26068201 |
| 33 | Brain tumor-Glioma | GSE2817 | 21653 | 25 | Affymetrix Human Genome U133 Plus 2.0 Array | 2019 | https://www.ncbi.nlm.nih.gov/geo/query/acc.cgi?acc=GSE2817 | 17140431 |
| 34 | Brain tumor-Glioma | TCGA_LGG | 34849 | 495 | HTSeq | 2021 | https://portal.gdc.cancer.gov/projectsTCGA-LGG | 26061751 |
| 35 | Brain tumor-Glioma/GBM | GSE108474 | 23519 | 541 | Affymetrix Human Genome U133 Plus 2.0 Array | 2019 | https://www.ncbi.nlm.nih.gov/geo/query/acc.cgi?acc=GSE108474 | 30106394 |
| 36 | Brain tumor-Glioma/GBM | CGGA_301 | 19416 | 300 | Agilent Whole Human Genome (Array) | 2021 | http://www.cgga.org.cn/download.jsp | 27564467 |
| 37 | Brain tumor-Glioma/GBM | E_MTAB_3892 | 21653 | 175 | Affymetrix GeneChip Human Genome U133 Plus 2.0 | 2018 | https://www.ebi.ac.uk/arrayexpress/experiments/E-MTAB-3892 | 27090007 |
| 38 | Brain tumor-Glioma/GBM | GSE16011 | 21653 | 284 | Affymetrix GeneChip Human Genome U133 Plus 2.0 Array | 2014 | https://www.ncbi.nlm.nih.gov/geo/query/acc.cgi?acc=GSE16011 | 19920198 |
| 39 | Brain tumor-Glioma/GBM | GSE4271_GPL96 | 12547 | 100 | Affymetrix Human Genome U133B Array | 2019 | https://www.ncbi.nlm.nih.gov/geo/query/acc.cgi?acc=GSE4271 | 16530701 |
| 40 | Brain tumor-Glioma/GBM | GSE4271_GPL97 | 10603 | 100 | Affymetrix Human Genome U133B Array | 2019 | https://www.ncbi.nlm.nih.gov/geo/query/acc.cgi?acc=GSE4271 | 16530701 |

| | | | | | | | | |
|---|---|---|---|---|---|---|---|---|
| 41 | Brain tumor-Glioma/GBM | GSE4412_GPL96 | 12547 | 85 | Affymetrix Human Genome U134A Array | 2018 | https://www.ncbi.nlm.nih.gov/geo/query/acc.cgi?acc=GSE4412 | 15374961 |
| 42 | Brain tumor-Glioma/GBM | GSE4412_GPL97 | 10603 | 85 | Affymetrix Human Genome U134B Array | 2018 | https://www.ncbi.nlm.nih.gov/geo/query/acc.cgi?acc=GSE4412 | 15374961 |
| 43 | Brain tumor-Glioma/GBM | CGGA_325 | 24326 | 325 | Illumina HiSeq 2000 or 2500 | 2021 | http://www.cgga.org.cn/download.jsp | 28291232 |
| 44 | Brain tumor-Glioma/GBM | CGGA_693 | 23987 | 693 | Illumina HiSeq | 2021 | http://www.cgga.org.cn/download.jsp | 25031032 |
| 45 | Brain tumor-Medulloblastoma | GSE30074 | 20202 | 30 | Affymetrix Human Gene 1.0 ST Array | 2019 | https://www.ncbi.nlm.nih.gov/geo/query/acc.cgi?acc=GSE30074 | 22090452 |
| 46 | Brain tumor-Medulloblastoma | GSE37418 | 21653 | 76 | Affymetrix Human Genome U133 Plus 2.0 Array | 2019 | https://www.ncbi.nlm.nih.gov/geo/query/acc.cgi?acc=GSE37418 | 22722829 |
| 47 | Brain tumor-Meningioma | GSE16581 | 21653 | 68 | Affymetrix GeneChip Human Genome U133 Plus 2.1 Array | 2019 | https://www.ncbi.nlm.nih.gov/geo/query/acc.cgi?acc=GSE16581 | 20015288 |
| 48 | Breast cancer | E_MTAB_7201 | 34729 | 94 | Agilent-072363 SurePrint G3 Human GE v3 8x60K Microarray 039494 | 2018 | https://www.ebi.ac.uk/arrayexpress/experiments/E-MTAB-7201 | 28123884 |
| 49 | Breast cancer | GSE3143 | 8619 | 158 | Affymetrix HGU95 | 2018 | https://www.ncbi.nlm.nih.gov/geo/query/acc.cgi?acc=GSE3143 | 16273092 |
| 50 | Breast cancer | GSE10886_GPL1390 | 10040 | 197 | Agilent Human 1A Oligo UNC custom Microarrays | 2017 | https://www.ncbi.nlm.nih.gov/geo/query/acc.cgi?acc=GSE10886 | 19204204 |
| 51 | Breast cancer | GSE10886_GPL887 | 16546 | 27 | Agilent-012097 Human 1A Microarray (V2) G4110B | 2017 | https://www.ncbi.nlm.nih.gov/geo/query/acc.cgi?acc=GSE10886 | 19204204 |

| | | | | | | | | |
|---|---|---|---|---|---|---|---|---|
| 52 | Breast cancer | GSE18229_GPL1390 | 10040 | 199 | Agilent Human 1A Oligo UNC custom Microarrays | 2017 | https://www.ncbi.nlm.nih.gov/geo/query/acc.cgi?acc=GSE18229 | 20813035 |
| 53 | Breast cancer | GSE18229_GPL887 | 16546 | 94 | Agilent-012097 Human 1A Microarray (V2) G4110B (Feature Number version) | 2017 | https://www.ncbi.nlm.nih.gov/geo/query/acc.cgi?acc=GSE18229 | 20813035 |
| 54 | Breast cancer | GSE22226_GPL1708 | 18841 | 129 | Agilent-012391 Whole Human Genome Oligo Microarray G4112A (Feature Number version) | 2018 | https://www.ncbi.nlm.nih.gov/geo/query/acc.cgi?acc=GSE22226 | 22198468 |
| 55 | Breast cancer | GSE22226_GPL4133 | 19749 | 20 | Agilent-014850 Whole Human Genome Microarray 4x44K G4112F (Feature Number version) | 2018 | https://www.ncbi.nlm.nih.gov/geo/query/acc.cgi?acc=GSE22226 | 22198468 |
| 56 | Breast cancer | GSE2607_GPL1390 | 10040 | 64 | Agilent Human 1A Oligo UNC custom Microarrays | 2017 | https://www.ncbi.nlm.nih.gov/geo/query/acc.cgi?acc=GSE2607 | 16626501 |
| 57 | Breast cancer | GSE2607_GPL887 | 16546 | 50 | Agilent-012097 Human 1A Microarray (V2) G4110B (Feature Number version) | 2017 | https://www.ncbi.nlm.nih.gov/geo/query/acc.cgi?acc=GSE2607 | 16626501 |
| 58 | Breast cancer | GSE6130_GPL1390 | 10040 | 96 | Agilent Human 1A Oligo UNC custom Microarrays | 2017 | https://www.ncbi.nlm.nih.gov/geo/query/acc.cgi?acc=GSE6130 | 17525107 |
| 59 | Breast cancer | GSE6130_GPL887 | 16546 | 53 | Agilent-012097 Human 1A Microarray (V2) G4110B (Feature Number version) | 2017 | https://www.ncbi.nlm.nih.gov/geo/query/acc.cgi?acc=GSE6130 | 17525107 |
| 60 | Breast cancer | GSE19536_GPL6480 | 19595 | 114 | Agilent-014850 Whole Human Genome | 2020 | https://www.ncbi.nlm.nih.gov/geo/query/acc.cgi?acc=GSE19536 | 21364938 |

| | | | | | | | | |
|---|---|---|---|---|---|---|---|---|
| | | | | | Microarray 4x44K G4112F (Probe Name version) | | | |
| 61 | Breast cancer | Caldas_2007 | 14708 | 135 | Agilent Human 1A 60-mer Oligo Microarray | 2011 | https://pubmed.ncbi.nlm.nih.gov/16936776/ | 16936776 |
| 62 | Breast cancer | GSE16987 | 18196 | 161 | Illumina humanRef-8 v2.0 expression beadchip | 2015 | https://www.ncbi.nlm.nih.gov/geo/query/acc.cgi?acc=GSE16987 | 21939527 |
| 63 | Breast cancer | GSE12071 | 12697 | 46 | SWEGENE H_v3.0.1 35K | 2012 | https://www.ncbi.nlm.nih.gov/geo/query/acc.cgi?acc=GSE12071 | 18778486 |
| 64 | Breast cancer | GSE9893 | 14819 | 155 | MLRG Human 21K V12.0 | 2015 | https://www.ncbi.nlm.nih.gov/geo/query/acc.cgi?acc=GSE9893 | 18347175 |
| 65 | Breast cancer | GSE6577 | 6742 | 88 | Swegene Human 27K RAP UniGene188 array | 2019 | https://www.ncbi.nlm.nih.gov/geo/query/acc.cgi?acc=GSE6577 | 17404078 |
| 66 | Breast cancer | GSE10510 | 19852 | 152 | DKFZ Division of Molecular Genome Analysis Human Operon 4.0 oligo Array 35k | 2013 | https://www.ncbi.nlm.nih.gov/geo/query/acc.cgi?acc=GSE10510 | 18592372 |
| 67 | Breast cancer | GSE37181 | 25438 | 123 | Illumina HumanWG-6 v3.0 expression beadchip | 2019 | https://www.ncbi.nlm.nih.gov/geo/query/acc.cgi?acc=GSE37181 | 24853384 |
| 68 | Breast cancer | GSE159956 | 12201 | 295 | Rosetta (Merck) GEL Breast Tumor Profiles | 2021 | https://www.ncbi.nlm.nih.gov/geo/query/acc.cgi?acc=GSE159956 | NA |
| 69 | Breast cancer | GSE22133_GPL5345 | 8427 | 359 | SWEGENE H_v2.1.1 55K | 2012 | https://www.ncbi.nlm.nih.gov/geo/query/acc.cgi?acc=GSE22133 | 20576095 |
| 70 | Breast cancer | GSE22219 | 21438 | 216 | Illumina humanRef-8 v1.0 expression beadchip | 2013 | https://www.ncbi.nlm.nih.gov/geo/query/acc.cgi?acc=GSE22219 | 21737487 |
| 71 | Breast cancer | GSE1379 | 11728 | 60 | Arcturus 22k human oligonucleotide microarray | 2012 | https://www.ncbi.nlm.nih.gov/geo/query/acc.cgi?acc=GSE1379 | 15193263 |

| | | | | | | | | |
|---|---|---|---|---|---|---|---|---|
| 72 | Breast cancer | GSE45725 | 18630 | 340 | Illumina HumanRef-8 v3.0 expression beadchip | 2017 | https://www.ncbi.nlm.nih.gov/geo/query/acc.cgi?acc=GSE45725 | 24996446 |
| 73 | Breast cancer | GSE175692 | 771 | 184 | nCounter Breast Cancer 360 Panel | 2021 | https://www.ncbi.nlm.nih.gov/geo/query/acc.cgi?acc=GSE175692 | 34051058 |
| 74 | Breast cancer | E_TABM_158 | 12888 | 100 | Affymetrix High Throughput Array U133AA of Av2 | 2014 | https://www.ebi.ac.uk/arrayexpress/experiments/E-TABM-158 | 17157792 |
| 75 | Breast cancer | GSE11121 | 12547 | 200 | Affymetrix Human Genome U133A Array | 2020 | https://www.ncbi.nlm.nih.gov/geo/query/acc.cgi?acc=GSE11121 | 18593943 |
| 76 | Breast cancer | GSE12093 | 12547 | 136 | Affymetrix Human Genome U133A Array | 2018 | https://www.ncbi.nlm.nih.gov/geo/query/acc.cgi?acc=GSE12093 | 18821012 |
| 77 | Breast cancer | GSE12276 | 21653 | 204 | Affymetrix HGU | 2019 | https://www.ncbi.nlm.nih.gov/geo/query/acc.cgi?acc=GSE12276 | 19421193 |
| 78 | Breast cancer | GSE1456_GPL96 | 12547 | 159 | Affymetrix HGU | 2018 | https://www.ncbi.nlm.nih.gov/geo/query/acc.cgi?acc=GSE1456 | 16280042 |
| 79 | Breast cancer | GSE146558 | 21653 | 109 | Affymetrix Human Genome U133 Plus 2.0 Array | 2021 | https://www.ncbi.nlm.nih.gov/geo/query/acc.cgi?acc=GSE146558 | 34387660 |
| 80 | Breast cancer | GSE158309 | 12547 | 461 | Affymetrix Human Genome U133A Array | 2020 | https://www.ncbi.nlm.nih.gov/geo/query/acc.cgi?acc=GSE158309 | 33003293 |
| 81 | Breast cancer | GSE16446 | 21653 | 120 | Affymetrix Human Genome U133 Plus 2.0 Array | 2019 | https://www.ncbi.nlm.nih.gov/geo/query/acc.cgi?acc=GSE16446 | 21422418 |
| 82 | Breast cancer | GSE17705 | 12547 | 298 | Affymetrix Human Genome U133A Array | 2018 | https://www.ncbi.nlm.nih.gov/geo/query/acc.cgi?acc=GSE17705 | 20697068 |

| | | | | | | | | |
|---|---|---|---|---|---|---|---|---|
| 83 | Breast cancer | GSE19615 | 21653 | 115 | Affymetrix Human Genome U133 Plus 2.0 Array | 2019 | https://www.ncbi.nlm.nih.gov/geo/query/acc.cgi?acc=GSE19615 | 20098429 |
| 84 | Breast cancer | GSE2034 | 12547 | 286 | Affymetrix Human Genome U134A Array | 2018 | https://www.ncbi.nlm.nih.gov/geo/query/acc.cgi?acc=GSE2034 | 17420468, 15721472 |
| 85 | Breast cancer | GSE20711 | 21653 | 90 | Affymetrix Human Genome U133 Plus 2.0 Array | 2021 | https://www.ncbi.nlm.nih.gov/geo/query/acc.cgi?acc=GSE20711 | 21910250 |
| 86 | Breast cancer | GSE21653 | 21653 | 266 | Affymetrix Human Genome U133 Plus 2.0 Array | 2019 | https://www.ncbi.nlm.nih.gov/geo/query/acc.cgi?acc=GSE21653 | 20490655 |
| 87 | Breast cancer | GSE25055 | 12547 | 310 | Affymetrix Human Genome U136A Array | 2018 | https://www.ncbi.nlm.nih.gov/geo/query/acc.cgi?acc=GSE25055 | 21558518 |
| 88 | Breast cancer | GSE25065 | 12547 | 198 | Affymetrix Human Genome U137A Array | 2018 | https://www.ncbi.nlm.nih.gov/geo/query/acc.cgi?acc=GSE25065 | 21558518 |
| 89 | Breast cancer | GSE2603 | 12547 | 121 | Affymetrix Human Genome U133A Array | 2020 | https://www.ncbi.nlm.nih.gov/geo/query/acc.cgi?acc=GSE2603 | 16049480 |
| 90 | Breast cancer | GSE2990 | 12547 | 189 | Affymetrix Human Genome U133A Array | 2018 | https://www.ncbi.nlm.nih.gov/geo/query/acc.cgi?acc=GSE2990 | 16478745, 17401012 |
| 91 | Breast cancer | GSE3494_GPL96 | 12547 | 251 | Affymetrix Human Genome U133A Array | 2019 | https://www.ncbi.nlm.nih.gov/geo/query/acc.cgi?acc=GSE3494 | 16141321 |
| 92 | Breast cancer | GSE37751 | 20202 | 108 | Affymetrix Human Gene 1.0 ST Array | 2019 | https://www.ncbi.nlm.nih.gov/geo/query/acc.cgi?acc=GSE37751 | 24316975 |
| 93 | Breast cancer | GSE42568 | 21653 | 121 | Affymetrix Human Genome U133 Plus 2.0 Array | 2019 | https://www.ncbi.nlm.nih.gov/geo/query/acc.cgi?acc=GSE42568 | 23740839 |

| | | | | | | | | |
|---|---|---|---|---|---|---|---|---|
| 94 | Breast cancer | GSE45255 | 12547 | 139 | Affymetrix Human Genome U133A Array | 2018 | https://www.ncbi.nlm.nih.gov/geo/query/acc.cgi?acc=GSE45255 | 23618380 |
| 95 | Breast cancer | GSE48390 | 21653 | 81 | Affymetrix Human Genome U133 Plus 2.0 Array | 2019 | https://www.ncbi.nlm.nih.gov/geo/query/acc.cgi?acc=GSE48390 | 24098497 |
| 96 | Breast cancer | GSE4922_GPL96 | 12547 | 249 | Affymetrix Human Genome U133A Array | 2018 | https://www.ncbi.nlm.nih.gov/geo/query/acc.cgi?acc=GSE4922 | 17079448 |
| 97 | Breast cancer | GSE53031 | 19040 | 167 | Affymetrix Human Genome U219 Array | 2019 | https://www.ncbi.nlm.nih.gov/geo/query/acc.cgi?acc=GSE53031 | 24825746 |
| 98 | Breast cancer | GSE5327 | 12547 | 58 | Affymetrix Human Genome U134A Array | 2018 | https://www.ncbi.nlm.nih.gov/geo/query/acc.cgi?acc=GSE5327 | 17420468 |
| 99 | Breast cancer | GSE58644 | 20202 | 321 | Affymetrix Gene1.0ST | 2018 | https://www.ncbi.nlm.nih.gov/geo/query/acc.cgi?acc=GSE58644 | 25284793 |
| 100 | Breast cancer | GSE58812 | 21653 | 107 | Affymetrix Human Genome U133 Plus 2.0 Array | 2019 | https://www.ncbi.nlm.nih.gov/geo/query/acc.cgi?acc=GSE58812 | 25887482 |
| 101 | Breast cancer | GSE6532_GPL570 | 21653 | 87 | Affymetrix Human Genome U133 Plus 2.0 Array | 2019 | https://www.ncbi.nlm.nih.gov/geo/query/acc.cgi?acc=GSE6532 | 17401012 |
| 102 | Breast cancer | GSE6532_GPL96 | 12547 | 138 | Affymetrix Human Genome U133A Array | 2019 | https://www.ncbi.nlm.nih.gov/geo/query/acc.cgi?acc=GSE6532 | 17401012 |
| 103 | Breast cancer | GSE7390 | 12547 | 198 | Affymetrix Human Genome U135A Array | 2018 | https://www.ncbi.nlm.nih.gov/geo/query/acc.cgi?acc=GSE7390 | 17545524 |
| 104 | Breast cancer | GSE9195 | 21653 | 77 | Affymetrix Human Genome U133 Plus 2.0 Array | 2019 | https://www.ncbi.nlm.nih.gov/geo/query/acc.cgi?acc=GSE9195 | 18498629 |

| | | | | | | | | |
|---|---|---|---|---|---|---|---|---|
| 105 | Breast cancer | ICGC_BRCA_FR | 20039 | 99 | Affymetrix Human U133 Plus 2.0 | 2019 | https://dcc.icgc.org/releases/current/Projects/BRCA-FR | NA |
| 106 | Breast cancer | ICGC_BRCA_KR | 26730 | 50 | HTSeq | 2019 | https://dcc.icgc.org/releases/current/Projects/BRCA-KR | NA |
| 107 | Breast cancer | GSE46563 | 25438 | 94 | Illumina HumanWG-6 v3.0 expression beadchip | 2019 | https://www.ncbi.nlm.nih.gov/geo/query/acc.cgi?acc=GSE46563 | 24599057 |
| 108 | Breast cancer | TCGA_BRCA | 34849 | 1050 | TCGA-BRCA | 2021 | https://www.ncbi.nlm.nih.gov/geo/query/acc.cgi?acc= | 23000897 |
| 109 | Breast cancer | METABRIC | 25233 | 2136 | METABRIC | 2021 | https://ega-archive.org/studies/EGAS00000000083 | 22522925 |
| 110 | Breast cancer | NCI | 4111 | 99 | In-house cDNA | 2003 | http://www.ncbi.nlm.nih.gov/pubmed/?term=12917485 | 12917485 |
| 111 | Breast cancer | NKI | 13114 | 337 | Agilent | 2002 | http://www.ncbi.nlm.nih.gov/pubmed/?term=12490681; http://www.ncbi.nlm.nih.gov/pubmed/?term=11823860 | 12490681, 11823860 |
| 112 | Breast cancer | UCSF | 6497 | 162 | In-house cDNA | 2007 | http://www.ncbi.nlm.nih.gov/pubmed/?term=17428335 | 17428335, 14612510 |
| 113 | Burkitt's lymphoma | GSE4475 | 12547 | 221 | Affymetrix Human Genome U133A Array | 2018 | https://www.ncbi.nlm.nih.gov/geo/query/acc.cgi?acc=GSE4475 | 16760442 |
| 114 | Cervical cancer | GSE44001 | 20818 | 300 | Illumina HumanHT-12 WG-DASL V4.0 R2 expression beadchip | 2017 | https://www.ncbi.nlm.nih.gov/geo/query/acc.cgi?acc=GSE44001 | 24145113 |
| 115 | Cervical cancer | TCGA_CESC | 34849 | 283 | HTSeq | 2021 | https://portal.gdc.cancer.gov/projectsTCGA-CESC | 28112728 |

| | | | | | | | | |
|---|---|---|---|---|---|---|---|---|
| 116 | Cholangiocarcinoma | E_MTAB_6389 | 25359 | 109 | Affymetrix GeneChip HTA-2_0 - Exon Level - HTA-2_0.r1.PsrsJucs.ps probesets | 2019 | https://www.ebi.ac.uk/arrayexpress/experiments/E-MTAB-6389 | NA |
| 117 | Cholangiocarcinoma | TCGA_CHOL | 34849 | 36 | HTSeq | 2021 | https://portal.gdc.cancer.gov/projectsTCGA-CHOL | 28658632 |
| 118 | Chromophobe renal cell carcinoma | TCGA_KICH | 34849 | 64 | HTSeq | 2021 | https://portal.gdc.cancer.gov/projectsTCGA-KICH | 25155756 |
| 119 | Chronic lymphocytic leukemia | GSE22762_GPL570 | 21653 | 107 | Affymetrix Human Genome U133 Plus 2.0 Array | 2019 | https://www.ncbi.nlm.nih.gov/geo/query/acc.cgi?acc=GSE22762 | 21625232 |
| 120 | Chronic lymphocytic leukemia | GSE22762_GPL96 | 12547 | 44 | Affymetrix Human Genome U133A Array | 2019 | https://www.ncbi.nlm.nih.gov/geo/query/acc.cgi?acc=GSE22762 | 21625232 |
| 121 | Chronic lymphocytic leukemia | GSE22762_GPL97 | 10603 | 44 | Affymetrix Human Genome U133B Array | 2019 | https://www.ncbi.nlm.nih.gov/geo/query/acc.cgi?acc=GSE22762 | 21625232 |
| 122 | Colon cancer | GSE28722 | 15240 | 125 | Rosetta custom human 23K array | 2012 | https://www.ncbi.nlm.nih.gov/geo/query/acc.cgi?acc=GSE28722 | 21251323 |
| 123 | Colon cancer | E_MTAB_863 | 3102 | 212 | Affymetrix Custom Array - Almac Diagnostics Colorectal Cancer DSATM research tool ADXCRCG2a520319 | 2014 | https://www.ebi.ac.uk/arrayexpress/experiments/E-MTAB-863 | 22067406 |

| | | | | | | | | |
|---|---|---|---|---|---|---|---|---|
| 124 | Colon cancer | E_MTAB_864 | 3102 | 144 | Affymetrix Custom Array - Almac Diagnostics Colorectal Cancer DSATM research tool ADXCRCG2a520319 | 2014 | https://www.ebi.ac.uk/arrayexpress/experiments/E-MTAB-864 | 22067406 |
| 125 | Colon cancer | GSE16125_GPL5175 | 497 | 36 | Affymetrix Human Exon 1.0 ST Array | 2019 | https://www.ncbi.nlm.nih.gov/geo/query/acc.cgi?acc=GSE16125 | 19672874 |
| 126 | Colon cancer | GSE17536 | 21653 | 177 | Affymetrix Human Genome U133 Plus 2.0 Array | 2020 | https://www.ncbi.nlm.nih.gov/geo/query/acc.cgi?acc=GSE17536 | 19914252 |
| 127 | Colon cancer | GSE17537 | 21653 | 55 | Affymetrix Human Genome U133 Plus 2.0 Array | 2020 | https://www.ncbi.nlm.nih.gov/geo/query/acc.cgi?acc=GSE17537 | 19914252 |
| 128 | Colon cancer | GSE29621 | 21653 | 65 | Affymetrix Human Genome U133 Plus 2.0 Array | 2019 | https://www.ncbi.nlm.nih.gov/geo/query/acc.cgi?acc=GSE29621 | 22362069 |
| 129 | Colon cancer | GSE31595 | 21653 | 37 | Affymetrix Human Genome U133 Plus 2.0 Array | 2019 | https://www.ncbi.nlm.nih.gov/geo/query/acc.cgi?acc=GSE31595 | 22710688 |
| 130 | Colon cancer | GSE38832 | 21653 | 122 | Affymetrix Human Genome U133 Plus 2.0 Array | 2019 | https://www.ncbi.nlm.nih.gov/geo/query/acc.cgi?acc=GSE38832 | 25320007 |
| 131 | Colon cancer | GSE39582 | 21653 | 585 | Affymetrix Human Genome U133 Plus 2.0 Array | 2021 | https://www.ncbi.nlm.nih.gov/geo/query/acc.cgi?acc=GSE39582 | 23700391 |

| | | | | | | | | |
|---|---|---|---|---|---|---|---|---|
| 132 | Colon cancer | TCGA_COAD | 34849 | 430 | HTSeq | 2021 | https://portal.gdc.cancer.gov/projectsTCGA-COAD | 22810696 |
| 133 | Colorectal cancer | GSE12945 | 12547 | 62 | Affymetrix Human Genome U134A Array | 2018 | https://www.ncbi.nlm.nih.gov/geo/query/acc.cgi?acc=GSE12945 | 19399471 |
| 134 | Colorectal cancer | GSE14333 | 21653 | 290 | Affymetrix Human Genome U133 Plus 2.0 Array | 2019 | https://www.ncbi.nlm.nih.gov/geo/query/acc.cgi?acc=GSE14333 | 19996206 |
| 135 | Colorectal cancer | GSE30378 | 497 | 95 | Affymetrix Human Exon 1.0 ST Array | 2019 | https://www.ncbi.nlm.nih.gov/geo/query/acc.cgi?acc=GSE30378 | 22213796 |
| 136 | Colorectal cancer | GSE41258 | 12547 | 182 | Affymetrix Human Genome U134A Array | 2019 | https://www.ncbi.nlm.nih.gov/geo/query/acc.cgi?acc=GSE41258 | 19359472 |
| 137 | Colorectal cancer | GSE24549_GPL5175 | 497 | 83 | Affymetrix Human Exon 1.0 ST Array | 2019 | https://www.ncbi.nlm.nih.gov/geo/query/acc.cgi?acc=GSE24549 | 21619627 |
| 138 | Colorectal cancer | GSE24550_GPL5175 | 497 | 90 | Affymetrix Human Exon 1.0 ST Array | 2019 | https://www.ncbi.nlm.nih.gov/geo/query/acc.cgi?acc=GSE24550 | 21619627 |
| 139 | Cutaneous melanoma | TCGA_SKCM | 34849 | 98 | HTSeq | 2021 | https://portal.gdc.cancer.gov/projectsTCGA-SKCM | 26091043 |
| 140 | Diffuse large B cell lymphoma | E_MEXP_3488 | 20088 | 43 | Affymetrix GeneChip Human Exon 1.0 ST Array version 1 | 2014 | https://www.ebi.ac.uk/arrayexpress/experiments/E-MEXP-3488 | 25381134 |
| 141 | Diffuse large B cell lymphoma | E_TABM_346 | 12547 | 53 | Affymetrix GeneChip Human Genome HG-U133A | 2014 | https://www.ebi.ac.uk/arrayexpress/experiments/E-TABM-346 | 18615101 |
| 142 | Diffuse large B cell lymphoma | GSE10846 | 21653 | 420 | Affymetrix Human Genome U133 Plus 2.0 Array | 2019 | https://www.ncbi.nlm.nih.gov/geo/query/acc.cgi?acc=GSE10846 | 19038878 |

| | | | | | | | | |
|---|---|---|---|---|---|---|---|---|
| 143 | Diffuse large B cell lymphoma | GSE23501 | 21653 | 69 | Affymetrix Human Genome U133 Plus 2.0 Array | 2019 | https://www.ncbi.nlm.nih.gov/geo/query/acc.cgi?acc=GSE23501 | 20610814 |
| 144 | Diffuse large B cell lymphoma | TCGA_DLBC | 34849 | 46 | HTSeq | 2021 | https://portal.gdc.cancer.gov/projectsTCGA-DLBC | 29641966 |
| 145 | Endometrial Carcinoma | TCGA_UCEC | 34849 | 533 | HTSeq | 2021 | https://portal.gdc.cancer.gov/projectsTCGA-UCEC | 23636398 |
| 146 | Esophageal adenocarcinomas | GSE19417 | 19882 | 70 | Rosetta/Merck Human 44k 1.1 microarray | 2012 | https://www.ncbi.nlm.nih.gov/geo/query/acc.cgi?acc=GSE19417 | 20621683 |
| 147 | Esophageal adenocarcinomas | TCGA_ESCA | 34849 | 151 | HTSeq | 2021 | https://portal.gdc.cancer.gov/projectsTCGA-ESCA | 28052061 |
| 148 | Follicular lymphoma | GSE16131_GPL96 | 12547 | 184 | Affymetrix Human Genome U133A Array | 2018 | https://www.ncbi.nlm.nih.gov/geo/query/acc.cgi?acc=GSE16131 | 19471018 |
| 149 | Follicular lymphoma | GSE16131_GPL97 | 10603 | 184 | Affymetrix Human Genome U133B Array | 2018 | https://www.ncbi.nlm.nih.gov/geo/query/acc.cgi?acc=GSE16131 | 19471018 |
| 150 | Gastric cancer | GSE26253 | 13509 | 432 | Illumina HumanRef-8 WG-DASL v3.0 | 2019 | https://www.ncbi.nlm.nih.gov/geo/query/acc.cgi?acc=GSE26253 | 24598828 |
| 151 | Gastric cancer | GSE15459 | 21653 | 192 | Affymetrix Human Genome U133 Plus 2.0 Array | 2019 | https://www.ncbi.nlm.nih.gov/geo/query/acc.cgi?acc=GSE15459 | 19798449 |
| 152 | Gastric cancer | GSE34942 | 21653 | 56 | Affymetrix Human Genome U133 Plus 2.0 Array | 2019 | https://www.ncbi.nlm.nih.gov/geo/query/acc.cgi?acc=GSE34942 | 25053715 |

| | | | | | | | | |
|---|---|---|---|---|---|---|---|---|
| 153 | Gastric cancer | GSE62254 | 21653 | 300 | Affymetrix Human Genome U133 Plus 2.0 Array | 2019 | https://www.ncbi.nlm.nih.gov/geo/query/acc.cgi?acc=GSE62254 | 25894828 |
| 154 | Gastric cancer | TCGA_STAD | 34849 | 348 | HTSeq | 2021 | https://portal.gdc.cancer.gov/projectsTCGA-STAD | 25079317 |
| 155 | Head and neck squamous cell carcinoma | GSE10300 | 21653 | 44 | Affymetrix Human Genome U133 Plus 2.0 Array | 2019 | https://www.ncbi.nlm.nih.gov/geo/query/acc.cgi?acc=GSE10300 | 19117988 |
| 156 | Head and neck squamous cell carcinoma | GSE65858 | 22000 | 270 | Illumina HumanHT-12 V4.0 expression beadchip | 2018 | https://www.ncbi.nlm.nih.gov/geo/query/acc.cgi?acc=GSE65858 | 26095926 |
| 157 | Head and neck squamous cell carcinoma | E_MTAB_1328 | 21653 | 89 | Affymetrix Human Genome U133 Plus 2.0 Array | 2014 | https://www.ebi.ac.uk/arrayexpress/experiments/E-MTAB-1328 | 23757353 |
| 158 | Head and neck squamous cell carcinoma | TCGA_HNSC | 34849 | 494 | HTSeq | 2021 | https://portal.gdc.cancer.gov/projectsTCGA-HNSC | 25631445 |
| 159 | Hepatocellular Carcinoma | ICGC_LIRI_JP | 22913 | 445 | HTSeq | 2019 | https://dcc.icgc.org/releases/current/Projects/LIRI-JP | NA |
| 160 | Hepatocellular Carcinoma | GSE10141 | 6100 | 80 | Human 6k Transcriptionally Informative Gene Panel for DASL | 2020 | https://www.ncbi.nlm.nih.gov/geo/query/acc.cgi?acc=GSE10141 | 18923165 |
| 161 | Hepatocellular Carcinoma | GSE17856 | 14313 | 52 | Agilent-014850 Whole Human Genome Microarray 4x44K G4112F | 2019 | https://www.ncbi.nlm.nih.gov/geo/query/acc.cgi?acc=GSE17856 | 20380719 |

| | | | | | | | | |
|---|---|---|---|---|---|---|---|---|
| 162 | Hepatocellular Carcinoma | GSE27150 | 2348 | 81 | State Key Lab Homo sapien 2.6K | 2012 | https://www.ncbi.nlm.nih.gov/geo/query/acc.cgi?acc=GSE27150 | NA |
| 163 | Hepatocellular Carcinoma | GSE14520 | 12742 | 221 | Affymetrix HT Human Genome U133A Array | 2021 | https://www.ncbi.nlm.nih.gov/geo/query/acc.cgi?acc=GSE14520 | 21159642 |
| 164 | Hepatocellular Carcinoma | TCGA_LIHC | 34849 | 363 | HTSeq | 2021 | https://portal.gdc.cancer.gov/projectsTCGA-LIHC | 28622513 |
| 165 | Laryngeal cancer | GSE27020 | 12547 | 109 | Affymetrix Human Genome U133A Array | 2018 | https://www.ncbi.nlm.nih.gov/geo/query/acc.cgi?acc=GSE27020 | 23950933 |
| 166 | Lung cancer | GSE41271 | 25438 | 275 | Illumina HumanWG-6 v3.0 expression beadchip | 2019 | https://www.ncbi.nlm.nih.gov/geo/query/acc.cgi?acc=GSE41271 | 23449933 |
| 167 | Lung cancer | GSE3141 | 21653 | 111 | Affymetrix Human Genome U133 Plus 2.0 Array | 2019 | https://www.ncbi.nlm.nih.gov/geo/query/acc.cgi?acc=GSE3141 | 16273092 |
| 168 | Lung cancer | GSE30219 | 21653 | 307 | Affymetrix Human Genome U133 Plus 2.0 Array | 2019 | https://www.ncbi.nlm.nih.gov/geo/query/acc.cgi?acc=GSE30219 | 23698379 |
| 169 | Lung cancer | GSE31547 | 12547 | 50 | Affymetrix Human Genome U133A Array | 2018 | https://www.ncbi.nlm.nih.gov/geo/query/acc.cgi?acc=GSE31547 | NA |
| 170 | Lung cancer-Lung adenocarcinoma | GSE26939 | 17108 | 116 | Agilent-UNC-custom-4X44K | 2012 | https://www.ncbi.nlm.nih.gov/geo/query/acc.cgi?acc=GSE26939 | 22590557 |
| 171 | Lung cancer-Lung adenocarcinoma | GSE72094 | 22115 | 442 | Rosetta/Merck Human RSTA Custom Affymetrix 2.0 microarray | 2018 | https://www.ncbi.nlm.nih.gov/geo/query/acc.cgi?acc=GSE72094 | 26477306 |

| | | | | | | | | |
|---|---|---|---|---|---|---|---|---|
| 172 | Lung cancer-Lung adenocarcinoma | GSE13213 | 18479 | 117 | Agilent-014850 Whole Human Genome Microarray 4x44K G4112F | 2019 | https://www.ncbi.nlm.nih.gov/geo/query/acc.cgi?acc=GSE13213 | 19414676 |
| 173 | Lung cancer-Lung adenocarcinoma | GSE11969 | 16624 | 163 | Agilent Homo sapiens 21.6K custom array | 2013 | https://www.ncbi.nlm.nih.gov/geo/query/acc.cgi?acc=GSE11969 | 16549822 |
| 174 | Lung cancer-Lung adenocarcinoma | GSE5843 | 13111 | 48 | PRHU05-S1-0006 (PC Human Operon v2 21k) | 2015 | https://www.ncbi.nlm.nih.gov/geo/query/acc.cgi?acc=GSE5843 | 17504995 |
| 175 | Lung cancer-Lung adenocarcinoma | E_MTAB_923 | 21653 | 103 | Affymetrix GeneChip Human Genome U133 Plus 2.0 | 2014 | https://www.ebi.ac.uk/arrayexpress/experiments/E-MTAB-923 | 22914773 |
| 176 | Lung cancer-Lung adenocarcinoma | GSE31210 | 21653 | 246 | Affymetrix Human Genome U133 Plus 2.0 Array | 2019 | https://www.ncbi.nlm.nih.gov/geo/query/acc.cgi?acc=GSE31210 | 22080568 |
| 177 | Lung cancer-Lung adenocarcinoma | GSE68465 | 12547 | 462 | Affymetrix Human Genome U133A Array | 2018 | https://www.ncbi.nlm.nih.gov/geo/query/acc.cgi?acc=GSE68465 | 18641660 |
| 178 | Lung cancer-Lung | GSE68571 | 5249 | 96 | Affymetrix Human Full Length HuGeneFL Array | 2016 | https://www.ncbi.nlm.nih.gov/geo/query/acc.cgi?acc=GSE68571 | 12118244 |

| | | | | | | | | |
|---|---|---|---|---|---|---|---|---|
| | adenocarcinoma | | | | | | | |
| 179 | Lung cancer-Lung adenocarcinoma | GSE63459 | 18630 | 65 | Illumina HumanRef-8 v3.0 expression beadchip | 2017 | https://www.ncbi.nlm.nih.gov/geo/query/acc.cgi?acc=GSE63459 | 26134223 |
| 180 | Lung cancer-Lung adenocarcinoma | TCGA_LUAD | 34849 | 497 | HTSeq | 2021 | https://portal.gdc.cancer.gov/projectsTCGA-LUAD | 25079552 |
| 181 | Lung cancer-Lung squamous cell carcinoma | GSE17710 | 17108 | 56 | Agilent-UNC-custom-4X44K | 2012 | https://www.ncbi.nlm.nih.gov/geo/query/acc.cgi?acc=GSE17710 | 20643781 |
| 182 | Lung cancer-Lung squamous cell carcinoma | E_MTAB_2435 | 21653 | 93 | Affymetrix GeneChip Human Genome U133 Plus 2.0 | 2018 | https://www.ebi.ac.uk/arrayexpress/experiments/E-MTAB-2435 | 25189482 |
| 183 | Lung cancer-Non-small cell lung cancer | GSE11117 | 9661 | 56 | Novachip human 34.5k | 2012 | https://www.ncbi.nlm.nih.gov/geo/query/acc.cgi?acc=GSE11117 | 19833826 |
| 184 | Lung cancer-Non-small cell lung cancer | GSE8894 | 21653 | 138 | Affymetrix Human Genome U133 Plus 2.0 Array | 2019 | https://www.ncbi.nlm.nih.gov/geo/query/acc.cgi?acc=GSE8894 | 19010856 |
| 185 | Lung cancer-Non-small cell lung cancer | GSE157009 | 21653 | 249 | Affymetrix Human Genome U133 Plus 2.0 Array | 2020 | https://www.ncbi.nlm.nih.gov/geo/query/acc.cgi?acc=GSE157009 | 32717408 |

| | | | | | | | | |
|---|---|---|---|---|---|---|---|---|
| 186 | Lung cancer-Non-small cell lung cancer | GSE4573 | 12547 | 129 | Affymetrix Human Genome U133A Array | 2018 | https://www.ncbi.nlm.nih.gov/geo/query/acc.cgi?acc=GSE4573 | 16885343 |
| 187 | Lung cancer-Non-small cell lung cancer | GSE42127 | 25438 | 176 | Illumina HumanWG-6 v3.0 expression beadchip | 2020 | https://www.ncbi.nlm.nih.gov/geo/query/acc.cgi?acc=GSE42127 | 23357979 |
| 188 | Lung cancer-Non-small cell lung cancer | GSE14814 | 12547 | 133 | Affymetrix Human Genome U133A Array | 2018 | https://www.ncbi.nlm.nih.gov/geo/query/acc.cgi?acc=GSE14814 | 20823422 |
| 189 | Lung cancer-Non-small cell lung cancer | GSE157010 | 21653 | 235 | Affymetrix Human Genome U133 Plus 2.0 Array | 2020 | https://www.ncbi.nlm.nih.gov/geo/query/acc.cgi?acc=GSE157010 | 32717408 |
| 190 | Lung cancer-Non-small cell lung cancer | GSE19188 | 21653 | 156 | Affymetrix Human Genome U133 Plus 2.0 Array | 2019 | https://www.ncbi.nlm.nih.gov/geo/query/acc.cgi?acc=GSE19188 | 20421987 |
| 191 | Lung cancer-Non-small cell lung cancer | GSE37745 | 21653 | 196 | Affymetrix Human Genome U133 Plus 2.0 Array | 2021 | https://www.ncbi.nlm.nih.gov/geo/query/acc.cgi?acc=GSE37745 | 23032747 |
| 192 | Lung cancer-Non-small cell lung cancer | GSE50081 | 21653 | 181 | Affymetrix Human Genome U133 Plus 2.0 Array | 2019 | https://www.ncbi.nlm.nih.gov/geo/query/acc.cgi?acc=GSE50081 | 24305008 |
| 193 | Lung cancer-Non-small cell lung cancer | TCGA_LUSC | 34849 | 489 | HTSeq | 2021 | https://portal.gdc.cancer.gov/projectsTCGA-LUSC | 22960745 |

| | | | | | | | | |
|---|---|---|---|---|---|---|---|---|
| 194 | Mesothelioma | E_MTAB_1719 | 21653 | 38 | Affymetrix GeneChip Human Genome U133 Plus 2.0 | 2018 | https://www.ebi.ac.uk/arrayexpress/experiments/E-MTAB-1719 | 24443521 |
| 195 | Mesothelioma | E_MTAB_6877 | 15653 | 67 | Affymetrix Human Gene 2.0 ST Array | 2020 | https://www.ebi.ac.uk/arrayexpress/experiments/E-MTAB-6877 | 30902996 |
| 196 | Mesothelioma | TCGA_MESO | 34849 | 79 | HTSeq | 2021 | https://portal.gdc.cancer.gov/projectsTCGA-MESO | 30322867 |
| 197 | Multiple myeloma | GSE9782_GPL96 | 12547 | 264 | Affymetrix Human Genome U133A Array | 2018 | https://www.ncbi.nlm.nih.gov/geo/query/acc.cgi?acc=GSE9782 | 17185464 |
| 198 | Multiple myeloma | GSE9782_GPL97 | 10603 | 264 | Affymetrix Human Genome U133B Array | 2018 | https://www.ncbi.nlm.nih.gov/geo/query/acc.cgi?acc=GSE9782 | 17185464 |
| 199 | Multiple myeloma | E_MTAB_1038 | 20202 | 73 | Affymetrix GeneChip Human Gene 1.0 ST Array | 2014 | https://www.ebi.ac.uk/arrayexpress/experiments/E-MTAB-1038 | 27234807 |
| 200 | Multiple myeloma | E_MTAB_4032 | 20202 | 151 | Affymetrix Human Gene 1.0 ST Array | 2016 | https://www.ebi.ac.uk/arrayexpress/experiments/E-MTAB-4032 | 27234807 |
| 201 | Multiple myeloma | GSE136337 | 21653 | 426 | Affymetrix Human Genome U133 Plus 2.0 Array | 2020 | https://www.ncbi.nlm.nih.gov/geo/query/acc.cgi?acc=GSE136337 | 33147277 |
| 202 | Multiple myeloma | GSE24080 | 21653 | 559 | Affymetrix Human Genome U133 Plus 2.0 Array | 2019 | https://www.ncbi.nlm.nih.gov/geo/query/acc.cgi?acc=GSE24080 | 20064235 |
| 203 | Multiple myeloma | GSE57317 | 21653 | 55 | Affymetrix Human Genome U133 Plus 2.0 Array | 2019 | https://www.ncbi.nlm.nih.gov/geo/query/acc.cgi?acc=GSE57317 | 25079174 |
| 204 | Multiple myeloma | MMRF_CoMMpass | 34849 | 787 | HTSeq | 2018 | https://portal.gdc.cancer.gov/projects | 27417553 |

| | | | | | | | | |
|---|---|---|---|---|---|---|---|---|
| 205 | Neuroblastoma | E_MTAB_8248 | 19860 | 223 | Agilent-020382 Human Custom Microarray 44k | 2019 | https://www.ebi.ac.uk/arrayexpress/experiments/E-MTAB-8248 | 32291317 |
| 206 | Neuroblastoma | TARGET_NBL | 58387 | 142 | Illumina HiSeq | 2019 | https://xenabrowser.net/datapages/ | 23334666 |
| 207 | Neuroblastoma | GSE62564 | 23465 | 498 | Illumina HiSeq 2000 (Homo sapiens) | 2019 | https://www.ncbi.nlm.nih.gov/geo/query/acc.cgi?acc=GSE62564 | 25150839 |
| 208 | Oral cancer | ICGC_ORCA_IN | 24003 | 40 | HTSeq | 2019 | https://dcc.icgc.org/releases/current/Projects/ORCA-IN | NA |
| 209 | Osteosarcoma | TARGET_OS | 58387 | 87 | Illumina HiSeq | 2019 | https://xenabrowser.net/datapages/ | 29228567 |
| 210 | Osteosarcoma | GSE21257 | 24996 | 53 | Illumina human-6 v2.0 expression beadchip (using nuIDs as identifier) | 2012 | https://www.ncbi.nlm.nih.gov/geo/query/acc.cgi?acc=GSE21257 | 21372215 |
| 211 | Ovarian cancer | GSE17260 | 19595 | 110 | Agilent-014850 Whole Human Genome Microarray 4x44K G4112F (Probe Name version) | 2019 | https://www.ncbi.nlm.nih.gov/geo/query/acc.cgi?acc=GSE17260 | 20300634 |
| 212 | Ovarian cancer | ICGC_OV_AU | 24650 | 111 | HTSeq | 2019 | https://dcc.icgc.org/releases/current/Projects/OV-AU | NA |
| 213 | Ovarian cancer | GSE73614 | 19584 | 107 | Agilent-014850 Whole Human Genome Microarray 4x44K G4112F | 2019 | https://www.ncbi.nlm.nih.gov/geo/query/acc.cgi?acc=GSE73614 | 27016234 |
| 214 | Ovarian cancer | GSE49997 | 16751 | 204 | ABI Human Genome Survey Microarray Version 2 | 2016 | https://www.ncbi.nlm.nih.gov/geo/query/acc.cgi?acc=GSE49997 | 22497737 |

| | | | | | | | | |
|---|---|---|---|---|---|---|---|---|
| 215 | Ovarian cancer | GSE13876 | 15971 | 415 | Operon human v3 ~35K 70-mer two-color oligonucleotide microarrays | 2013 | https://www.ncbi.nlm.nih.gov/geo/query/acc.cgi?acc=GSE13876 | 19192944 |
| 216 | Ovarian cancer | GSE14764 | 12547 | 80 | Affymetrix Human Genome U133A Array | 2019 | https://www.ncbi.nlm.nih.gov/geo/query/acc.cgi?acc=GSE14764 | 19294737 |
| 217 | Ovarian cancer | GSE18520 | 21653 | 63 | Affymetrix Human Genome U133 Plus 2.0 Array | 2019 | https://www.ncbi.nlm.nih.gov/geo/query/acc.cgi?acc=GSE18520 | 19962670 |
| 218 | Ovarian cancer | GSE19829 _GPL570 | 21653 | 28 | Affymetrix Human Genome U133 Plus 2.0 Array | 2019 | https://www.ncbi.nlm.nih.gov/geo/query/acc.cgi?acc=GSE19829 | 20547991 |
| 219 | Ovarian cancer | GSE19829 _GPL8300 | 8619 | 42 | Affymetrix Human Genome U95 Version 2 Array | 2019 | https://www.ncbi.nlm.nih.gov/geo/query/acc.cgi?acc=GSE19829 | 20547991 |
| 220 | Ovarian cancer | GSE23554 | 12547 | 28 | Affymetrix Human Genome U133A Array | 2018 | https://www.ncbi.nlm.nih.gov/geo/query/acc.cgi?acc=GSE23554 | 21849418 |
| 221 | Ovarian cancer | GSE26712 | 87 | 195 | Affymetrix Human Genome U134A Array | 2018 | https://www.ncbi.nlm.nih.gov/geo/query/acc.cgi?acc=GSE26712 | 18593951 |
| 222 | Ovarian cancer | GSE30161 | 21653 | 58 | Affymetrix Human Genome U133 Plus 2.0 Array | 2019 | https://www.ncbi.nlm.nih.gov/geo/query/acc.cgi?acc=GSE30161 | 22348014 |
| 223 | Ovarian cancer | GSE31245 | 8619 | 57 | Affymetrix Human Genome U95 Version 2 Array | 2018 | https://www.ncbi.nlm.nih.gov/geo/query/acc.cgi?acc=GSE31245 | 16204010 |

| | | | | | | | | |
|---|---|---|---|---|---|---|---|---|
| 224 | Ovarian cancer | GSE63885 | 21653 | 101 | Affymetrix Human Genome U133 Plus 2.0 Array | 2019 | https://www.ncbi.nlm.nih.gov/geo/query/acc.cgi?acc=GSE63885 | 24478986 |
| 225 | Ovarian cancer | GSE9891 | 21653 | 285 | Affymetrix Human Genome U133 Plus 2.0 Array | 2019 | https://www.ncbi.nlm.nih.gov/geo/query/acc.cgi?acc=GSE9891 | 18698038 |
| 226 | Ovarian cancer | GSE32062_GPL6480 | 19595 | 260 | Agilent-014850 Whole Human Genome Microarray 4x44K G4112F (Probe Name version) | 2019 | https://www.ncbi.nlm.nih.gov/geo/query/acc.cgi?acc=GSE32062 | 22241791 |
| 227 | Ovarian cancer | GSE32063 | 19595 | 40 | Agilent-014850 Whole Human Genome Microarray 4x44K G4112F | 2019 | https://www.ncbi.nlm.nih.gov/geo/query/acc.cgi?acc=GSE32063 | 22241791 |
| 228 | Ovarian cancer | TCGA_OV | 34849 | 353 | HTSeq | 2021 | https://portal.gdc.cancer.gov/projectsTCGA-OV | 21720365 |
| 229 | Pancreatic ductal adenocarcinoma | ICGC_PACA_CA | 25731 | 262 | HTSeq | 2019 | https://dcc.icgc.org/releases/current/Projects/PACA-CA | NA |
| 230 | Pancreatic endocrine neoplasms | ICGC_PAEN_AU | 24650 | 33 | HTSeq | 2019 | https://dcc.icgc.org/releases/current/Projects/PAEN-AU | NA |
| 231 | Pancreatic ductal adenocarcinoma | GSE21501 | 19749 | 132 | Agilent-014850 Whole Human Genome Microarray 4x44K G4112F | 2013 | https://www.ncbi.nlm.nih.gov/geo/query/acc.cgi?acc=GSE21501 | 20644708 |

| | | | | | | | | |
|---|---|---|---|---|---|---|---|---|
| 232 | Pancreatic ductal adenocarcinoma | ICGC_PACA_AU | 24650 | 92 | HTSeq | 2019 | https://dcc.icgc.org/releases/current/Projects/PACA-AU | 21436628 |
| 233 | Pancreatic ductal adenocarcinoma | GSE71729 | 19749 | 357 | Agilent-014850 Whole Human Genome Microarray 4x44K G4112F | 2015 | https://www.ncbi.nlm.nih.gov/geo/query/acc.cgi?acc=GSE71729 | 26343385 |
| 234 | pancreatic ductal adenocarcinoma | E_MEXP_2780 | 21653 | 26 | Affymetrix GeneChip Human Genome U133 Plus 2.0 | 2014 | https://www.ebi.ac.uk/arrayexpress/experiments/E-MEXP-2780 | 22615549 |
| 235 | pancreatic ductal adenocarcinoma | E_MTAB_6134 | 19040 | 309 | Affymetrix Human Genome U219 Array | 2018 | https://www.ebi.ac.uk/arrayexpress/experiments/E-MTAB-6134 | 30165049 |
| 236 | Pancreatic ductal adenocarcinoma | GSE28735 | 20202 | 90 | Affymetrix Human Gene 1.0 ST Array | 2018 | https://www.ncbi.nlm.nih.gov/geo/query/acc.cgi?acc=GSE28735 | 22363658 |
| 237 | Pancreatic ductal adenocarcinoma | GSE57495 | 22115 | 63 | Rosetta/Merck Human RSTA Custom Affymetrix 2.0 microarray | 2018 | https://www.ncbi.nlm.nih.gov/geo/query/acc.cgi?acc=GSE57495 | 26247463 |
| 238 | Pancreatic ductal | GSE62452 | 20202 | 130 | Affymetrix Human Gene 1.0 ST Array | 2018 | https://www.ncbi.nlm.nih.gov/geo/query/acc.cgi?acc=GSE62452 | 27197190 |

| | | | | | | | | |
|---|---|---|---|---|---|---|---|---|
| | adenocarcinoma | | | | | | | |
| 239 | Pancreatic ductal adenocarcinoma | GSE78229 | 20202 | 50 | Affymetrix Human Gene 1.0 ST Array | 2018 | https://www.ncbi.nlm.nih.gov/geo/query/acc.cgi?acc=GSE78229 | 27401251 |
| 240 | Pancreatic ductal adenocarcinoma | GSE85916 | 19040 | 80 | Affymetrix Human Genome U219 Array | 2019 | https://www.ncbi.nlm.nih.gov/geo/query/acc.cgi?acc=GSE85916 | NA |
| 241 | Pancreatic ductal adenocarcinoma | GSE79668 | 24658 | 51 | Illumina HiSeq 2000 (Homo sapiens) | 2019 | https://www.ncbi.nlm.nih.gov/geo/query/acc.cgi?acc=GSE79668 | 27282075 |
| 242 | Pancreatic ductal adenocarcinoma | TCGA_PAAD | 34849 | 176 | HTSeq | 2021 | https://portal.gdc.cancer.gov/projectsTCGA-PAAD | 28810144 |
| 243 | Pheochromocytoma and paraganglioma | TCGA_PCPG | 34849 | 175 | HTSeq | 2021 | https://portal.gdc.cancer.gov/projectsTCGA-PCPG | 28162975 |
| 244 | Prostate cancer | GSE70768 | 22000 | 199 | Illumina HumanHT-12 V4.0 expression beadchip | 2018 | https://www.ncbi.nlm.nih.gov/geo/query/acc.cgi?acc=GSE70768 | 26501111 |
| 245 | Prostate cancer | GSE70769 | 22000 | 94 | Illumina HumanHT-12 V4.0 expression beadchip | 2018 | https://www.ncbi.nlm.nih.gov/geo/query/acc.cgi?acc=GSE70769 | 26501111 |

| | | | | | | | | |
|---|---|---|---|---|---|---|---|---|
| 246 | Prostate cancer | GSE16560 | 6100 | 281 | Human 6k Transcriptionally Informative Gene Panel for DASL | 2013 | https://www.ncbi.nlm.nih.gov/geo/query/acc.cgi?acc=GSE16560 | 20233430 |
| 247 | Prostate cancer | GSE26022 | 522 | 192 | Illumina Custom Prostate Cancer DASL Panel 1.5K expression beadchip | 2012 | https://www.ncbi.nlm.nih.gov/geo/query/acc.cgi?acc=GSE26022 | NA |
| 248 | Prostate cancer | GSE26242 | 522 | 96 | Illumina Custom Prostate Cancer DASL Panel 1.5K expression beadchip | 2012 | https://www.ncbi.nlm.nih.gov/geo/query/acc.cgi?acc=GSE26242 | NA |
| 249 | Prostate cancer | TCGA_PRAD | 34849 | 481 | HTSeq | 2021 | https://portal.gdc.cancer.gov/projectsTCGA-PRAD | 26544944 |
| 250 | Prostate cancer | ICGC_PRAD_FR | 22187 | 25 | HTSeq | 2019 | https://dcc.icgc.org/releases/current/Projects/PRAD-FR | NA |
| 251 | Rectum adenocarcinoma | TCGA_READ | 34849 | 154 | HTSeq | 2021 | https://portal.gdc.cancer.gov/projectsTCGA-READ | 22810696 |
| 252 | Renal carcinoma | GSE167573 | 55630 | 76 | HiSeq X Ten (Homo sapiens) | 2021 | https://www.ncbi.nlm.nih.gov/geo/query/acc.cgi?acc=GSE167573 | 34489456 |
| 253 | Renal carcinoma | ICGC_RECA_EU | 23698 | 136 | HTSeq | 2019 | https://dcc.icgc.org/releases/current/Projects/RECA-EU | NA |
| 254 | Renal carcinoma-Clear-cell renal cell carcinoma | E_MTAB_1980 | 22682 | 101 | Agilent Human Gene Expression 4x44K v2 Microarray 026652 G4845A | 2014 | https://www.ebi.ac.uk/arrayexpress/experiments/E-MTAB-1980 | 23797736 |

| | | | | | | | | |
|---|---|---|---|---|---|---|---|---|
| 255 | Renal carcinoma-Clear-cell renal cell carcinoma | GSE29609 | 18841 | 39 | Agilent-012391 Whole Human Genome Oligo Microarray G4112A | 2018 | https://www.ncbi.nlm.nih.gov/geo/query/acc.cgi?acc=GSE29609 | 22626276 |
| 256 | Renal carcinoma-Clear-cell renal cell carcinoma | E_MTAB_3218 | 19040 | 59 | Affymetrix Human Genome U219 Array | 2021 | https://www.ebi.ac.uk/arrayexpress/experiments/E-MTAB-3218 | 27169994 |
| 257 | Renal carcinoma-Clear-cell renal cell carcinoma | E_MTAB_3267 | 20202 | 59 | Affymetrix GeneChip Human Gene 1.0 ST Array | 2018 | https://www.ebi.ac.uk/arrayexpress/experiments/E-MTAB-3267 | 25583177 |
| 258 | Renal carcinoma-Clear-cell renal cell carcinoma | TCGA_KIRC | 34849 | 522 | HTSeq | 2021 | https://portal.gdc.cancer.gov/projectsTCGA-KIRC | 23792563 |
| 259 | Renal carcinoma-Renal papillary cell carcinoma | TCGA_KIRP | 34849 | 284 | HTSeq | 2021 | https://portal.gdc.cancer.gov/projectsTCGA-KIRP | 26536169 |
| 260 | Rhabdomyosarcoma | E_TABM_1202 | 21653 | 101 | Affymetrix GeneChip Human Genome U133 Plus 2.0 | 2012 | https://www.ebi.ac.uk/arrayexpress/experiments/E-TABM-1202 | 23536298 |
| 261 | Testicular germ cell tumor | TCGA_TGCT | 34849 | 133 | HTSeq | 2021 | https://portal.gdc.cancer.gov/projectsTCGA-TGCT | 29898407 |

| | | | | | | | | |
|---|---|---|---|---|---|---|---|---|
| 262 | Thymoma | TCGA_THYM | 34849 | 118 | HTSeq | 2021 | https://portal.gdc.cancer.gov/projectsTCGA-THYM | 29438696 |
| 263 | Thyroid carcinoma | TCGA_THCA | 34849 | 496 | HTSeq | 2021 | https://portal.gdc.cancer.gov/projectsTCGA-THCA | 25417114 |
| 264 | Uterine carcinosarcoma | TCGA_UCS | 34849 | 54 | HTSeq | 2021 | https://portal.gdc.cancer.gov/projectsTCGA-UCS | 28292439 |
| 265 | Uveal melanoma | GSE39717 | 21453 | 31 | Illumina humanRef-8 v1.0 expression beadchip | 2015 | https://www.ncbi.nlm.nih.gov/geo/query/acc.cgi?acc=GSE39717 | 17332290 |
| 266 | Uveal melanoma | GSE22138 | 21653 | 63 | Affymetrix Human Genome U133 Plus 2.0 Array | 2019 | https://www.ncbi.nlm.nih.gov/geo/query/acc.cgi?acc=GSE22138 | 21135111 |
| 267 | Uveal melanoma | TCGA_UVM | 34849 | 77 | HTSeq | 2021 | https://portal.gdc.cancer.gov/projectsTCGA-UVM | 29316429 |
| 268 | Wilms tumor | TARGET_WT | 58387 | 120 | Illumina HiSeq | 2019 | https://xenabrowser.net/datapages/ | 28825729 |

**Supplementary table 2.** Univariate Cox proportional hazards regression model on the overall survival of patients with breast cancer in METABRIC cohort

| Genes | Coefficient | HR | LCI | UCI | PValue | Padjusted |
|---|---|---|---|---|---|---|
| RACGAP1 | 0.376 | 1.456 | 1.316 | 1.610 | <0.001 | <0.001 |
| CDCA5 | 0.240 | 1.271 | 1.190 | 1.358 | <0.001 | <0.001 |
| TROAP | 0.319 | 1.376 | 1.260 | 1.502 | <0.001 | <0.001 |
| STIP1 | 0.421 | 1.524 | 1.353 | 1.716 | <0.001 | <0.001 |
| PKMYT1 | 0.381 | 1.464 | 1.313 | 1.631 | <0.001 | <0.001 |
| UBE2C | 0.189 | 1.208 | 1.144 | 1.275 | <0.001 | <0.001 |
| TPX2 | 0.282 | 1.326 | 1.223 | 1.438 | <0.001 | <0.001 |
| KIF20A | 0.279 | 1.321 | 1.219 | 1.432 | <0.001 | <0.001 |
| CFL1 | 0.559 | 1.749 | 1.485 | 2.060 | <0.001 | <0.001 |
| SUSD3 | -0.142 | 0.868 | 0.833 | 0.905 | <0.001 | <0.001 |
| CLIC6 | -0.106 | 0.900 | 0.872 | 0.928 | <0.001 | <0.001 |
| FAM83D | 0.272 | 1.313 | 1.211 | 1.424 | <0.001 | <0.001 |
| LARP1 | 0.613 | 1.846 | 1.539 | 2.216 | <0.001 | <0.001 |
| PTTG1 | 0.230 | 1.259 | 1.175 | 1.349 | <0.001 | <0.001 |
| CPT1A | 0.445 | 1.561 | 1.365 | 1.785 | <0.001 | <0.001 |
| KIF4A | 0.380 | 1.462 | 1.304 | 1.639 | <0.001 | <0.001 |
| PIGV | -0.480 | 0.619 | 0.535 | 0.715 | <0.001 | <0.001 |
| AURKA | 0.241 | 1.273 | 1.184 | 1.369 | <0.001 | <0.001 |
| FCER1A | -0.208 | 0.812 | 0.762 | 0.865 | <0.001 | <0.001 |
| PPIL3 | -0.500 | 0.606 | 0.521 | 0.706 | <0.001 | <0.001 |
| PLK1 | 0.531 | 1.701 | 1.445 | 2.003 | <0.001 | <0.001 |
| PRC1 | 0.238 | 1.268 | 1.179 | 1.365 | <0.001 | <0.001 |
| UHRF1 | 0.248 | 1.282 | 1.187 | 1.384 | <0.001 | <0.001 |
| CKAP2L | 0.317 | 1.373 | 1.244 | 1.516 | <0.001 | <0.001 |
| PARP3 | -0.348 | 0.706 | 0.633 | 0.787 | <0.001 | <0.001 |
| VEGFA | 0.281 | 1.325 | 1.212 | 1.447 | <0.001 | <0.001 |
| MELK | 0.216 | 1.241 | 1.159 | 1.328 | <0.001 | <0.001 |
| GTSE1 | 0.381 | 1.464 | 1.298 | 1.651 | <0.001 | <0.001 |
| GSK3B | 0.359 | 1.432 | 1.278 | 1.604 | <0.001 | <0.001 |
| CENPO | 0.606 | 1.832 | 1.510 | 2.224 | <0.001 | <0.001 |
| FEN1 | 0.292 | 1.339 | 1.220 | 1.471 | <0.001 | <0.001 |
| AK3 | -0.438 | 0.645 | 0.561 | 0.742 | <0.001 | <0.001 |
| TUBA1B | 0.483 | 1.620 | 1.387 | 1.893 | <0.001 | <0.001 |
| CCNB2 | 0.209 | 1.232 | 1.152 | 1.318 | <0.001 | <0.001 |
| TRIM4 | -0.377 | 0.686 | 0.607 | 0.775 | <0.001 | <0.001 |
| SHMT2 | 0.359 | 1.432 | 1.275 | 1.609 | <0.001 | <0.001 |
| NUSAP1 | 0.250 | 1.284 | 1.184 | 1.392 | <0.001 | <0.001 |
| ENC1 | 0.319 | 1.376 | 1.241 | 1.525 | <0.001 | <0.001 |
| CDC20 | 0.169 | 1.184 | 1.121 | 1.251 | <0.001 | <0.001 |
| RBBP8 | -0.245 | 0.782 | 0.723 | 0.847 | <0.001 | <0.001 |
| CENPE | 0.317 | 1.374 | 1.239 | 1.523 | <0.001 | <0.001 |

| | | | | | | |
|---|---|---|---|---|---|---|
| USP30 | 0.786 | 2.194 | 1.697 | 2.838 | <0.001 | <0.001 |
| FGD3 | -0.170 | 0.844 | 0.798 | 0.892 | <0.001 | <0.001 |
| ESPL1 | 0.551 | 1.735 | 1.447 | 2.081 | <0.001 | <0.001 |
| OMD | -0.239 | 0.787 | 0.727 | 0.853 | <0.001 | <0.001 |
| S100P | 0.086 | 1.090 | 1.059 | 1.122 | <0.001 | <0.001 |
| CENPL | 0.437 | 1.548 | 1.338 | 1.791 | <0.001 | <0.001 |
| GSTM3 | -0.188 | 0.829 | 0.778 | 0.882 | <0.001 | <0.001 |
| ZWINT | 0.285 | 1.329 | 1.208 | 1.462 | <0.001 | <0.001 |
| PTTG3P | 0.217 | 1.242 | 1.155 | 1.335 | <0.001 | <0.001 |
| STAT5B | -0.428 | 0.652 | 0.565 | 0.752 | <0.001 | <0.001 |
| LSR | 0.396 | 1.485 | 1.301 | 1.696 | <0.001 | <0.001 |
| AURKB | 0.228 | 1.256 | 1.163 | 1.356 | <0.001 | <0.001 |
| BCL2 | -0.190 | 0.827 | 0.776 | 0.882 | <0.001 | <0.001 |
| RALGAPB | 0.482 | 1.619 | 1.377 | 1.904 | <0.001 | <0.001 |
| DYNLRB2 | -0.245 | 0.782 | 0.720 | 0.850 | <0.001 | <0.001 |
| UTP23 | 0.392 | 1.481 | 1.297 | 1.691 | <0.001 | <0.001 |
| GSTK1 | -0.394 | 0.674 | 0.590 | 0.770 | <0.001 | <0.001 |
| FANCD2 | 0.341 | 1.407 | 1.253 | 1.579 | <0.001 | <0.001 |
| GPI | 0.335 | 1.398 | 1.248 | 1.566 | <0.001 | <0.001 |

**Abbreviations:** HR, hazard ratio; LCI, lower limit of confidence interval; UCI, upper limit of confidence interval.

**Supplementary table 3.** Differences between ABAT high expression group and ABAT low expression group

| **Variable** | **ABAT expression levels** | | **P value** |
|---|---|---|---|
| | **High expression (N=1068)** | **Low expression (N=1068)** | |
| **ER** | | | |
| negative | 46(4.3%) | 394(36.9%) | <0.001 |
| positive | 984(92.1%) | 524(49.1%) | |
| Missing | 38(3.6%) | 150(14.0%) | |
| **Tumor size** | | | |
| Mean (SD) | 2.53(1.36) | 2.72(1.72) | 0.00536 |
| Median [Min, Max] | 2.20[0,18.0] | 2.30[0,18.2] | |
| Missing | 30(2.8%) | 134(12.5%) | |
| **Node status** | | | |
| Mean (SD) | 0.444(0.497) | 0.513(0.500) | 0.00207 |
| Median [Min, Max] | 0[0,1.00] | 1.00[0,1.00] | |
| Missing | 19(1.8%) | 125(11.7%) | |
| **Age(years)** | | | |

| | | | |
|---|---|---|---|
| Mean (SD) | 63.2(12.5) | 59.0(13.2) | <0.001 |
| Median [Min, Max] | 64.3[26.4,92.1] | 59.8[21.9,96.3] | |
| Missing | 2(0.2%) | 11(1.0%) | |
| **Grade** | | | |
| Mean (SD) | 2.20(0.657) | 2.64(0.556) | <0.001 |
| Median [Min, Max] | 2.00[1.00,3.00] | 3.00[1.00,3.00] | |
| Missing | 75(7.0%) | 159(14.9%) | |

**Supplementary table 4.** Comparison of clinical features in the GSE31210 cohort (training set)

| Variable | Subtype 1 (N=86) | Subtype 2 (N=40) | Subtype 3 (N=70) | Subtype 4 (N=30) | P value |
|---|---|---|---|---|---|
| **Age (years)** | | | | | |
| Mean (SD) | 59.0(8.05) | 60.0(7.17) | 60.8(6.29) | 58.0(8.05) | 0.28 |
| Median [Min, Max] | 60.0[30.0,76.0] | 61.0[38.0,71.0] | 61.0[35.0,72.0] | 59.0[39.0,69.0] | |
| **Cluster** | | | | | |
| Cluster1 | 24(27.9%) | 23(57.5%) | 0(0%) | 0(0%) | <0.001 |
| Cluster2 | 8(9.3%) | 0(0%) | 20(28.6%) | 4(13.3%) | |
| Missing | 54(62.8%) | 17(42.5%) | 50(71.4%) | 26(86.7%) | |
| **Gender** | | | | | |
| Female | 48(55.8%) | 11(27.5%) | 43(61.4%) | 19(63.3%) | 0.00292 |
| Male | 38(44.2%) | 29(72.5%) | 27(38.6%) | 11(36.7%) | |
| **MYC** | | | | | |
| High | 4(4.7%) | 4(10.0%) | 3(4.3%) | 6(20.0%) | 0.127 |
| Low | 81(94.2%) | 36(90.0%) | 66(94.3%) | 24(80.0%) | |
| ND | 1(1.2%) | 0(0%) | 1(1.4%) | 0(0%) | |
| **Pathological stage** | | | | | |
| IA | 35(40.7%) | 8(20.0%) | 47(67.1%) | 24(80.0%) | <0.001 |
| IB | 17(19.8%) | 13(32.5%) | 20(28.6%) | 4(13.3%) | |
| II | 34(39.5%) | 19(47.5%) | 3(4.3%) | 2(6.7%) | |
| **Gene alteration status** | | | | | |
| ALK-fusion+ | 9(10.5%) | 2(5.0%) | 0(0%) | 0(0%) | <0.001 |
| EGFR mutation+ | 43(50.0%) | 10(25.0%) | 49(70.0%) | 25(83.3%) | |
| EGFR/KRAS/ALK- | 23(26.7%) | 21(52.5%) | 20(28.6%) | 4(13.3%) | |
| KRAS mutation+ | 11(12.8%) | 7(17.5%) | 1(1.4%) | 1(3.3%) | |
| **Smoking status** | | | | | |
| Ever-smoker | 44(51.2%) | 31(77.5%) | 22(31.4%) | 14(46.7%) | <0.001 |
| Never-smoker | 42(48.8%) | 9(22.5%) | 48(68.6%) | 16(53.3%) | |

**Abbreviations:** ND, not determined

**Supplementarytable5.**Comparison of clinical features in the TCGA-LUAD cohort (validations et)

| Variable | Subtype1 (N=7) | Subtype2 (N=375) | Subtype3 (N=84) | Subtype4 (N=31) | P value |
|---|---|---|---|---|---|
| **Age(years)** | | | | | |
| Mean (SD) | 66.6(8.54) | 64.7(10.3) | 67.3(9.36) | 66.3(8.74) | |
| Median [Min, Max] | 70.0[56.0,78.0] | 65.0[33.0,88.0] | 67.0[41.0,86.0] | 67.0[48.0,80.0] | 0.181 |
| Missing | 0(0%) | 7(1.9%) | 3(3.6%) | 0(0%) | |
| **Gender** | | | | | |
| Female | 3(42.9%) | 190(50.7%) | 54(64.3%) | 22(71.0%) | 0.0268 |
| Male | 4(57.1%) | 185(49.3%) | 30(35.7%) | 9(29.0%) | |
| **Ethnicity** | | | | | |
| Not Hispanic or Latino | 5(71.4%) | 278(74.1%) | 70(83.3%) | 25(80.6%) | 0.859 |
| Hispanic or Latino | 0(0%) | 6(1.6%) | 1(1.2%) | 0(0%) | |
| Missing | 2(28.6%) | 91(24.3%) | 13(15.5%) | 6(19.4%) | |
| **Race** | | | | | |
| Black or African American | 1(14.3%) | 38(10.1%) | 8(9.5%) | 4(12.9%) | |
| White | 5(71.4%) | 286(76.3%) | 70(83.3%) | 23(74.2%) | 0.681 |
| American Indian or Alaska native | 0(0%) | 1(0.3%) | 0(0%) | 0(0%) | |
| Asian | 0(0%) | 4(1.1%) | 1(1.2%) | 2(6.5%) | |
| Missing | 1(14.3%) | 46(12.3%) | 5(6.0%) | 2(6.5%) | |
| **Number pack years smoked** | | | | | |
| Mean (SD) | 42.1(13.2) | 44.4(28.4) | 34.9(23.1) | 30.0(15.5) | 0.0218 |
| Median[ Min, Max] | 40.3[28.0,60.0] | 40.0[1.00,154] | 30.0[0.150,100] | 35.0[4.50,50.0] | |
| Missing | 3(42.9%) | 114(30.4%) | 27(32.1%) | 12(38.7%) | |
| **Tobacco smoking history** | | | | | |
| Mean (SD) | 2.57(0.976) | 2.84(1.10) | 2.75(1.01) | 2.77(1.06) | |
| Median [Min, Max] | 3.00[1.00,4.00] | 3.00[1.00,5.00] | 3.00[1.00,4.00] | 3.00[1.00,4.00] | 0.852 |
| Missing | 0(0%) | 11(2.9%) | 3(3.6%) | 0(0%) | |
| **Pathologic stage** | | | | | |
| Stage I | 3(42.9%) | 186(49.6%) | 59(70.2%) | 19(61.3%) | 0.0131 |
| Stage II | 2(28.6%) | 97(25.9%) | 9(10.7%) | 10(32.3%) | |
| Stage III | 1(14.3%) | 68(18.1%) | 9(10.7%) | 2(6.5%) | |

| | | | | |
|---|---|---|---|---|
| Stage IV | 1(14.3%) | 20(5.3%) | 4(4.8%) | 0(0%) |
| Missing | 0(0%) | 4(1.1%) | 3(3.6%) | 0(0%) |

**Supplementary table 6.** Parameter setting range of each model used in this model

| Learner | Parameter | Parameter type | Parameter range |
|---|---|---|---|
| Random forest | mtry | Integer | 1,15 |
| | Node size | Discrete | 3,5,8,10,15,18,20 |
| | ntree | Discrete | 500,1000,1500,2000 |
| glmboost | Node depth | Integer | 5,20 |
| | mstop | Integer | 1e2,1e3 |
| | nu | Discrete | 0.05,0.1,0.3,0.5,0.8,1 |
| coxboost | stepno | Integer | lower=50, upper=200 |
| Elasticnet | alpha | Numeric | lower=0, upper=1 |
| | s | Numeric | lower=0.001, upper=30 |
| Ridge | s | Numeric | lower=0, upper=20 |
| Lasso | s | Numeric | lower=0, upper=20 |
